# How many lives can be saved? A global view on the impact of testing, herd immunity and demographics on COVID-19 fatality rates

**DOI:** 10.1101/2020.04.29.20084400

**Authors:** Miguel Sánchez-Romero, Vanessa di Lego, Alexia Prskawetz, Bernardo L Queiroz

## Abstract

In this work, we assess the global impact of COVID-19 showing how demographic factors, testing policies and herd immunity are key for saving lives. We extend a standard epidemiological *SEIR* model in order to: (a) identify the role of demographics (population size and population age distribution) on COVID-19 fatality rates; (b) quantify the maximum number of lives that can be saved according to different testing strategies, different levels of herd immunity, and specific population characteristics; and (d) infer from the observed case fatality rates (CFR) what the true fatality rate might be. Different from previous *SEIR* model extensions, we implement a Bayesian Melding method in our calibration strategy which enables us to account for data limitation on the total number of deaths. We derive a distribution of the set of parameters that best replicate the observed evolution of deaths by using information from both the model and the data.

**One Sentence Summary:** Demographics factors, testing policies and herd immunity are key for quantifying the maximum number of lives that can be saved from COVID-19.

## Main Text

As the prospects for the development of a COVID-19/ SARS-CoV-2 vaccine are not in the near future, countries will have to manage the dramatic consequences of long-term lockdowns and quarantine regimes, which are the most employed non-pharmaceutical public health interventions (NPIs) that control the infection *(1–3)*. While for SARS-CoV-1 the peak viral shedding occurred after patients were very ill with respiratory symptoms that could easily be traced, identified and isolated (*1*), viral shedding for COVID-19 is long and observed in asymptomatic individuals (*3*) making it difficult to control human-to-human transmission, with many countries only realizing the spread of the disease when they were already seriously hit (*4*). With the rapid undetected spread and lack of fast response or testing availability, most countries, with few exceptions^1^, relied heavily on widespread social distance measures and lockdowns, while restricting testing to symptomatic individuals. These measures are effective to control the infection, but have important socioeconomic, psychological and health consequences, and unless herd immunity considerably develops, most likely a second wave (or waves) of infection will arise as people slowly return to their activities (*5*). In addition, due to the major role of infectious asymptomatic individuals, restricting testing to the symptomatic makes it difficult to estimate the true case fatality rates and appropriately measure the death toll, since the less tests you make, the lower the number of detected persons and the higher the case fatality rates (*6*).

These factors make these strategies unsustainable in the long-run and both country endurance to social distance measures and their effect on fatality rates will most likely depend on socioeconomic settings (*7*), detecting asymptomatic individuals (*8*), undocumented cases (*9*), demographic characteristics *(10)*, and the level of herd immunity (*11*). We account in a combined fashion for those last three factors by extending a standard epidemiological *SEIR* (Susceptible-Exposed-Infected-Removed) compartment model by (a) accounting for the age structure of the population and age-specific mortality rates, (b) explicitly modeling the mortality rate of the COVID-19 epidemics and (c) introducing isolation periods during incubation *(E)* and the infectious period *(I)* after testing (see **Fig. S1** for a schematic diagram of the *SEIR* model implemented). Different from previous *SEIR* model extensions, we implement a Bayesian Melding method in our calibration strategy, which provides an inferential framework that takes into account both model’s inputs and outputs. This is a unique feature that allows us to deal with incomplete data on deaths since we derive a distribution of the set of parameters that best replicate the observed deaths by using the information from both the model and the data *(12, 13*). The Bayesian Melding algorithm is applied to the evolution of the total number of deaths in the province of Hubei (China), since that is where the first case was reported (see details on section 1.2 of the supplementary material).

### Modeling the spread of COVID-19 and the role of asymptomatic individuals on the fatality rate

The COVID-19 outbreak is characterized by a large uncertainty on the number of people infected. The number of deaths from COVID-19 is also not exempt from limitation, being subject to underreporting as well as to overreporting, due to competing causes of death *(14)*. However, information on deaths still present less uncertainty than data on infections (15), and that is why we calibrate the epidemiological model using the evolution of the number of deaths, and account for overreporting by including all causes of death that are not COVID-19 (See **Fig S2**). In addition, COVID-19 deaths by age show a sizable age gradient *(15, 16)*, which is quite similar to the age gradient observed in standard mortality rates, making it important to acknowledge that the fatality rate from COVID-19 increases with age. To account for the age difference, we fitted through an OLS the log of the age-specific mortality rates for the coronavirus using a quadratic function by age (see **Fig. S2** in the supplementary material). As the number of infected cases by age are likely underreported *(17)*, actual mortality rates by age from COVID-19 are probably lower. Therefore, in order to control for the unknown number of infected people who were not developing symptoms (i.e. asymptomatic) and were not tested against the virus, we have introduced in the calibration process an adjustment factor (details on the model and the calibration are in the **section 1** of the supp. material).

We run our model assuming that the first COVID-19 case appeared in November 17, 2019, the first death occurred on January 11, 2020, and the epidemic curve started to flatten on February 12, 2020. After drawing a sample of two million values from the joint prior distribution of the underlying epidemic model parameters (see **section 1.2** supp. material for details), we obtain that the mean incubation period is slightly higher than the reported median incubation of 3.0 days reported for pediatric patients (*18*) and one day shorter than the average incubation period of 5.2 days reported for patients older than 50 years *(19)*. In addition, the recovery period is on average 11 days (IQR 6–15), the time the virus affects individuals is 14.5 days (IQR 7.7–19), and the transmission rate 0.432 (IQR 0.325–0.489). **Fig. S3** shows the posterior distribution of the basic reproduction number 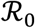. Our results show that the most probable 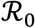 values range between 2.6-5.1, with a mean value of 4.51, very close to the basic reproduction number estimated for Wuhan (20) (for a detailed description of the calibration procedures and the epidemic parameters of the *SEIR* model see section 1.2 and Table S1 in the supp. material). In addition, our calibration shows that 56% of the infected individuals are on average asymptomatic, with the distribution of the infected individuals according to their symptoms shown in Table 1.

**Table 1.** Distribution of the infected individuals according to their symptoms. The distribution is derived following previous work (21) and assuming that among the symptomatic individuals roughly 80% are mild, 15% are severe, and 5% are critical (see supp. material 1.2).

|  | Asymptomatic | Symptomatic |  |  |  |
| --- | --- | --- | --- | --- | --- |
|  | Total | Total | Critical | Severe | Mild |
|  |  | % |  |  |  |
| 1st. Qu. | 44.65 | 55.35 | 2.77 | 8.30 | 44.28 |
| Mean | 56.13 | 43.86 | 2.19 | 6.58 | 35.09 |
| 3rd.Qu. | 68.31 | 31.68 | 1.58 | 4.75 | 25.35 |

As the role of the asymptomatic individuals is particularly important for the spread of COVID-19, taking into account that on average 56% of infected individuals are asymptomatic gives a picture of the potential for infection spread. By multiplying the modeled fatality rate by the fraction of symptomatic people among all infectious people, the distribution of the true fatality rate is derived (**Fig. S4** shows the inferred COVID-19 fatality rate after the calibration process). We obtain that the average fatality rate exceeds 1% at age 60, 5% at age 80, and 10% at age 90, which is sixty percent lower than the fatality rates that have been reported (22).

### Global fatality rates and the role of demographic factors

We adjust the epidemic curve to each country in the world (see supplementary material Section 2), which allows us to analyze the global spread and evolution of the total number of deaths throughout 365 days. Because we are particularly concerned with the aftermath of the virus, we present all results for day 365, i.e., after one year of outbreak for each country (the daily distribution is available upon request). We first estimate the impact of COVID-19 death toll in the absence of a lockdown policy and no available vaccine, assuming a well-mixed population. For comparability reasons, we apply in all countries the same initial number of imported and infected individuals, which follows a temporal Poisson process Pois(*λt*), with *λ* = 10. This implies that countries receive a monthly average of 300 infected individuals. The age of imported cases is assumed to be a random number drawn from a uniform distribution with a minimum age of 18 and a maximum of 65 which corresponds to the age of potential workers (average age of the infected individuals is 41.5 years old, which does not necessarily coincide with the average age of the population analyzed). **Fig. S6** depicts how the model is able to accurately track the pandemic evolution in different countries after the outbreak onset. Our results show that the proportion of people infected depends on country size and the fatality rate depends on the age distribution of the population. **Fig. 1** shows in panel **A** an example with extreme cases on how the transmission rate is faster in countries with a small population size like Iceland (over 340 thousand inhabitants) compared to larger countries like Brazil (over 210 million inhabitants) and China (over 1400 million inhabitants). This implies that in a context of no lockdown policy, no available vaccine, and assuming a well-mixed population, small populations get infected faster than large populations. As a consequence, the peak of the mortality rate will be sooner in small populations relative to larger ones (see **Table S2** for the average death toll of COVID-19 after one year in the world). Panel **B** shows the positive relationship between the average fatality rate and the mean age of the population for 200 countries in the world. Younger populations (mean age < 30 years) face a fatality rate of around 0.2%, whereas older populations (mean age > 40 years) face a fatality rate close to 0.9%. When comparing to the grey dots that represent deaths unrelated to COVID-19, it is clear that COVID-19 is not the main cause of death in younger populations, but has the potential to double the number of deaths in ageing populations, if no policies are implemented (see **Table S2**).

**Fig 1.**
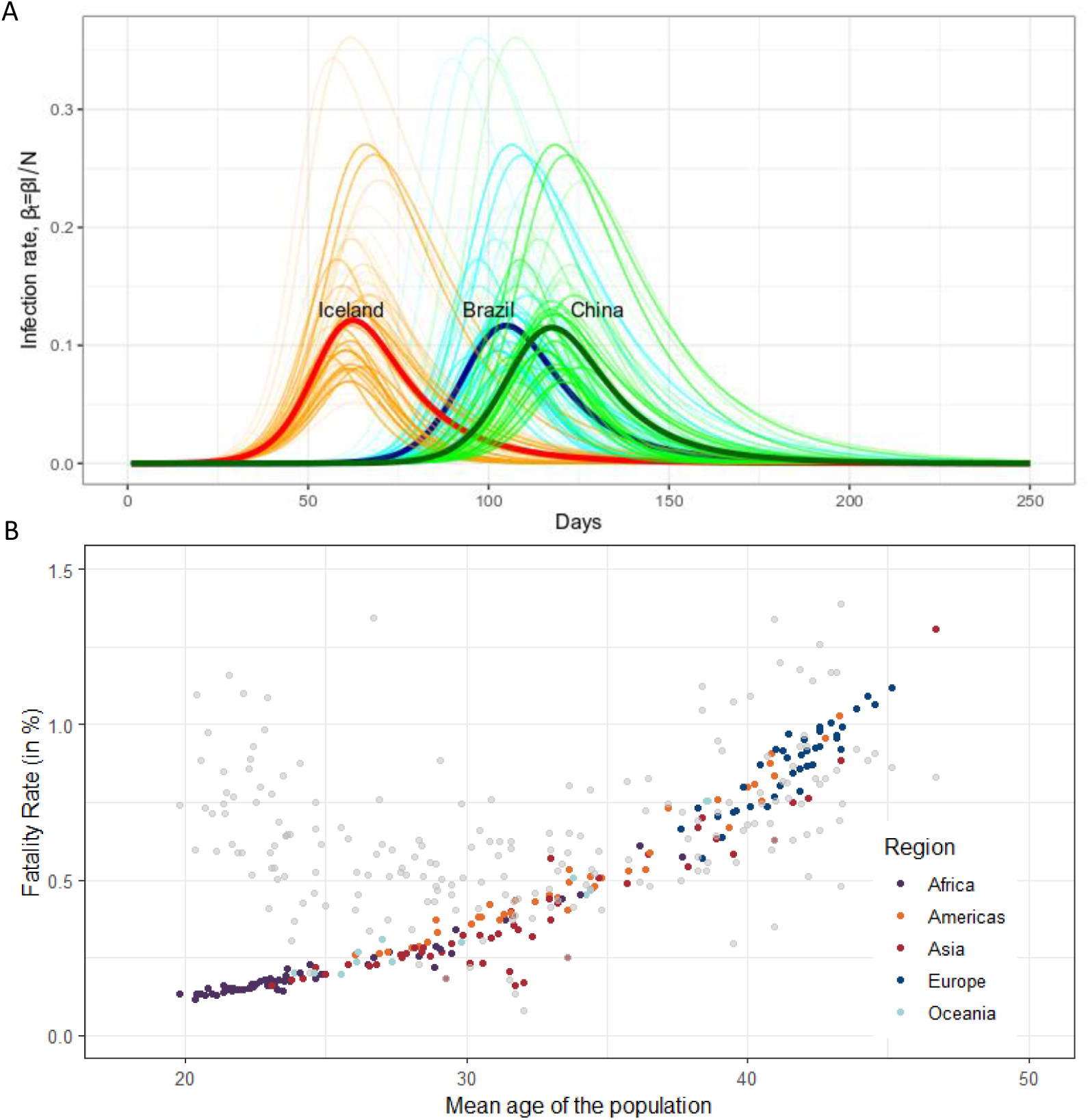
The impact of population size and mean age on the infection and fatality rate. **Panel A** is the COVID-19 infection rate in Iceland (red), Brazil (blue), and China (green). Note: Infectious rate calculated under the assumptions of no isolation measures and that the population is well mixed. **Panel B**. Relationship between the average COVID-19 fatality rate and the mean age of the population after 365 days. Gray dots depict the death rate (without COVID-19) in each country according to the mean age of the population. Source: UN Population data and authors’ calculations.

### The potential for saving lives: the joint role of testing, herd immunity and demographic factors

After estimating the global death toll in a no-policy scenario, we first include testing and isolation as the only interventions adopted to control the outbreak and its effect on saving lives. We consider both people who develop symptoms and those infectious individuals who are asymptomatic. Following evidence, we assume that among the fraction of symptomatic individuals, 80% have mild symptoms, while 20% develop critical and severe symptoms (*21*). See **Table S3** and **eq. S9-S10, section 3** for a detailed description of the alternative testing options that are implemented. **Panel A** in **Fig. 2** shows that countries with higher mean-age have higher average fatality rates than younger ones and that the higher the level of testing, the lower the fatality rates. **Panel B in Fig. 2** shows the average proportion of lives saved after one year under different testing strategies and considering population size.^2^ The death toll is reduced by 2% in large countries when severe or critical care individuals are tested and isolated. If the total coverage of testing reaches 22%, the death toll will be reduced by 9.4% in large countries. Only when all the symptomatic individuals are tested and isolated, which corresponds to almost 44% of the total infectious population, the death toll is reduced by 46% in large countries.

**Fig.2.**
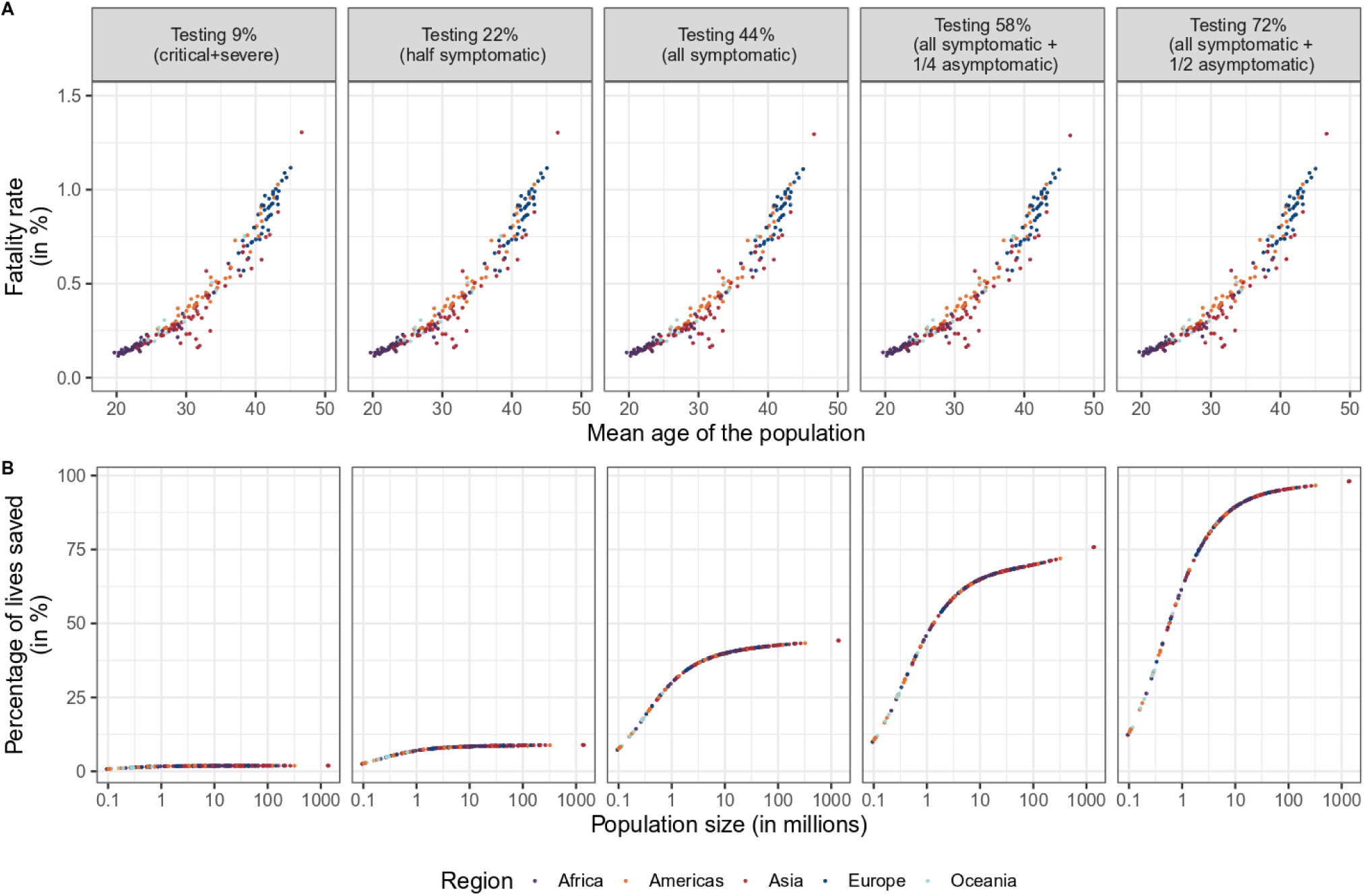
Fatality rate by mean age of the population (A) and proportion of lives saved by population size (B) at different levels of testing after one year (day=365). Source: UN Population data and authors’ calculations. Notes: Populations are assumed to be fully susceptible to COVID-19. We also analyzed the sensitivity of COVID-deaths with respect to the timing of testing by simulating eight alternative intervention days assuming that governments start testing after 45 days from the first imported and infected case. See **Table S4** for the impact of the day of intervention and the level of testing on the total number of deaths in US after 365 days.

When all the symptomatic plus half of the asymptomatic are tested in large countries, the average reduction of the death toll reaches 98%. This emphasizes the importance of testing asymptomatic individuals but also the role of population size on the potential of saving a higher proportion of lives. It is especially important to start mass testing sooner in countries with smaller populations.

After testing and isolation are included, we then add herd immunity to estimate the increasing chance of saving lives. Herd immunity and serological survey can increase the amount of lives saved both through decreasing the transmission rate and increasing the number of recovered, since people who have fully recovered from COVID-19 have antibodies in their plasma that can attack the virus and be used for treatment of severe patients (*23*, *24*). **Panel A** in **Fig. 3** shows the relationship between the average COVID-19 fatality rate and the mean age of the population for five different levels of testing and three possible degrees of herd immunity. It may come as a surprise that the fatality rate does not change much across different levels of herd immunity, but this is due to the fact that both the number of deaths and the number of infected individuals decrease, affecting the numerator and the denominator that compose the fatality rate. However, the herd immunity level *does* change the percentage of lives saved relative to the population size. **Fig. 3 panel B** shows that, for a given level of testing, the higher is the herd immunity level in a population, the greater is the percentage of lives saved (relative to the initial susceptible population). Moreover, this effect is more intense in large populations and for greater levels of testing. The fact that the fatality rate is rather insensitive to different levels of herd immunity reinforces the importance of looking not only into the fatality rates as a way to assess the impact on lives lost, but also considering the proportion of lives saved.

**Fig. 3:**
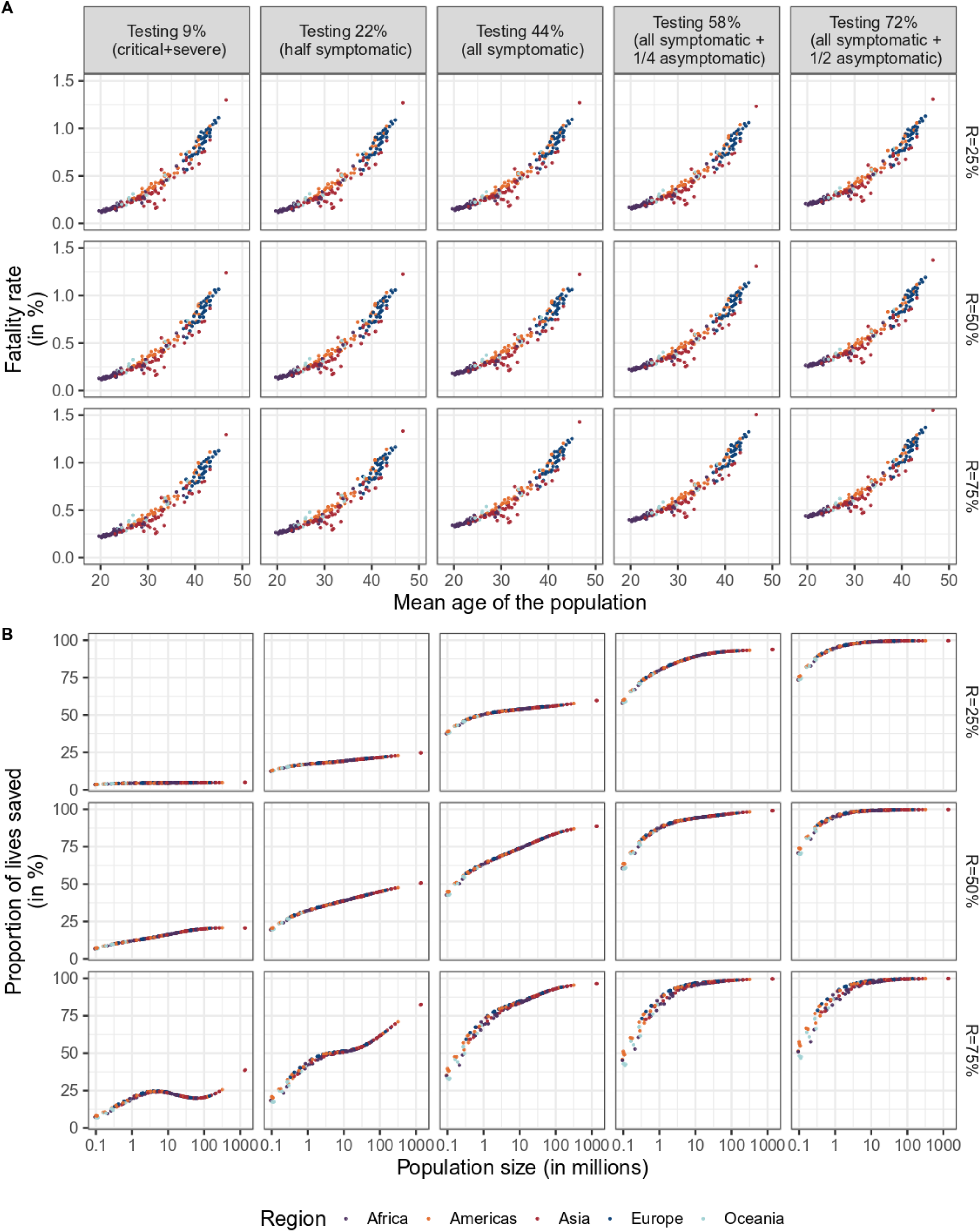
Fatality rate by mean age of the population (A) and proportion of lives saved by population size (B) at different levels of testing and herd immunity level after one year (day=365). Source: UN Population data and authors’ calculations.

### Demographics as the bridge between the (true) fatality rate and the observed CFR

As a final analysis, we distinguish between the (true) fatality rate that we derive from the model, which was used in previous sections, and the case fatality rate (CFR). We define CFR as the ratio between the total number of deaths and the total infected individuals who are detected. CFRs have been reported by countries based on the number of deaths and infected individuals they can detect. In our case, we can derive the CFR by dividing the (true) fatality rate by the fraction of infected individuals who are tested.^3^ **Fig. 4** shows our estimated average CFR by testing scenarios for eight selected countries that have different demographic characteristics (Italy, Spain, Austria, USA, China, Brazil, India, and Niger).

**Fig. 4.**
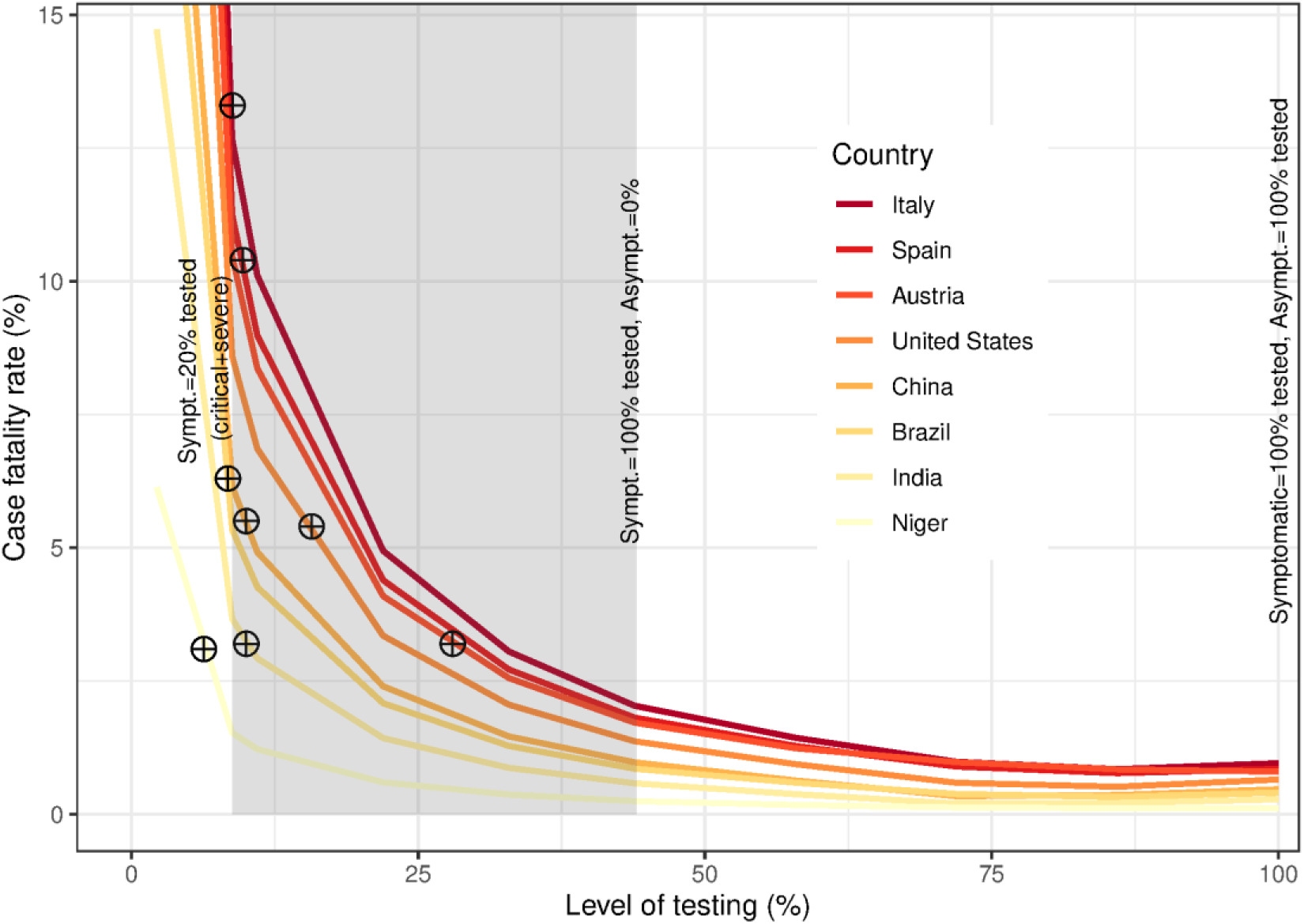
Average case fatality rate (CFR) in percentage by level of testing according to symptomatic and asymptomatic individuals. Source: Own calculations. Note: The gray area represents the level of testing for the symptomatic individuals. Specifically, the CFRs at the left-hand side of the gray area correspond to testing only 20% of symptomatic individuals, which are the critical and severe cases and a strategy adopted by many countries, while the CFR at the right-hand side of the gray area correspond to testing 100% of the symptomatic individuals. The true fatality rates and the CFRs only coincide when the level of testing is one hundred percent, which corresponds to the far-right point in the figure.

The reported CFR values by countries on April 21st, 2020, represented by the crossed circles in the figure, were 13.3% in Italy, 10.4% in Spain, 3.2% in Austria, 5.4% in US, 5.5% in China, 6.3% in Brazil, 3.2% in India, and 3.1% in Niger. The current reported CFR values for China, India, Italy, and Spain, when compared to our estimated values and given that not all the infected individuals are yet recovered or dead, suggest that these countries are mainly testing individuals who develop critical and severe symptoms. In China, India, Italy, and Spain the total number of infected people is at least ten times higher than those being reported. Rates in Brazil and Niger suggest that less than 9% of the total infected individuals are tested. Thus, in these two countries the total number of infected people is more than ten times higher than those being reported. In the US, the CFR value suggests that around 15% of the infected individuals are tested and therefore the likely total number of infected people is more than six times higher than those reported. In Austria, which is a country that started testing at high proportions, the CFR value suggests that around than 25% of the infected individuals are tested. As a result, the total number of infected individuals is at least more than four times the total infected individuals tested, which coincides with the lower bound estimated by a recent study (25). We extend the analysis to the whole world, with results detailed in the supplementary materials (see **Fig. S12** and **Table S7**). Overall, only testing the critical and severe cases implies that CFRs are above 10% in countries with a mean age of the population above 42 years and close to 2% in countries with a mean age of the population below 25 years. Once the level of testing reaches all symptomatic individuals (i.e. 44% of all infected), we observe CFR values that are lower than 2.5% in all countries. This implies that CFRs can be reduced by half by testing at least 72% of the total infected population (relative to the symptomatic individuals). Additionally, intervention day, population size, and herd immunity are factors that influence the total number of people infected, as shown in **Table S4** and **Figs. 4** and S10. However, conditional on being infected, the probability of dying from the epidemic (fatality rate) just depends on age. Thus, given the demographic characteristic of each population —e.g. the mean age of the population— we can infer through the case fatality rate the total number of people infected in each country, which is a feature that may help us shed light on the number of people infected.

## Discussion

It is fundamental to prepare for the aftermath of this global pandemic with the most flexible and wide range of strategies possible, that envisage short, mid and long-term solutions, since mitigation and suppression strategies will need to be maintained until vaccines or effective treatments become widely available *(26)*. We show that the role of testing, demographics and herd immunity are key for assessing the global impact of the COVID-19 epidemic. CFRs can be reduced by half by testing at least 72% of the total infected population and the number of infected in some countries is at least ten times higher than those being reported. Countries that test only severe and critical COVID-19 cases have CFRs above 10% in countries with a mean age of the population above 42 years and close to 2% in countries with a mean age of the population below 25 years, so younger countries experience lower impacts in terms of fatality rates relative to older ones. Smaller countries are more vulnerable to a faster spread of the virus and herd immunity is important, but affects more the proportion of lives being saved than the fatality rates. We also show how we can use demographic characteristics to infer from the case fatality rate the total number of people infected in each country, which can be a valuable tool for indirectly estimating the number of people infected.

However, we are not accounting for underlying health conditions and country-specific healthcare system capacity which affects resilience levels of countries coping with the pandemic *(27)*. Nonetheless, because the fatality rates by health characteristics are likely to present important bias at this moment, focusing on age provides more robust results. Gender differences *(28)* were also not considered and because there is an infection differential by gender and women have higher life expectancies than men, they may feel more stringent effects on savings, income support and loss of spouse or other safety nets *(29)*. Spatial flows were also not accounted for and living arrangements are precluded from the analysis, when multigenerational households might have an important impact on transmission rates *(30)*. Other important consequences of coping with the pandemic such as the increase in suicide rates among medical staff and leaders *(42)*, detrimental psychological and physical effects of isolation, and vulnerable people with preexisting mental disorders *(31, 33*) are also not the focus of our study.

As a final remark, despite the undebatable importance of testing, the recent strong worldwide demand caused a severe disruption in the production and supply of laboratory reagents, affecting test availability, particularly for low-income countries *(26, 34)*. Given the importance of testing, it is paramount that institutions make a concerted effort at increasing testing availability.

## Data Availability

All data is available in the main text or the supplementary materials, as well as in the github repository sent together with this submission. All data used in this study is public.

https://github.com/vdilego/COVID-19_SEIR_Bayesian_Melding

## Acknowledgments

We would like to acknowledge all our colleagues from the Wittgenstein Centre for Demography and Global Human Capital (Univ. Vienna, IIASA, VID/ÖAW) for their helpful comments that allowed us to improve our work. We would like to most notably thank Wolfgang Lutz, Michael Kuhn, Raya Muttarak, Thomas Sobotka, Jesus Crespo Cuaresma, and Zuzanna Brzozowska for their insightful suggestions. **Author contributions:** Miguel Sánchez-Romero: Conceptualization, Methodology, Software, Validation, Formal analysis, Investigation, Writing – Review & Editing, Visualization, Writing original draft (supp material); Vanessa di Lego: Conceptualization, Methodology, Formal analysis, Investigation, Writing original draft (main text), Writing – Review & Editing Visualization; Alexia: Conceptualization, Formal analysis, Investigation, Writing – Review & Editing; Bernardo L Queiroz: Conceptualization, Investigation, Writing – Review & Editing **Competing interests:** Authors declare no competing interests; **Data and materials availability:** All data is available in the main text or the supplementary materials.

## Supplementary Materials

1. Material and Methods

2. Mortality without testing

3. Mortality: Potential factors to reduce the death toll

4. Total infected individuals and the case fatality rates

5. The impact of COVID-19 in the World

Figs. S1 to S13

Tables S1 to S8

References

## Supplementary Materials for

This PDF file includes:

1. Material and Methods

2. Mortality without testing

3. Mortality: Potential factors to reduce the death toll

4. Total infected individuals and the case fatality rates

5. The impact of COVID-19 in the World

Figs. S1 to S13

Tables S1 to S8

References

## 1 Material and methods

### 1.1 Model

To predict the evolution of the number of deaths caused by the epidemic and highlight the importance of testing we extend a standard epidemiological age-structured SEIR (susceptible-exposed-infected-removed) model by (a) accounting for the age-specific mortality rates, (b) explicitly modeling the mortality rate of the COVID-19 epidemics and (c) introducing isolation periods during the incubation period (E) and the infectious period (I) after testing.

To account for the differential effect of mortality by age, each state is comprised of 95 (Ω) age-groups. We distinguish vectors and matrices from scalars by using bold letters. The dynamics of our extended age-structured SEIR model are as follows:

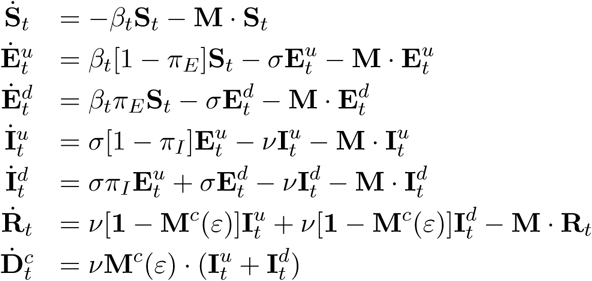

Where 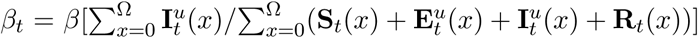 is the infection rate and *β* is the effective transmission rate.^1^ **S***_t_* denotes the vector of susceptible individuals. 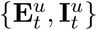 denote the vector of exposed and infectious individuals, respectively, who are undetected. 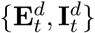 denote the vector of exposed and infectious individuals, respectively, who are detected (tested) and isolated. Note, that similar to (8) we also denote the state 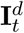 as the infectious detected people and 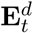 as the detected exposed people (those having been in close contact with infected people and therefore been isolated). **R***_t_* is the vector of recovered individuals and 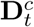 is the vector of deaths from the COVID-19 outbreak. M and **M***^c^*(*ε*) are, respectively, diagonal matrices with age-specific death rates not caused by COVID-19 and the estimated age-specific fatality rates for the COVID-19. The term *ε* in **M***^c^*(*ε*) denotes the share of infectious and symptomatic individuals among all the infectious individuals (i.e. also including the asymptomatic cases). We assume that the (true) fatality rate, **M***^c^*(*ε*), is a fraction *ε* of the (observed) case fatality rate. The (true) fatality rate is defined as the ratio between the total number of deaths from COVID-19 and the total number of infectious individuals, whereas the (observed) case fatality rate is the ratio between the total number of deaths from COVID-19 and the total infected individuals who are detected (tested). Thus, the lower the share of symptomatic individuals the lower the (true) fatality rate compared to the (observed) case fatality rate. 1 denotes the identity matrix. The set of parameters {*β*,*π_E_,π_I_,ν,σ, ε*} denotes the effective transmission rate of the disease, the fraction of people isolated out of the exposed individuals, the fraction of people isolated out of the total infectious individuals, the removal rate, the inverse of the incubation period, and the share of infectious and symptomatic individuals.

**Figure S1:**
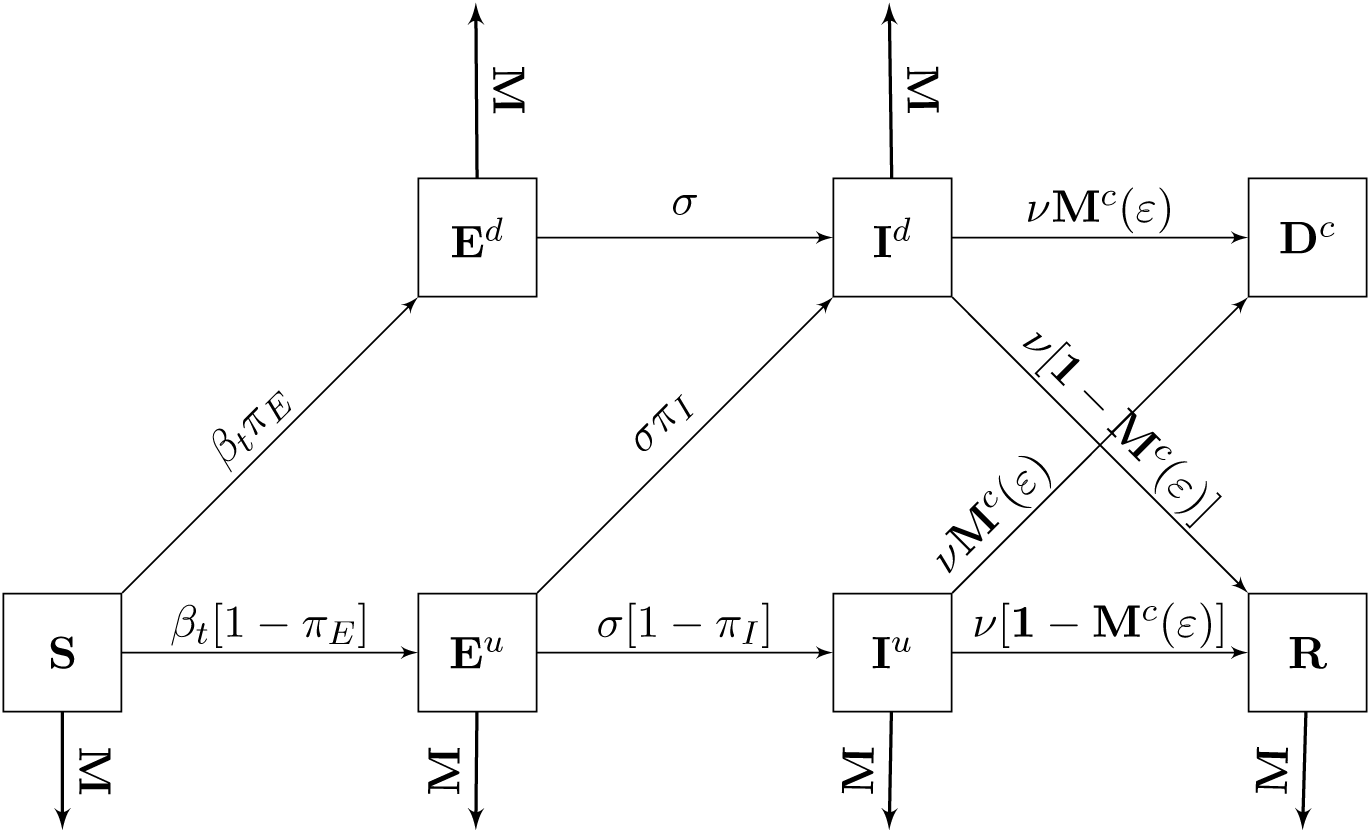
Schematic diagram of the SEIR model with isolation and deaths from COVID-19

The dynamics are presented in the flow diagram Figure S1. In each time step (day) a susceptible individual may become exposed with probability *β_t_*. We assume that a fraction *π_E_* of exposed individuals are isolated. An exposed individual spends an average period of *σ*−^1^ days in incubation until becoming infectious. We assume that a fraction *π_I_* of infected people are isolated. After an average period of ν^−1^ days, infected individuals can either recover with probability 1 − **M***^c^*(*ε*) or die due to COVID-19 with probability **M***^c^*(*ε*).

### 1.2 Calibration

The COVID-19 outbreak is characterized by a large uncertainty on the number of people infected. The number of deaths from COVID-19 seems, however, to be more reliable despite the fact that it can also be subject to under-reporting (6) as well as to over-reporting, due to competing causes of death. Under-reporting is occurring because there is no common agreement across countries on how COVID-19 deaths should be counted. Indeed, many countries are only counting as COVID-19 deaths those individuals who were tested positive, despite the fact that many people who died with COVID-19 symptoms were not tested. To avoid over-reporting, we account in the model for all causes of death, that are not COVID-19, by introducing the matrix of age-specific death rates **M**.

So far, most of the papers on COVID-19 have mainly implemented the infectious disease models using data of reported infected cases and of reported and underreported infections (19). However, given that mortality data presents less uncertainty than data on infections, we propose to calibrate the epidemiological model using the evolution of the number of deaths caused by COVID-19. There are some key features of the mortality pattern shown by COVID-19. First, early statistics on death by COVID-19 have shown a sizable age gradient (3), which is quite similar to the age gradient observed at old age in standard mortality rates. For this reason, it is important to consider in the model that the fatality rate of COVID-19 is increasing with age. To account for the age pattern, we regressed through an OLS the function: log *m_x_* = *γ*_0_ + *γ*_1_*x* + *γ*_2_*x*^2^ + *v* to the log of COVID-19 age-specific fatality rates for ages older than 30 years from (15). We do not fit the fatality rate data below age 30, given that the share of infected and asymptomatic individuals below age 30 is likely to be underreported. The dotted red line in Figure S2 shows the fit of our regression function to the data (red squares) where *γ*_0_ = −10.5063, *γ*_1_ = 0.1310, and *γ*_2_ = −0.0003. Note that the estimated fatality rate (**m***^c^*) fits well the data after age 30.

**Figure S2:**
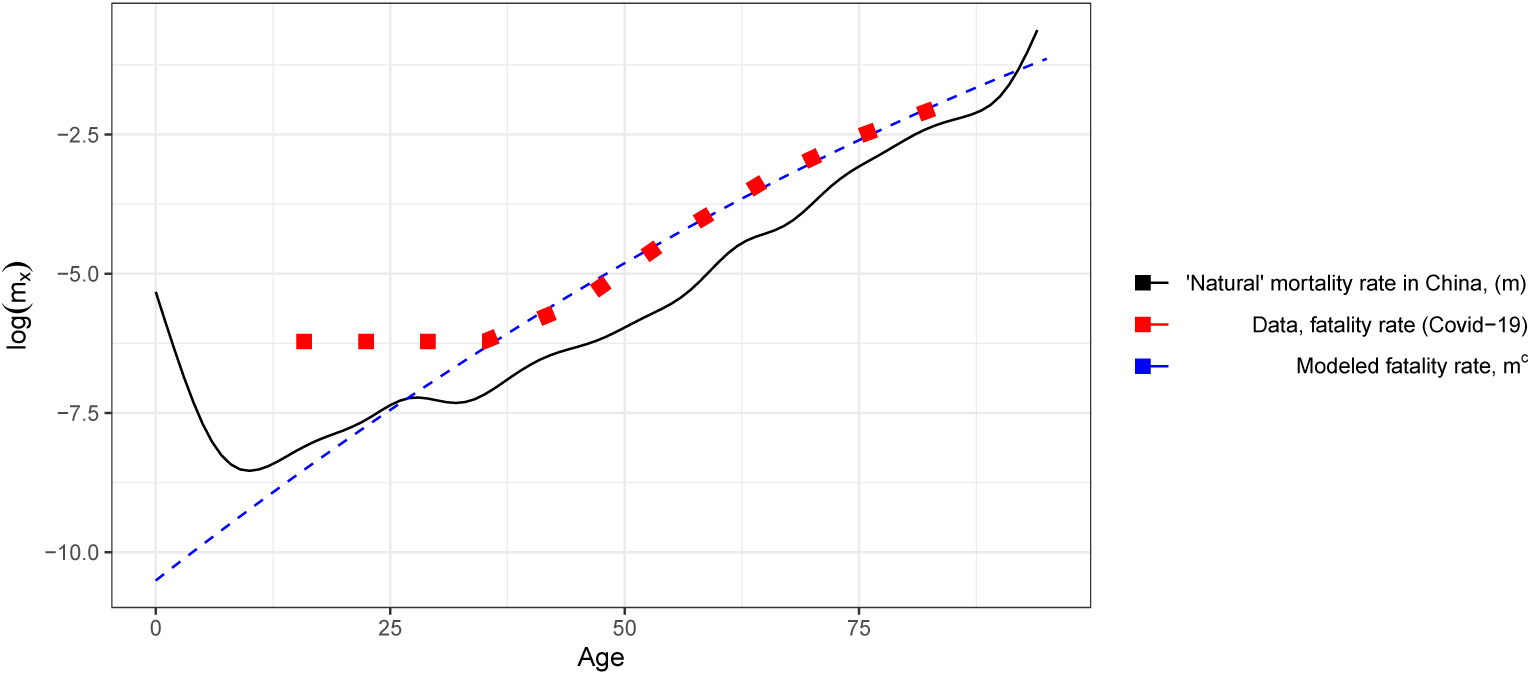
Fitted fatality rate for COVID-19. Source: “Natural” age-specific mortality rates were calculated using data from the UN Population Division. Data on COVID-19 fatality rates by age is taken from (15).

To control for the unknown number of infected people who were not developing symptoms (i.e. asymptomatic) and were not tested against the virus, we have introduced in the calibration process the adjustment factor *ε* ∊ (0,1). Thus, the fatality rate implemented in the SEIR model is **M***^c^*(*ε*) = diag(*ε***m***^c^*) and the death rate excluding COVID-19 is **M** = diag(**m** · *dt*), with *dt* adjusting the annual rates, shown in Fig. S2, to daily rates.

The vector of age-specific mortality rates, **m**, for each country is computed using the estimated deaths *(D_x_)* and exposures *(E_x_)* from (17). Since both deaths and population counts are grouped into 5-year age intervals (with the exception of ages 0–1) we first ungroup the data using a penalized composite link model (14; 11). After ungrouping the data we estimate the observed death rates (*M_x_* = *D_x_/E_x_)* and later, following the procedure adopted by the Human Mortality Database (HMD), we substitute them for smoothed observed death rates from ages 80 to 95 by fitting a Kannisto model. This procedure is to deal with erratic behavior and random fluctuations that are common at older ages (for more details see (16) and the https://www.mortality.org/ for their complete protocol). Thus, assuming that *D_x_* ~ Pois(*E_x_μ_x_*_+0.5_(*a*, *b*)) we estimate the parameters *a* ≥ 0 and *b ≥* 0 by maximizing the log-likelihood function:

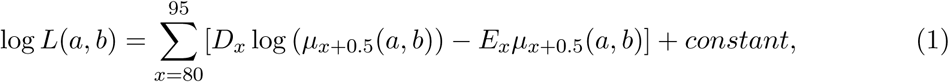

where the function *μ_x_* is given by

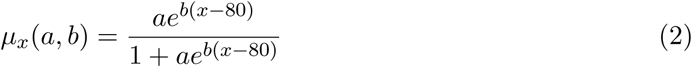

and the constant term accounts for all the functions not dependent on (*a*, *b*). Substituting Eq. (1) into (2) yields the smoothed death rates 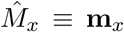. In this way, smoothed rates cannot decline after age 80.^2^

**Bayesian Melding** The aim of our calibration strategy is to fit our SEIR model against the total number of deaths. Since the necessary information to fit the model is likely incomplete, we implement the Bayesian Melding method (12), which provides an inferential framework that takes into account both model’s inputs and outputs. Given that we fit a time-series (evolution of the number of deaths), we implement the Bayesian Melding following (1). The basic purpose with the Bayesian Melding method is to derive the distribution of the set of parameters that best replicate the observed evolution of deaths by using the information from the model and the data.

Let 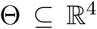 be the set of parameters –inputs– of our SEIR model. For simplicity, we denote by *M*(Θ) the application of our SEIR model given the inputs. Let a realization of Θ be *θ* = *{σ, ν, β*, *ε*}.^3^ Each parameter is considered a random variable with a joint prior distribution *q*_1_(Θ). Given that the epidemiological characteristics of COVID-19 are unknown, we assume an uninformative prior distribution on the inputs.^4^ Let the prior distribution on 0 be

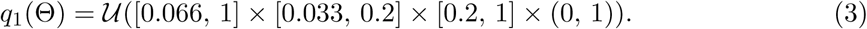

The domains of parameters *σ, ν*, and *β* are taken from the existing literature (13), while we consider any potential value between 0 and 1 for the proportion of symptomatic individuals, i. e. *ε* ∊ (0, 1).

Let Φ be the set of outputs of our SEIR model. Given that *M*(Θ) = Φ, the outputs Φ are also a random variable with a joint prior distribution *q*_2_(Φ). Let a realization of Φ be *ϕ* = {*e*_1_, *e*_2_,…,*e_T_*}, where the output *e_s_* is the difference between the model’s total number of deaths (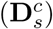) and the total observed deaths from COVID-19 (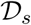) until time s or, the error of fit until time *s*,

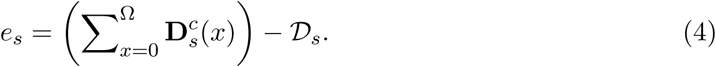

We define time s as the number of days since the first observed death and *T* as the number of days since the first observed death from which the quarantine measures may start slowing the spread of the infection. Since the observed number of deaths until time *T* is subject to under- and over-reporting, we assume the following uninformative joint prior distribution on outputs

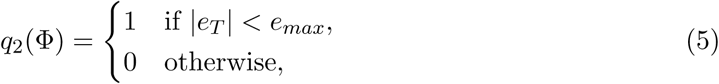

where *e_max_* is the maximum discrepancy of the model with respect to the observed number of deaths until time *T*. Therefore, we assume following (12) that the prior joint distribution of inputs and outputs are independent.

Since our goal is to obtain the joint posterior distribution of Θ, which we denote by *π*^[Θ]^(Θ), we need to update the joint prior distribution of inputs, *q*_1_(Θ), using the observed data through a likelihood function, i.e. 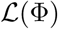. Because *M*(Θ) might not be invertible, we calculate the pooled joint prior distribution on outputs, denoted by *q*^~[Φ]^(Φ), through geometric pooling^5^

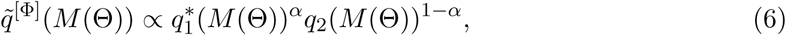

where 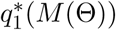 is the induced joint prior distribution of the outputs and *α* is the pooling weight. This is equivalent to finding the region on which both priors have common support (12). A value of *α* close to one (resp. zero) will give a low (resp. high) weight of the information provided by the model on the posterior distribution of inputs. Thus, the Bayesian joint posterior distribution of the outputs is defined as

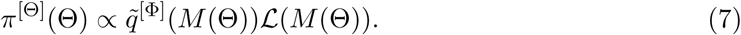

Notice that because the COVID-19 is a new virus we assume no likelihood for the inputs. For the application of the calibration, we assume the error *e_s_* is distributed according to a Normal distribution with *μ* = 0 and 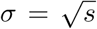, i.e., 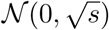.^6^ Thus, we calculate the likelihood of retaining the set of parameters *θ* ∊ Θ as

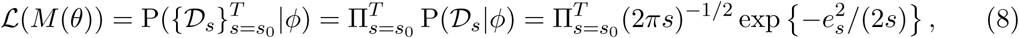

where *s*_0_ is the date at which the total number of deaths is above 50.

Our analysis and calibration were performed with the use of Julia 1.3 (Julia Lab) and R software (R Foundation for Statistical Computing). We calibrate the model to the evolution of the total number of deaths in the province of Hubei (China). We run our model assuming that the first COVID-19 case appeared at November 17th, 2019 (*t* = 1), the first death occurred on January 11th, 2020 (*s* = 1 and *t* = 56), and the epidemic curve started to flatten on February 12, 2020 (*T* = 33 and *t* = 88). The maximum discrepancy *e_max_* is set at 150 deaths in order to allow for sufficient output variability. Moreover, we set the pooling weight, α, at 0.5 in order to give a similar importance to the model and the data. Since the age distribution of the population in Hubei (China) resembles that of China, we scaled down the Chinese population to the total population size of Hubei. However, we keep the death tolls reported from official statistics, since the majority of the cases belong to Hubei. Next, we detail the steps of the Bayesian Melding algorithm:

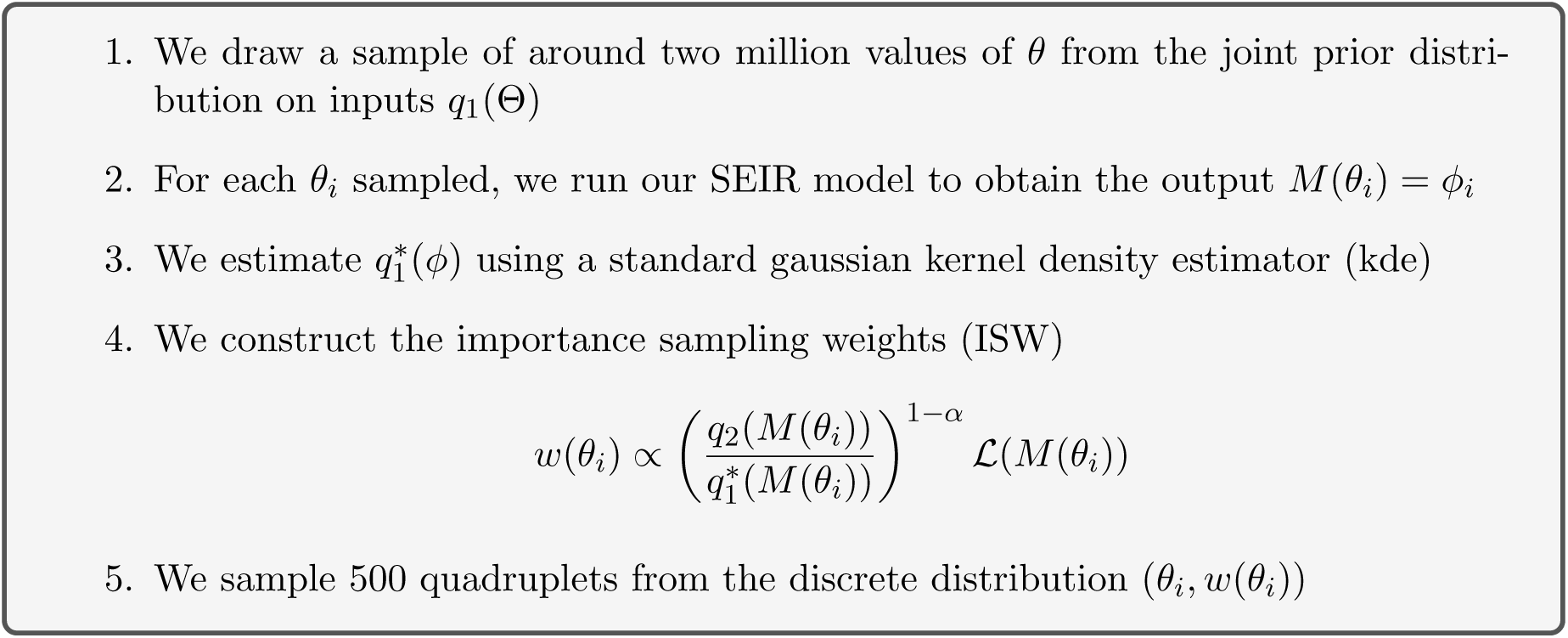

Table S1 summarizes the marginal posterior distributions of the model parameters (Θ).^7^ Parameters *σ*^−1^ and ν^−1^ measure the average durations that a representative individual spends in states **E** and **I**, respectively.^8^ Parameter *β* measures the transmission rate and 1 − *ε* accounts for the proportion of asymptomatic individuals among the infected. We obtain that the mean incubation period is slightly higher than the median incubation of 3.0 days reported for pediatric patients (2) and one day shorter than the average incubation period of 5.2 days reported for patients older than 50 years by (7). Our calibration gives that the recovery period ranges between 6 and 15 days, with an average period of 11 days. By adding the incubation and recovery periods we obtain that the time the virus affects individuals ranges between 7.7 (1st Qu.) and 19 days (3rd Qu.), with an average time of 14.5 days. The transmission rate takes values between 0.325 and 0.489, where the average is 0.432.

**Table S1:** The epidemic parameters of the SEIR model

|  | Min. | 1st Qu. | Median | Mean | 3rd Qu. | Max. |
| --- | --- | --- | --- | --- | --- | --- |
| Incubation period $\sigma^{-1}$ (days) | 1.04 | 1.45 | 1.96 | 3.38 | 4.01 | 13.61 |
| Recovery period $\nu^{-1}$ (days) | 5.12 | 6.31 | 8.51 | 11.07 | 14.98 | 28.79 |
| Transmission rate $\beta$ | 0.258 | 0.325 | 0.401 | 0.432 | 0.489 | 0.977 |
| Fraction of asymptomatic $1 - \varepsilon$ | 0.000 | 0.447 | 0.593 | 0.561 | 0.683 | 0.957 |

Figure S3 shows the posterior distribution of the basic reproduction number (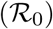). This plot shows how frequent an 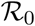 value is obtained across the 500 combinations of the parameters drawn from the discrete distribution (*θ*,*ω*(*θ*)). Our results give that the most probable 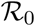 values range between 2.6 (1st Qu.) and 5.1 (3rd. Qu.). The mean value of 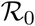, which is represented by a vertical red line in Fig. S3, is 4.51. Thus, the mean 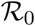 is very close to the basic reproduction number estimated for Wuhan (9), while the most frequently obtained value is between 2.5 and 3.0.

**Figure S3:**
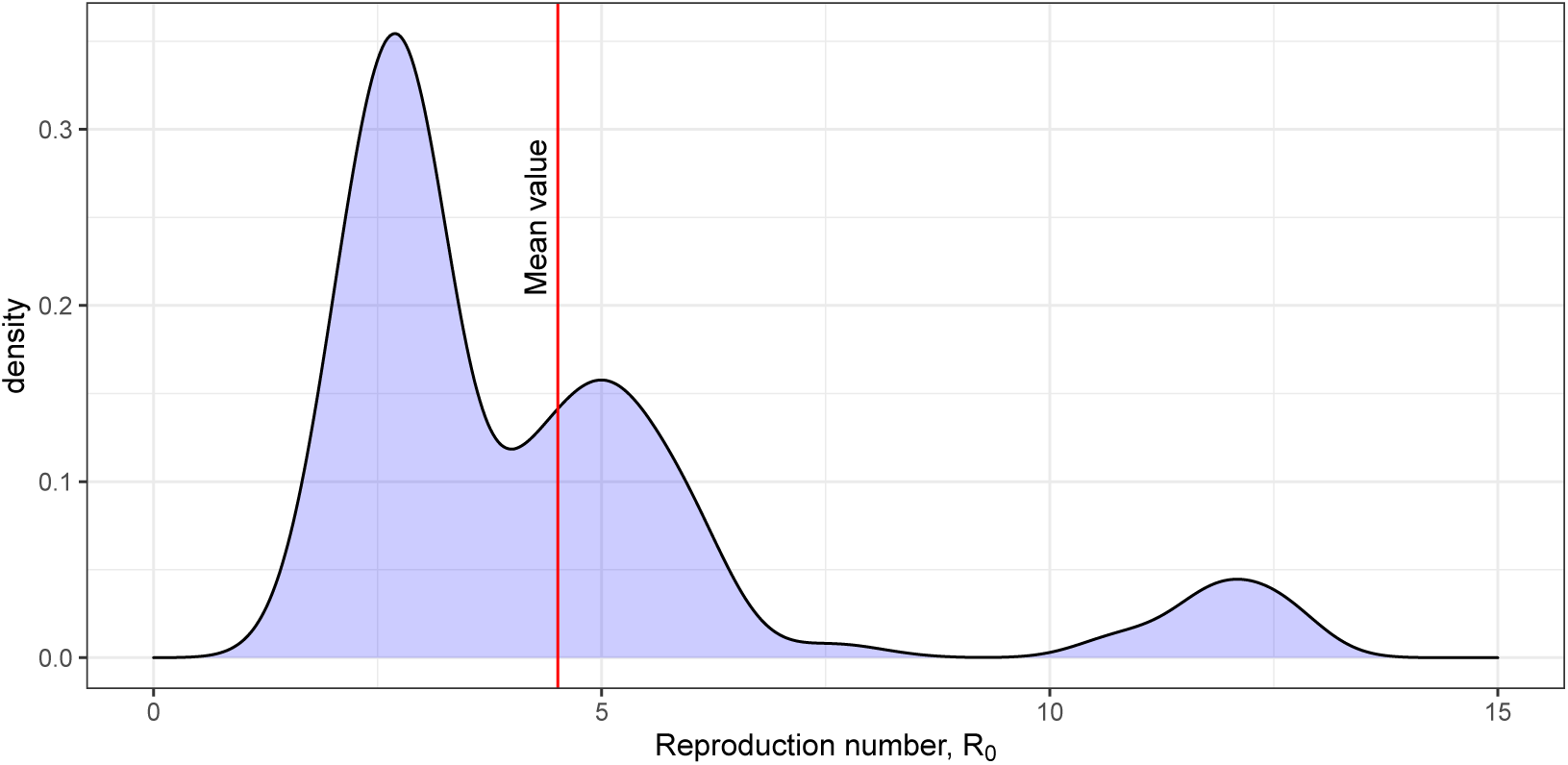
Posterior distribution of the basic reproduction number 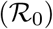.

The last row in Table S1 reports the 1st and 3rd quartiles and the median and mean values for the fraction of asymptomatic individuals, 1 − *ε*. Our calibration suggests that 56 percent of the infected individuals are on average asymptomatic and with 50% probability the fraction of asymptomatic individuals range between 44.65% and 68.31%. Moreover, following (10) and assuming that among the symptomatic individuals roughly 80% are mild, 15% are severe, and 5% are critical, our model suggests that the distribution of the infected individuals according to their symptoms is:

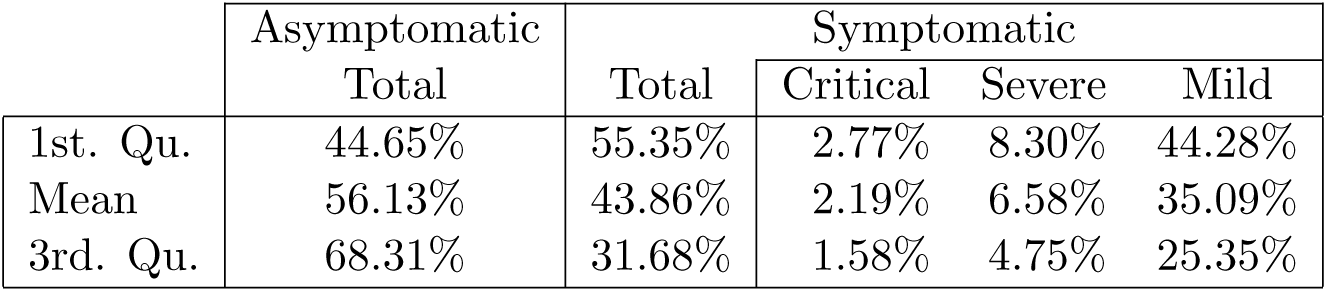

Multiplying the modeled fatality rate **m***^c^* by the fraction of symptomatic people (*ε*), the distribution of the true fatality rate is derived. Figure S4 shows the inferred COVID-19 fatality rate after the calibration process. We obtain that the average fatality rate exceeds 1% at age 60, 5% at age 80, and 10% at age 90, which is sixty percent lower than the fatality rates reported by (18).

## 2 Mortality without testing

To analyze the spread of the virus and the evolution of the total number of deaths, we need an initial number of imported and infected individuals and the date of the onset of COVID-19 outbreak. For comparability reasons, we apply in all countries the same initial number of imported cases and, hence, we use the onset of the outbreak to adjust the epidemic curve to each country. Thus, by assuming similar initial characteristics for all countries, this strategy will allow us to understand, in future work, whether the rapid spread of the virus was due to the number of undetected and infected cases or to the number of days since the first undetected and infected case appeared in the country.

**Figure S4:**
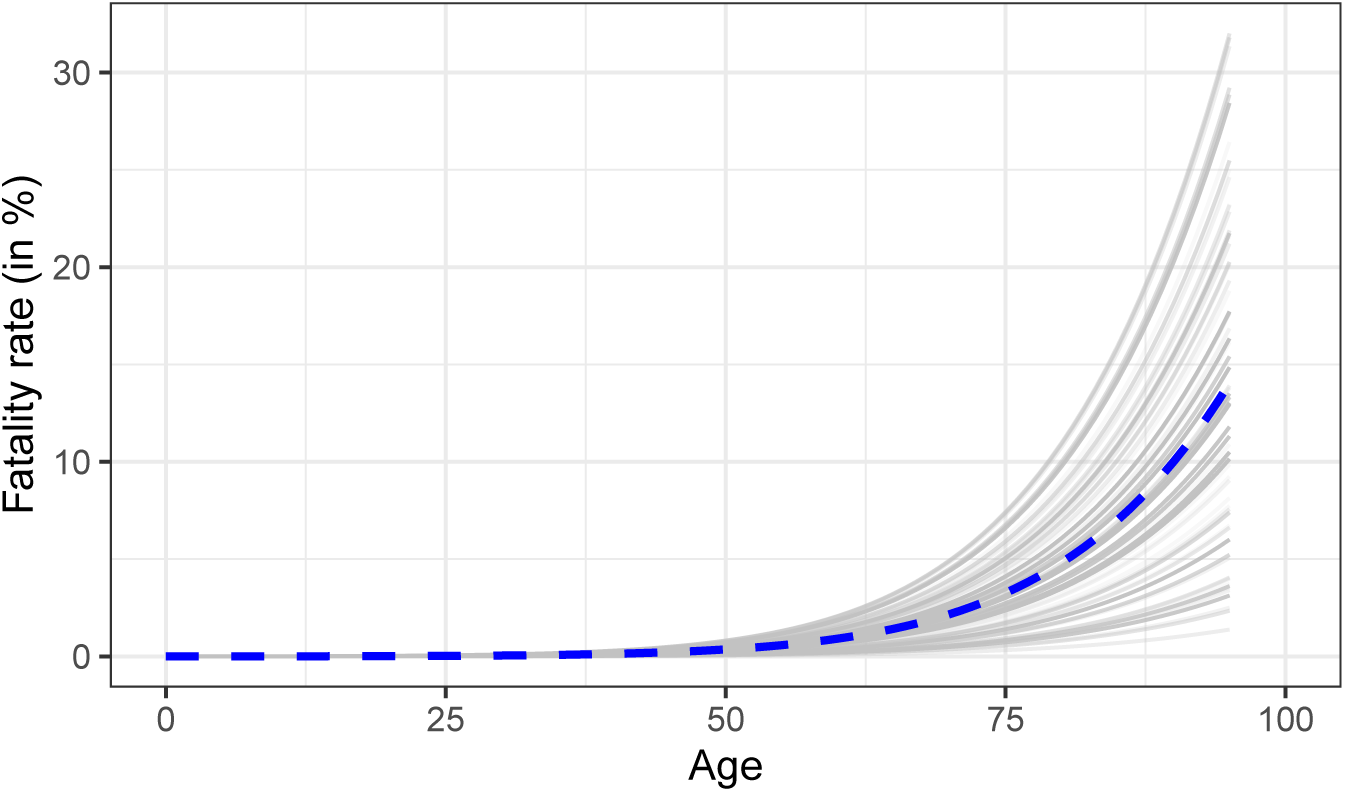
Inferred COVID-19 fatality rate, **M***^c^*(*ε*). *Source:* Authors’ estimates using the Bayesian Melding method. *Notes:* Each gray line depicts the fatality rate resulting from a given set of parameters *θ* drawn from the posterior distribution. The blue dashed line is the average fatality rate across all simulations.

Let us consider that the number of imported and infected individuals follows a temporal Poisson process, Pois(*λt*), with *λ* = 10. This process, which is shown in Figure S5, implies that each month a country receives on average 300 infected individuals and, therefore, an outbreak can occur at any time in the year. Let us also assume that the age of the infected individuals is a random number drawn from a uniform distribution with a minimum age of 18 and a maximum of 65, 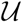(18, 65), which corresponds to the age of potential workers. This age range implies that the average age of the infected individuals is 41.5 years old, which does not necessarily coincide with the average age of the population of each country we consider. Also, these two assumptions yield that the average period to observe the first death after the first infected cases enter in the country is 18 days.

Figure S6 shows that the model is capable of tracking well the evolution of the total number of deaths in eight different countries (United States of America, Italy, Spain, France, United Kingdom, Belgium, Netherlands, Brazil), which have different demographic characteristics. At the end of the period analyzed circles deviate from the model prediction because of the implementation of lockdown measures. These countries are chosen because they had more than 300 deaths by April 3rd, 2020 —the date at which the first data was collected— and because their case fatality rate was greater than 2.5%.^9^ These two assumptions imply that neither massive testing nor isolation measure were initially implemented. In Fig. S6 the average estimate of the model is depicted with a dashed blue line, data is represented with black circles, and gray lines correspond to each simulation drawn from the posterior distribution, (*θ*, *w*(*θ*)), obtained with the Bayesian Melding method. Countries are distributed according to the total number of deaths. To calculate the number of days since the onset of COVID-19 outbreak, we modify the date in which the first infected case is reported. In particular, we assume that in the Netherlands the first imported cases occurred 22 days before the first infected case was detected, in Brazil 15 days, in Italy 13 days, in Spain 9 days, in United Kingdom 0 days, in France -3 days, and in Belgium and US -4 days. A negative number implies that the assumed flow of imported cases (see Fig. S5) is higher than what the actual data shows.

**Figure S5:**
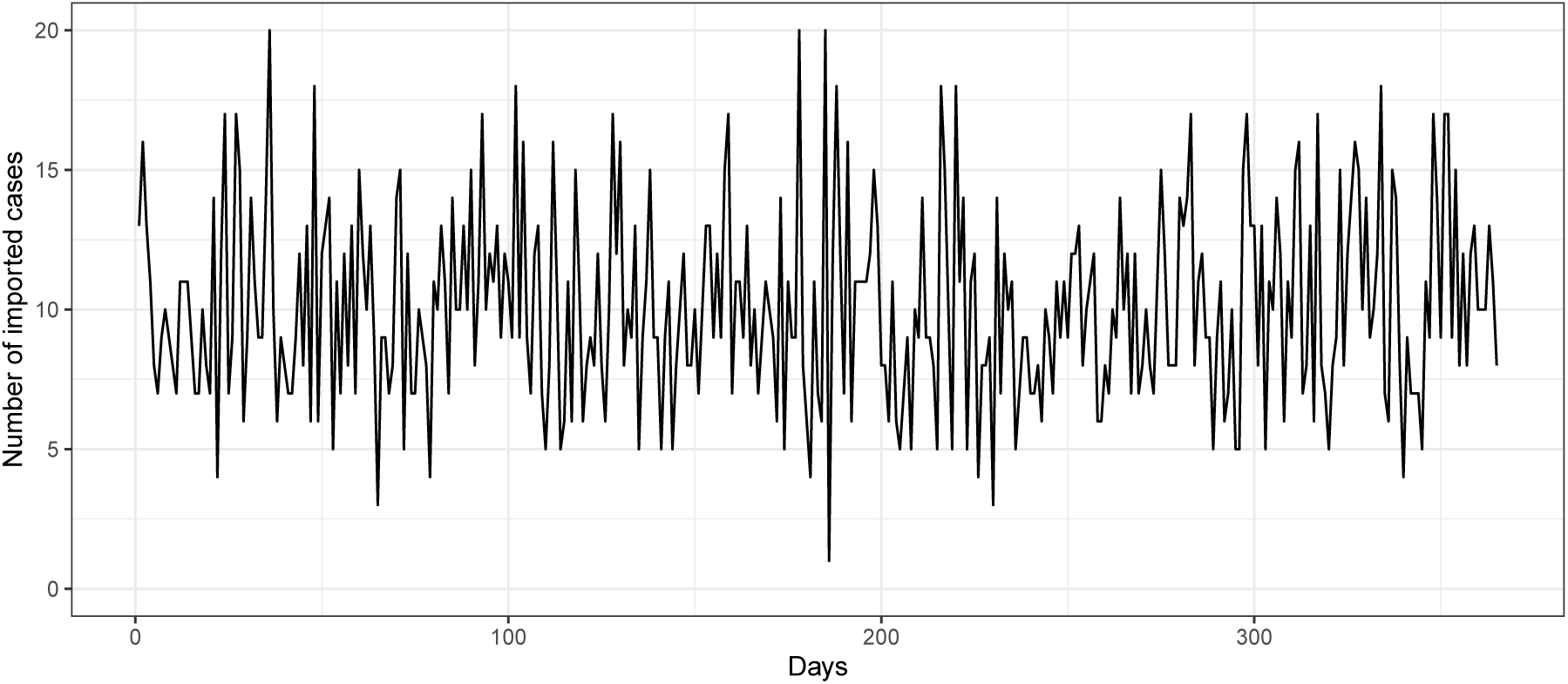
Number of imported cases used in all countries

To investigate the factors explaining the potential difference in fatality rates across countries, we assume that there is no isolation measures and the population is well mixed. Under these two assumptions we obtain that one key factor explaining differences in the fatality rate across countries is the population size. To show the extreme cases, we show in Figure S7 how the transmission rate is faster in countries with a small population size like Iceland (over 340 thousand inhabitants) compared to countries with a large population size like Brazil (over 210 million inhabitants) and China (over 1400 million inhabitants). This implies that whenever the above mentioned assumptions are satisfied, small populations will get infected faster than large population. As a consequence, the peak of the mortality rate is reached sooner in small populations than in large populations.

Another key factor explaining differences in fatality rates across countries is the age distribution of the population. Assuming no isolation measures, Figure S8 shows the positive relationship between the average COVID-19 fatality rate and the mean age of the population for two hundred countries after one year since the onset of the outbreak. Gray dots depict the death rate (excluding COVID-19 deaths) in each country according to the mean age of the population. We can see in Fig. S8 that younger populations -mean age younger than 30 years-face a fatality rate around 0.2%, whereas older populations -mean age older than 40 years-face a fatality rate close to 0.9%. By comparing the average fatality rates to the death rates (gray dots) we can observe that COVID-19 is not the main caused of death in younger populations. In contrast, since the average fatality rates and the deaths rates are almost similar in aging countries, COVID-19 has the potential to double the number of deaths in aging populations, unless policies to contain the spread of the virus are implemented. All the data used for plotting Fig. S8 is reported in Table S2.^10^

**Figure S6:**
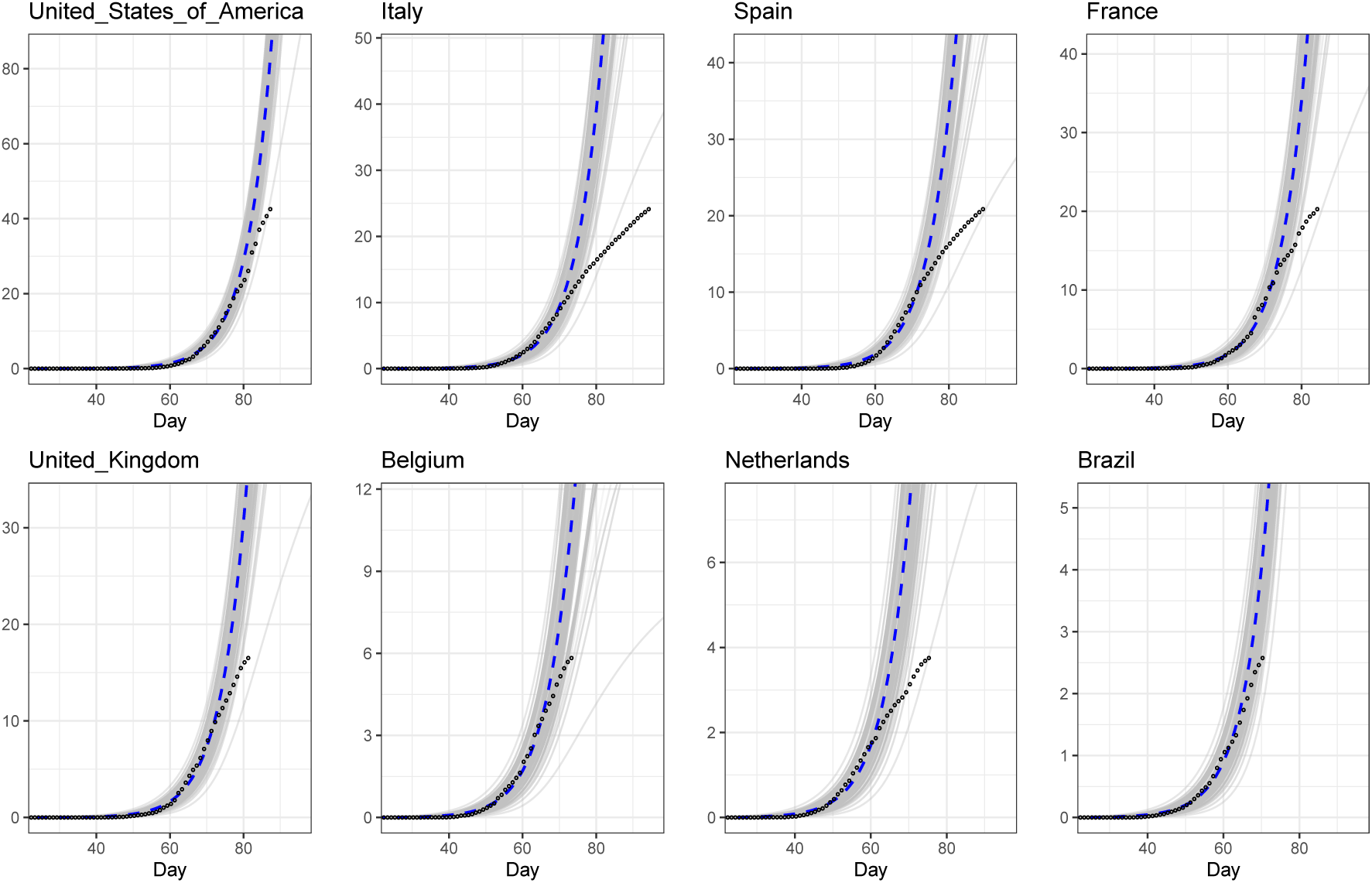
Estimated COVID-19 death toll (in 1 000s) for selected countries without testing and isolation. Source: Data taken from the European Centre for Disease Prevention and Control (ECDC) collected on April 21st, 2020. Note: Black circles depict the data. In all countries the data deviates from the model prediction due to the later implementation of lockdown measures.

**Figure S7:**
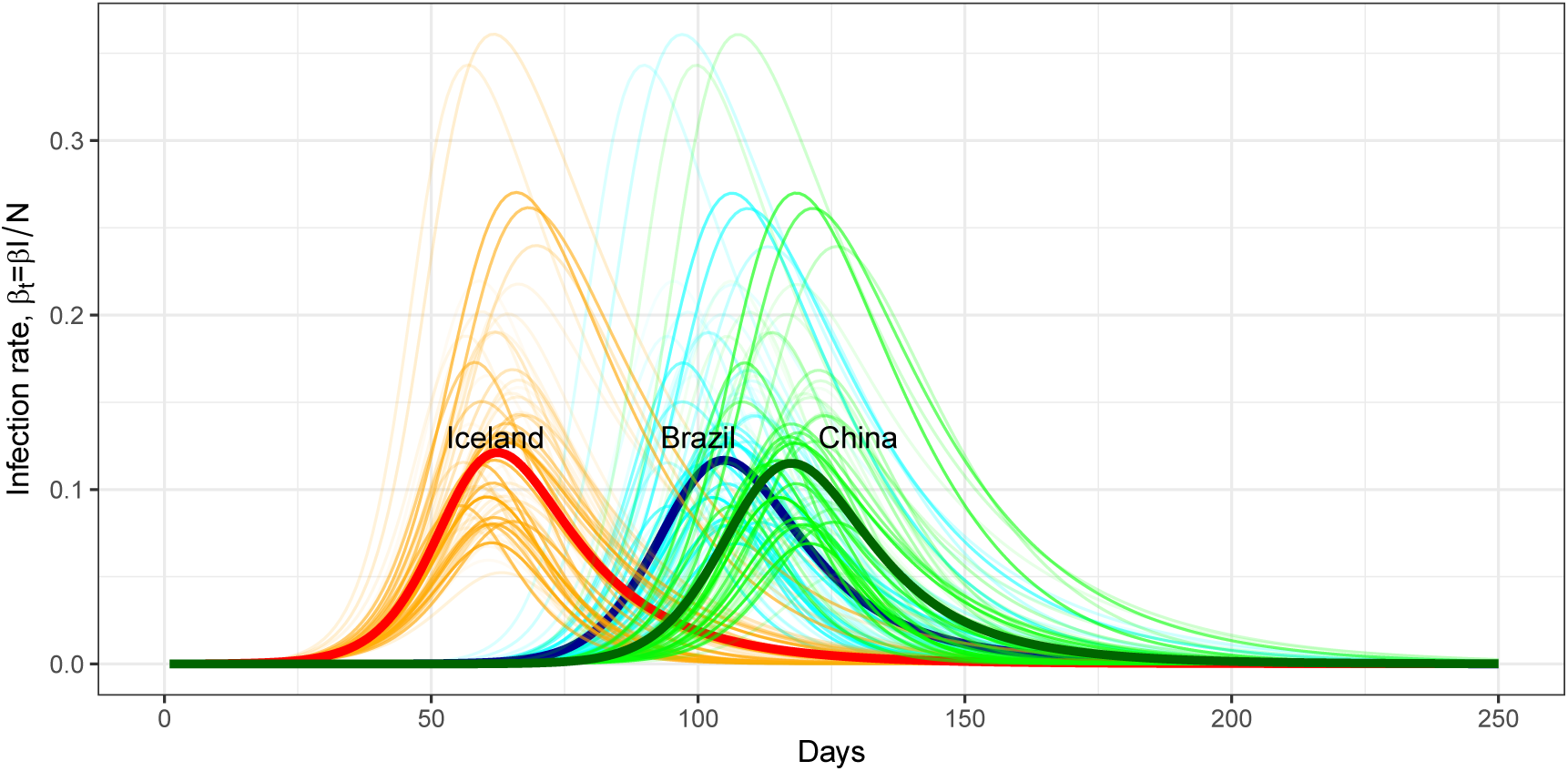
COVID-19 infection rate in Iceland (red), Brazil (blue), and China (green). *Source:* Authors’ calculations. *Note:* Infectious rate calculated under two assumptions: a) no isolation measures and b) the population is well mixed.

**Figure S8:**
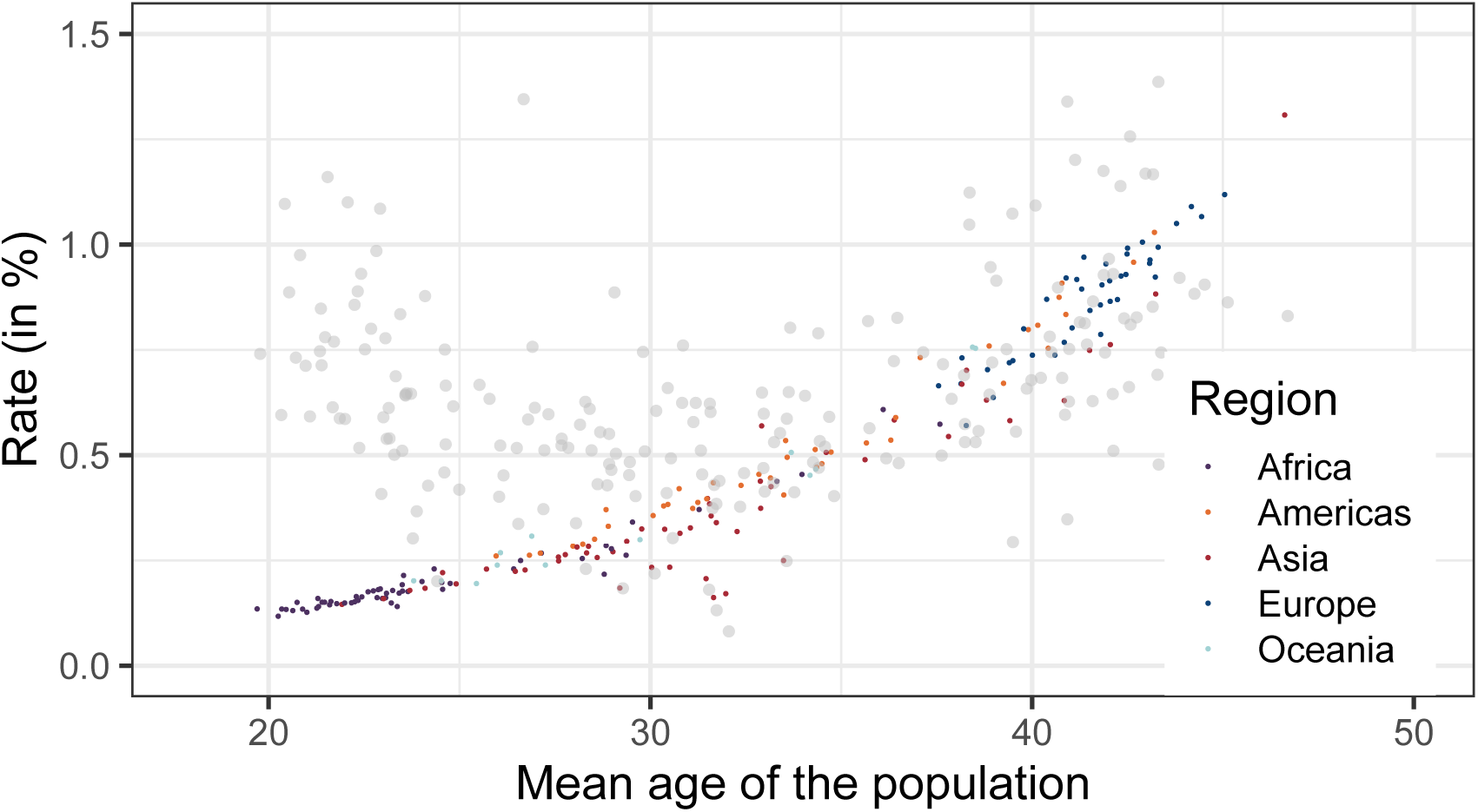
Relationship between the average COVID-19 fatality rate and the mean age of the population after 365 days. *Source:* UN Population data and authors’ calculations. *Note:* Gray dots depict the death rate (without COVID-19) in each country according to the mean age of the population. The data used in this plot is reported in Table S2.

**Table S2:**
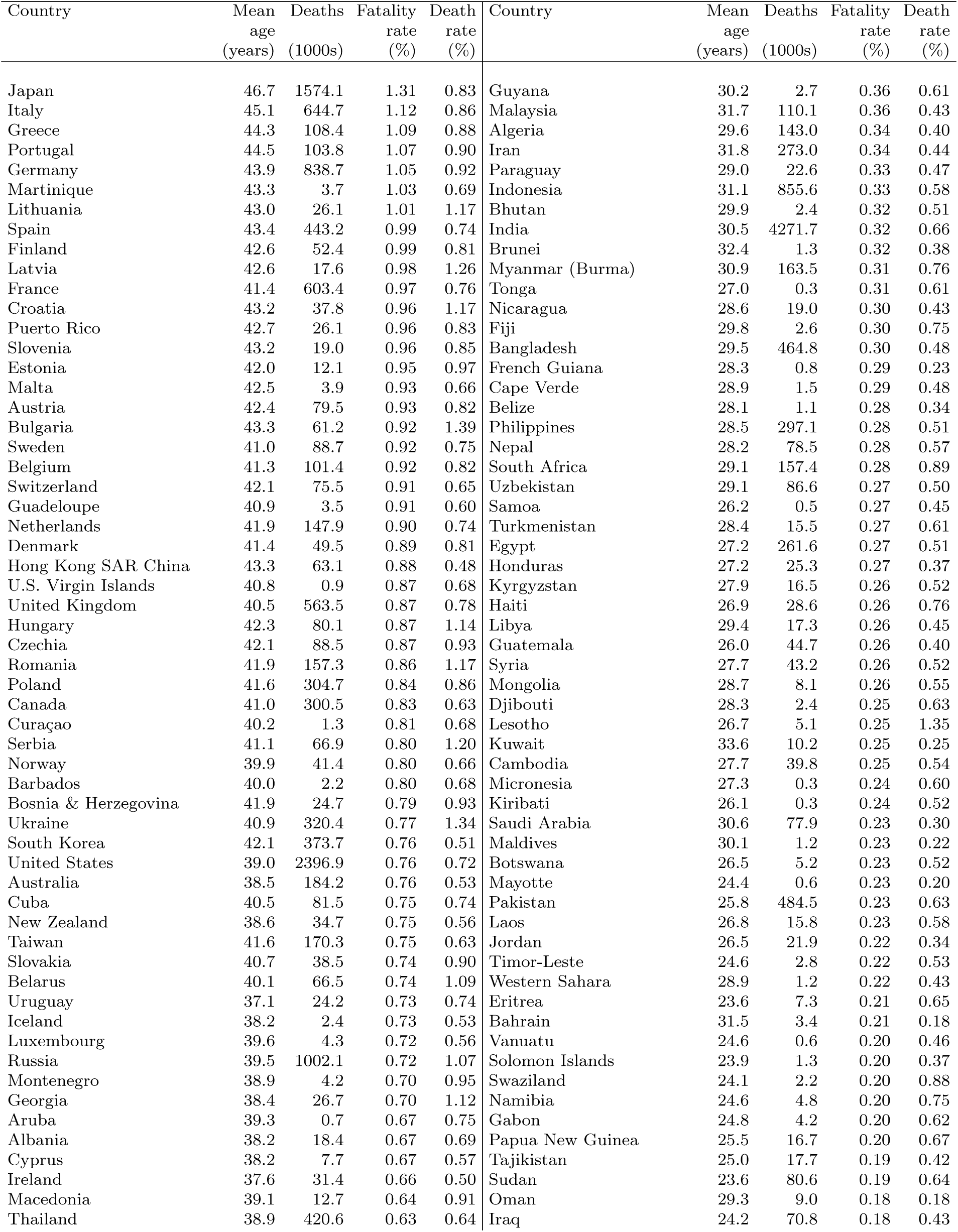
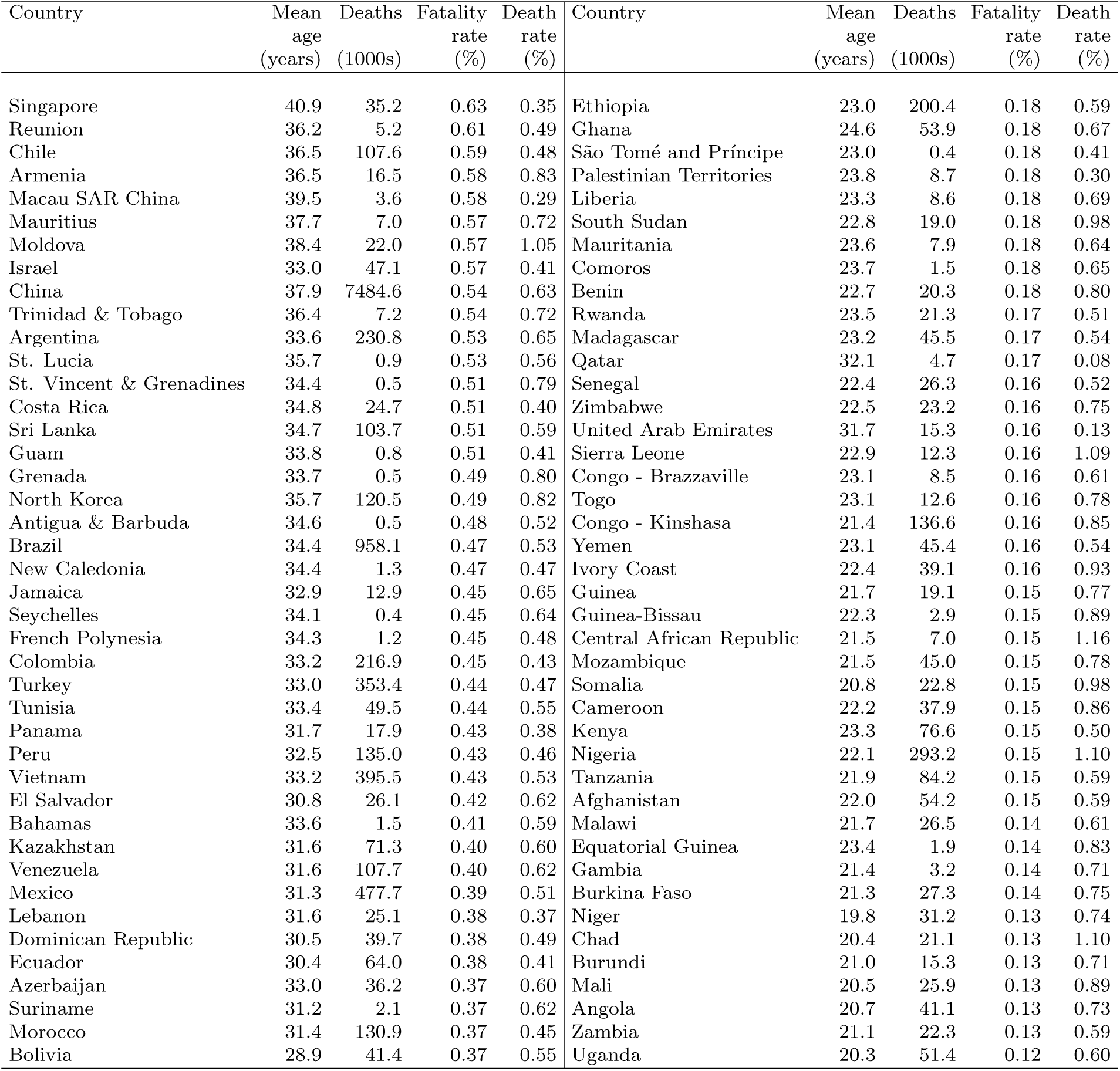
(Average) death toll of COVID-19 after 365 days in the World: Ordered according to the fatality rate.

| Country | Mean<br>age<br>(years) | Deaths<br>(1000s) | Fatality<br>rate<br>(%) | Death<br>rate<br>(%) | Country | Mean<br>age<br>(years) | Deaths<br>(1000s) | Fatality<br>rate<br>(%) | Death<br>rate<br>(%) |
| --- | --- | --- | --- | --- | --- | --- | --- | --- | --- |
| Japan | 46.7 | 1574.1 | 1.31 | 0.83 | Guyana | 30.2 | 2.7 | 0.36 | 0.61 |
| Italy | 45.1 | 644.7 | 1.12 | 0.86 | Malaysia | 31.7 | 110.1 | 0.36 | 0.43 |
| Greece | 44.3 | 108.4 | 1.09 | 0.88 | Algeria | 29.6 | 143.0 | 0.34 | 0.40 |
| Portugal | 44.5 | 103.8 | 1.07 | 0.90 | Iran | 31.8 | 273.0 | 0.34 | 0.44 |
| Germany | 43.9 | 838.7 | 1.05 | 0.92 | Paraguay | 29.0 | 22.6 | 0.33 | 0.47 |
| Martinique | 43.3 | 3.7 | 1.03 | 0.69 | Indonesia | 31.1 | 855.6 | 0.33 | 0.58 |
| Lithuania | 43.0 | 26.1 | 1.01 | 1.17 | Bhutan | 29.9 | 2.4 | 0.32 | 0.51 |
| Spain | 43.4 | 443.2 | 0.99 | 0.74 | India | 30.5 | 4271.7 | 0.32 | 0.66 |
| Finland | 42.6 | 52.4 | 0.99 | 0.81 | Brunei | 32.4 | 1.3 | 0.32 | 0.38 |
| Latvia | 42.6 | 17.6 | 0.98 | 1.26 | Myanmar (Burma) | 30.9 | 163.5 | 0.31 | 0.76 |
| France | 41.4 | 603.4 | 0.97 | 0.76 | Tonga | 27.0 | 0.3 | 0.31 | 0.61 |
| Croatia | 43.2 | 37.8 | 0.96 | 1.17 | Nicaragua | 28.6 | 19.0 | 0.30 | 0.43 |
| Puerto Rico | 42.7 | 26.1 | 0.96 | 0.83 | Fiji | 29.8 | 2.6 | 0.30 | 0.75 |
| Slovenia | 43.2 | 19.0 | 0.96 | 0.85 | Bangladesh | 29.5 | 464.8 | 0.30 | 0.48 |
| Estonia | 42.0 | 12.1 | 0.95 | 0.97 | French Guiana | 28.3 | 0.8 | 0.29 | 0.23 |
| Malta | 42.5 | 3.9 | 0.93 | 0.66 | Cape Verde | 28.9 | 1.5 | 0.29 | 0.48 |
| Austria | 42.4 | 79.5 | 0.93 | 0.82 | Belize | 28.1 | 1.1 | 0.28 | 0.34 |
| Bulgaria | 43.3 | 61.2 | 0.92 | 1.39 | Philippines | 28.5 | 297.1 | 0.28 | 0.51 |
| Sweden | 41.0 | 88.7 | 0.92 | 0.75 | Nepal | 28.2 | 78.5 | 0.28 | 0.57 |
| Belgium | 41.3 | 101.4 | 0.92 | 0.82 | South Africa | 29.1 | 157.4 | 0.28 | 0.89 |
| Switzerland | 42.1 | 75.5 | 0.91 | 0.65 | Uzbekistan | 29.1 | 86.6 | 0.27 | 0.50 |
| Guadeloupe | 40.9 | 3.5 | 0.91 | 0.60 | Samoa | 26.2 | 0.5 | 0.27 | 0.45 |
| Netherlands | 41.9 | 147.9 | 0.90 | 0.74 | Turkmenistan | 28.4 | 15.5 | 0.27 | 0.61 |
| Denmark | 41.4 | 49.5 | 0.89 | 0.81 | Egypt | 27.2 | 261.6 | 0.27 | 0.51 |
| Hong Kong SAR China | 43.3 | 63.1 | 0.88 | 0.48 | Honduras | 27.2 | 25.3 | 0.27 | 0.37 |
| U.S. Virgin Islands | 40.8 | 0.9 | 0.87 | 0.68 | Kyrgyzstan | 27.9 | 16.5 | 0.26 | 0.52 |
| United Kingdom | 40.5 | 563.5 | 0.87 | 0.78 | Haiti | 26.9 | 28.6 | 0.26 | 0.76 |
| Hungary | 42.3 | 80.1 | 0.87 | 1.14 | Libya | 29.4 | 17.3 | 0.26 | 0.45 |
| Czechia | 42.1 | 88.5 | 0.87 | 0.93 | Guatemala | 26.0 | 44.7 | 0.26 | 0.40 |
| Romania | 41.9 | 157.3 | 0.86 | 1.17 | Syria | 27.7 | 43.2 | 0.26 | 0.52 |
| Poland | 41.6 | 304.7 | 0.84 | 0.86 | Mongolia | 28.7 | 8.1 | 0.26 | 0.55 |
| Canada | 41.0 | 300.5 | 0.83 | 0.63 | Djibouti | 28.3 | 2.4 | 0.25 | 0.63 |
| Curaçao | 40.2 | 1.3 | 0.81 | 0.68 | Lesotho | 26.7 | 5.1 | 0.25 | 1.35 |
| Serbia | 41.1 | 66.9 | 0.80 | 1.20 | Kuwait | 33.6 | 10.2 | 0.25 | 0.25 |
| Norway | 39.9 | 41.4 | 0.80 | 0.66 | Cambodia | 27.7 | 39.8 | 0.25 | 0.54 |
| Barbados | 40.0 | 2.2 | 0.80 | 0.68 | Micronesia | 27.3 | 0.3 | 0.24 | 0.60 |
| Bosnia & Herzegovina | 41.9 | 24.7 | 0.79 | 0.93 | Kiribati | 26.1 | 0.3 | 0.24 | 0.52 |
| Ukraine | 40.9 | 320.4 | 0.77 | 1.34 | Saudi Arabia | 30.6 | 77.9 | 0.23 | 0.30 |
| South Korea | 42.1 | 373.7 | 0.76 | 0.51 | Maldives | 30.1 | 1.2 | 0.23 | 0.22 |
| United States | 39.0 | 2396.9 | 0.76 | 0.72 | Botswana | 26.5 | 5.2 | 0.23 | 0.52 |
| Australia | 38.5 | 184.2 | 0.76 | 0.53 | Mayotte | 24.4 | 0.6 | 0.23 | 0.20 |
| Cuba | 40.5 | 81.5 | 0.75 | 0.74 | Pakistan | 25.8 | 484.5 | 0.23 | 0.63 |
| New Zealand | 38.6 | 34.7 | 0.75 | 0.56 | Laos | 26.8 | 15.8 | 0.23 | 0.58 |
| Taiwan | 41.6 | 170.3 | 0.75 | 0.63 | Jordan | 26.5 | 21.9 | 0.22 | 0.34 |
| Slovakia | 40.7 | 38.5 | 0.74 | 0.90 | Timor-Leste | 24.6 | 2.8 | 0.22 | 0.53 |
| Belarus | 40.1 | 66.5 | 0.74 | 1.09 | Western Sahara | 28.9 | 1.2 | 0.22 | 0.43 |
| Uruguay | 37.1 | 24.2 | 0.73 | 0.74 | Eritrea | 23.6 | 7.3 | 0.21 | 0.65 |
| Iceland | 38.2 | 2.4 | 0.73 | 0.53 | Bahrain | 31.5 | 3.4 | 0.21 | 0.18 |
| Luxembourg | 39.6 | 4.3 | 0.72 | 0.56 | Vanuatu | 24.6 | 0.6 | 0.20 | 0.46 |
| Russia | 39.5 | 1002.1 | 0.72 | 1.07 | Solomon Islands | 23.9 | 1.3 | 0.20 | 0.37 |
| Montenegro | 38.9 | 4.2 | 0.70 | 0.95 | Swaziland | 24.1 | 2.2 | 0.20 | 0.88 |
| Georgia | 38.4 | 26.7 | 0.70 | 1.12 | Namibia | 24.6 | 4.8 | 0.20 | 0.75 |
| Aruba | 39.3 | 0.7 | 0.67 | 0.75 | Gabon | 24.8 | 4.2 | 0.20 | 0.62 |
| Albania | 38.2 | 18.4 | 0.67 | 0.69 | Papua New Guinea | 25.5 | 16.7 | 0.20 | 0.67 |
| Cyprus | 38.2 | 7.7 | 0.67 | 0.57 | Tajikistan | 25.0 | 17.7 | 0.19 | 0.42 |
| Ireland | 37.6 | 31.4 | 0.66 | 0.50 | Sudan | 23.6 | 80.6 | 0.19 | 0.64 |
| Macedonia | 39.1 | 12.7 | 0.64 | 0.91 | Oman | 29.3 | 9.0 | 0.18 | 0.18 |
| Thailand | 38.9 | 420.6 | 0.63 | 0.64 | Iraq | 24.2 | 70.8 | 0.18 | 0.43 |
| Singapore | 40.9 | 35.2 | 0.63 | 0.35 | Ethiopia | 23.0 | 200.4 | 0.18 | 0.59 |
| Reunion | 36.2 | 5.2 | 0.61 | 0.49 | Ghana | 24.6 | 53.9 | 0.18 | 0.67 |
| Chile | 36.5 | 107.6 | 0.59 | 0.48 | São Tomé and Príncipe | 23.0 | 0.4 | 0.18 | 0.41 |
| Armenia | 36.5 | 16.5 | 0.58 | 0.83 | Palestinian Territories | 23.8 | 8.7 | 0.18 | 0.30 |
| Macau SAR China | 39.5 | 3.6 | 0.58 | 0.29 | Liberia | 23.3 | 8.6 | 0.18 | 0.69 |
| Mauritius | 37.7 | 7.0 | 0.57 | 0.72 | South Sudan | 22.8 | 19.0 | 0.18 | 0.98 |
| Moldova | 38.4 | 22.0 | 0.57 | 1.05 | Mauritania | 23.6 | 7.9 | 0.18 | 0.64 |
| Israel | 33.0 | 47.1 | 0.57 | 0.41 | Comoros | 23.7 | 1.5 | 0.18 | 0.65 |
| China | 37.9 | 7484.6 | 0.54 | 0.63 | Benin | 22.7 | 20.3 | 0.18 | 0.80 |
| Trinidad & Tobago | 36.4 | 7.2 | 0.54 | 0.72 | Rwanda | 23.5 | 21.3 | 0.17 | 0.51 |
| Argentina | 33.6 | 230.8 | 0.53 | 0.65 | Madagascar | 23.2 | 45.5 | 0.17 | 0.54 |
| St. Lucia | 35.7 | 0.9 | 0.53 | 0.56 | Qatar | 32.1 | 4.7 | 0.17 | 0.08 |
| St. Vincent & Grenadines | 34.4 | 0.5 | 0.51 | 0.79 | Senegal | 22.4 | 26.3 | 0.16 | 0.52 |
| Costa Rica | 34.8 | 24.7 | 0.51 | 0.40 | Zimbabwe | 22.5 | 23.2 | 0.16 | 0.75 |
| Sri Lanka | 34.7 | 103.7 | 0.51 | 0.59 | United Arab Emirates | 31.7 | 15.3 | 0.16 | 0.13 |
| Guam | 33.8 | 0.8 | 0.51 | 0.41 | Sierra Leone | 22.9 | 12.3 | 0.16 | 1.09 |
| Grenada | 33.7 | 0.5 | 0.49 | 0.80 | Congo - Brazzaville | 23.1 | 8.5 | 0.16 | 0.61 |
| North Korea | 35.7 | 120.5 | 0.49 | 0.82 | Togo | 23.1 | 12.6 | 0.16 | 0.78 |
| Antigua & Barbuda | 34.6 | 0.5 | 0.48 | 0.52 | Congo - Kinshasa | 21.4 | 136.6 | 0.16 | 0.85 |
| Brazil | 34.4 | 958.1 | 0.47 | 0.53 | Yemen | 23.1 | 45.4 | 0.16 | 0.54 |
| New Caledonia | 34.4 | 1.3 | 0.47 | 0.47 | Ivory Coast | 22.4 | 39.1 | 0.16 | 0.93 |
| Jamaica | 32.9 | 12.9 | 0.45 | 0.65 | Guinea | 21.7 | 19.1 | 0.15 | 0.77 |
| Seychelles | 34.1 | 0.4 | 0.45 | 0.64 | Guinea-Bissau | 22.3 | 2.9 | 0.15 | 0.89 |
| French Polynesia | 34.3 | 1.2 | 0.45 | 0.48 | Central African Republic | 21.5 | 7.0 | 0.15 | 1.16 |
| Colombia | 33.2 | 216.9 | 0.45 | 0.43 | Mozambique | 21.5 | 45.0 | 0.15 | 0.78 |
| Turkey | 33.0 | 353.4 | 0.44 | 0.47 | Somalia | 20.8 | 22.8 | 0.15 | 0.98 |
| Tunisia | 33.4 | 49.5 | 0.44 | 0.55 | Cameroon | 22.2 | 37.9 | 0.15 | 0.86 |
| Panama | 31.7 | 17.9 | 0.43 | 0.38 | Kenya | 23.3 | 76.6 | 0.15 | 0.50 |
| Peru | 32.5 | 135.0 | 0.43 | 0.46 | Nigeria | 22.1 | 293.2 | 0.15 | 1.10 |
| Vietnam | 33.2 | 395.5 | 0.43 | 0.53 | Tanzania | 21.9 | 84.2 | 0.15 | 0.59 |
| El Salvador | 30.8 | 26.1 | 0.42 | 0.62 | Afghanistan | 22.0 | 54.2 | 0.15 | 0.59 |
| Bahamas | 33.6 | 1.5 | 0.41 | 0.59 | Malawi | 21.7 | 26.5 | 0.14 | 0.61 |
| Kazakhstan | 31.6 | 71.3 | 0.40 | 0.60 | Equatorial Guinea | 23.4 | 1.9 | 0.14 | 0.83 |
| Venezuela | 31.6 | 107.7 | 0.40 | 0.62 | Gambia | 21.4 | 3.2 | 0.14 | 0.71 |
| Mexico | 31.3 | 477.7 | 0.39 | 0.51 | Burkina Faso | 21.3 | 27.3 | 0.14 | 0.75 |
| Lebanon | 31.6 | 25.1 | 0.38 | 0.37 | Niger | 19.8 | 31.2 | 0.13 | 0.74 |
| Dominican Republic | 30.5 | 39.7 | 0.38 | 0.49 | Chad | 20.4 | 21.1 | 0.13 | 1.10 |
| Ecuador | 30.4 | 64.0 | 0.38 | 0.41 | Burundi | 21.0 | 15.3 | 0.13 | 0.71 |
| Azerbaijan | 33.0 | 36.2 | 0.37 | 0.60 | Mali | 20.5 | 25.9 | 0.13 | 0.89 |
| Suriname | 31.2 | 2.1 | 0.37 | 0.62 | Angola | 20.7 | 41.1 | 0.13 | 0.73 |
| Morocco | 31.4 | 130.9 | 0.37 | 0.45 | Zambia | 21.1 | 22.3 | 0.13 | 0.59 |
| Bolivia | 28.9 | 41.4 | 0.37 | 0.55 | Uganda | 20.3 | 51.4 | 0.12 | 0.60 |

## 3 Mortality: Potential factors to reduce the death toll

### 3.1 The role of testing and isolation

In the previous section we have shown the potential death toll of COVID-19 assuming that (a) no lockdown policies are implemented and (b) the population is well mixed. In this section, we show that testing and isolation may help to control the outbreak and give estimates on how many lives can be saved. Let *ε* be the fraction of infectious people who develop some symptoms and 1 − *ε* those infectious individuals who are asymptomatic. Following (10) we assume that among the fraction *ε* of infectious individuals, 80% develop mild symptoms, while 20% develop critical and severe symptoms. Moreover, we consider that governments do not start isolating individuals until time *t_I_*. From time *t_I_* onwards, we consider that governments may commit to find those individuals who are infected. Let the proportion of asymptomatic and positively tested people who become isolated be *τ*_1_. Let the proportion of symptomatic and positively tested people who become isolated be *τ*_2_. Thus, we define the fraction of people who are exposed and isolated as

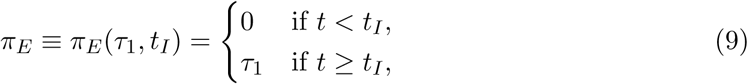

while we define the fraction of people infected who become isolated as

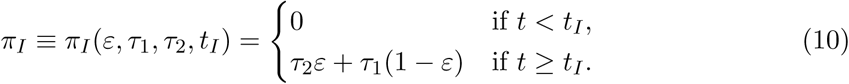

**Table S3:**
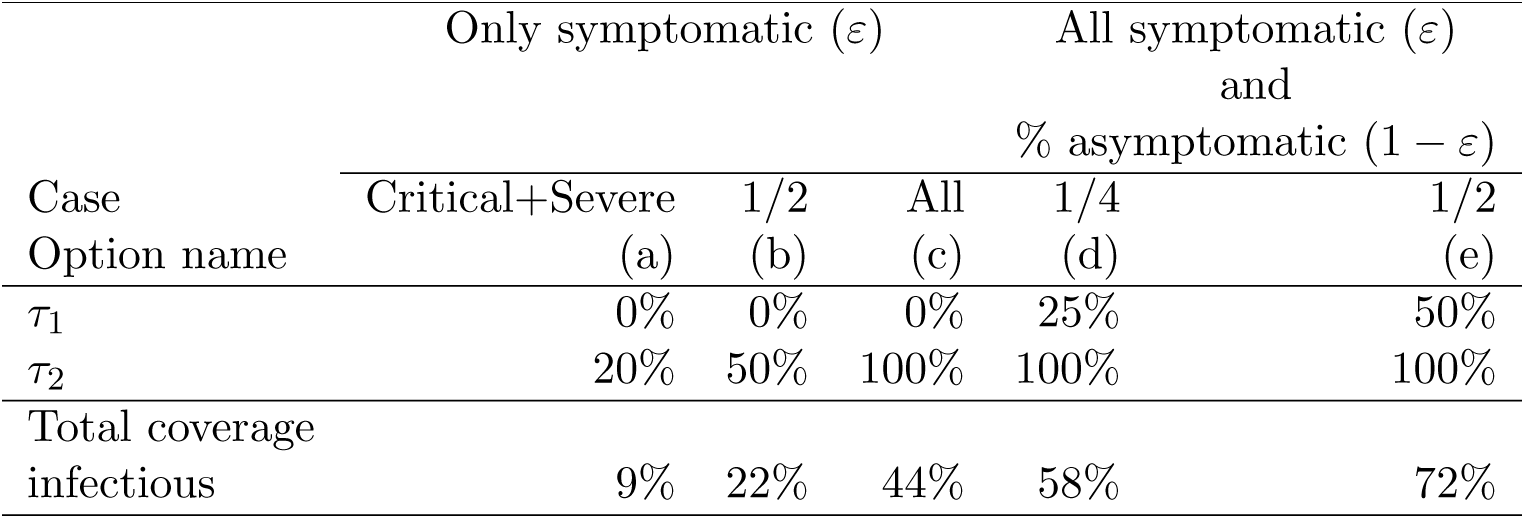
Testing options

| Case<br>Option name | Only symptomatic ($\varepsilon$) | | | All symptomatic ($\varepsilon$)<br>and<br>% asymptomatic ($1 - \varepsilon$) | |
| --- | --- | --- | --- | --- | --- |
|  | Critical+Severe | 1/2 | All | 1/4 | 1/2 |
|  | (a) | (b) | (c) | (d) | (e) |
| $\tau_1$ | 0% | 0% | 0% | 25% | 50% |
| $\tau_2$ | 20% | 50% | 100% | 100% | 100% |
| Total coverage<br>infectious | 9% | 22% | 44% | 58% | 72% |

In order to analyze the impact that different degrees of testing and isolation have on controlling the spread of COVID-19 we implement five different testing options, which are summarized in Table S3. In option (a) only individuals who develop critical or severe symptoms are tested {*τ*_1_ =0, *τ*_2_ = 0.2}. In option (b) half of the symptomatic individuals are tested {*τ*_1_ =0, *τ*_2_ = 0.5}. In option (c) all the symptomatic individuals are tested {*τ*_1_ =0, *τ*_2_ =1.0}. In option (d) all symptomatic are tested and also one-quarter of asymptomatic individuals { *τ*_1_ = 0.25, *τ*_2_ = 1.0}. In option (e) all symptomatic are tested and also half of the asymptomatic individuals {*τ*_1_ = 0.5, *τ*_2_ =1.0}.

In all the options summarized in Table S3 we assume governments do not start the intervention until one and a half months after the onset of the outbreak (i.e., *t_I_* = 45). The impact of alternative intervention days is analyzed in the next subsection. Under the assumption *t_I_* = 45, we show in Figure S9 the impact of testing on the fatality rate by mean-age of the population and on lives saved by population size. Panel A in Figure S9 shows that countries with aged populations (i.e. higher mean-age) have higher average fatality rates than countries with younger populations. However, given that the fatality rate only informs about the ratio between total deaths and total infected individuals, Panel A in Fig. S9 does not represents well how many people are saved by various testing strategies. The proportion of lives saved is defined as the relative difference between the total deaths without testing and the total deaths for a specific level of testing. Panel B in Figure S9 shows the average proportion of lives saved by testing and isolating after one year. Comparing across levels of testing Figure S9 shows that by testing and isolating individuals who develop severe or critical symptoms, the death toll is reduced by 2% in large countries. If the total coverage of testing reaches 22%, the death toll will be reduced by 9.4% in large countries. Only when all the symptomatic individuals are tested and isolated, which correspond to almost 44% of the total infectious population, the death toll is reduced by 46% in large countries. When all the symptomatic plus half of the asymptomatic are tested in large countries, the average reduction of the death toll reaches 98%. These results emphasizes the importance of mass testing. Indeed, Panel B, Figure S9, clearly shows that it is especially important to start mass testing sooner in countries with smaller populations.^11^

**Figure S9:**
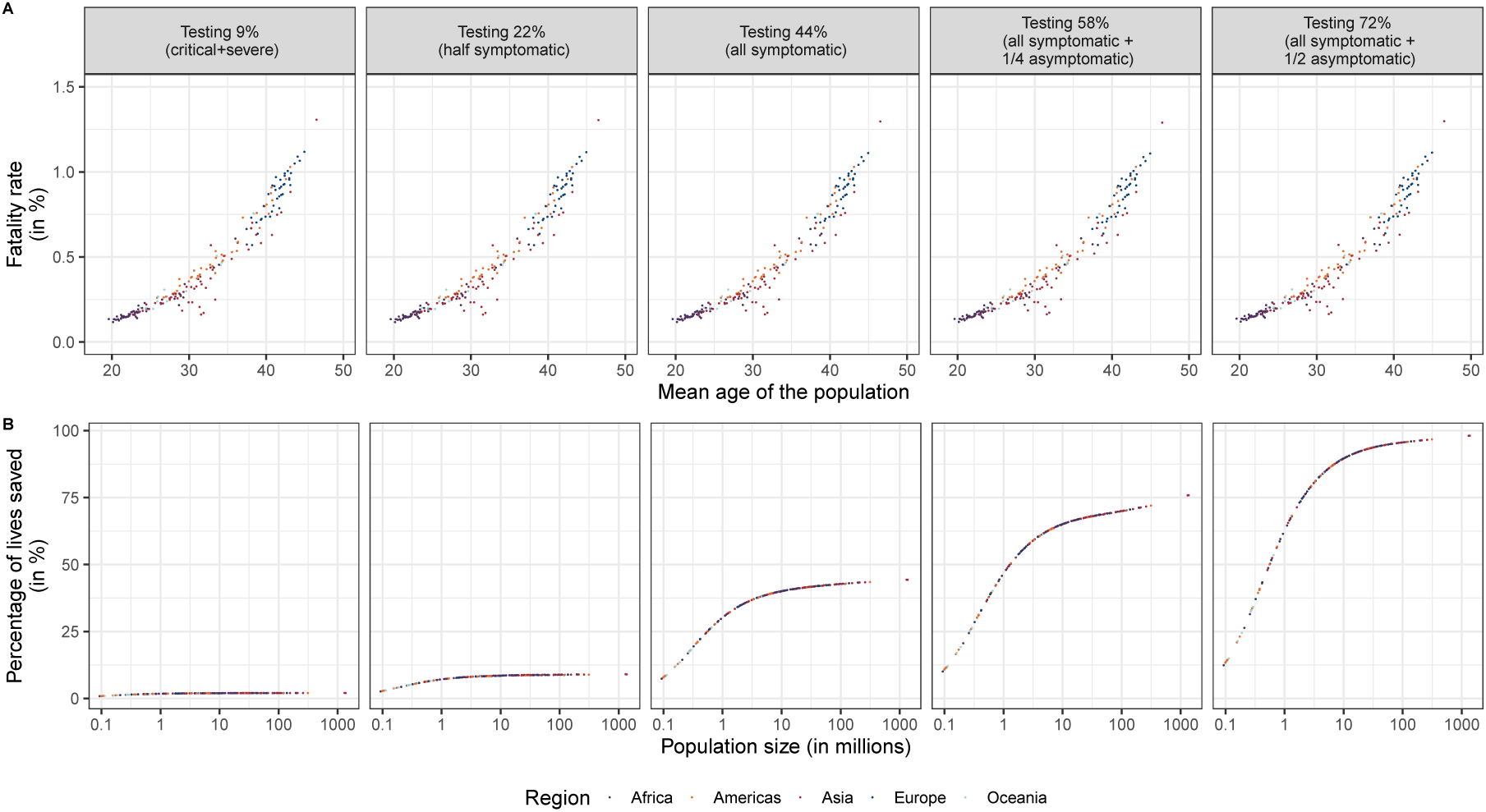
Fatality rate by mean age of the population (**A**) and proportion of lives saved by population size (**B**) at different levels of testing after one year (day=365). Source: UN Population data and authors’ calculations. Notes: Populations are assumed to be fully susceptible to COVID-19 and governments start testing after 45 days since the first imported and infected case.

### 3.2 The role of the intervention day

Another alternative for controlling the spread of COVID-19 is to start testing and isolating the infectious as soon as possible. To analyze the sensitivity of COVID-deaths with respect to the timing of testing we simulate eight alternative intervention days *t_I_*. The first possible day of intervention (*t_I_*) is set at day 31 and all subsequent dates of intervention, *t_I_*, are delayed by one week until *t_I_* = 80. Table S4 shows the impact of the day of intervention and the level of testing on the total number of deaths in the US after 365 days. For a given level of testing, the death toll is higher, the later the day of intervention. Similarly, for a given day of intervention, the lower is the level of testing, the higher is the death toll. Table S4 also shows that when the level of testing is lower than 45%, the day of intervention does not have a significant effect on the death toll. Therefore, in order to save more lives the best policy is testing all the infected individuals as soon as possible.

**Table S4:**
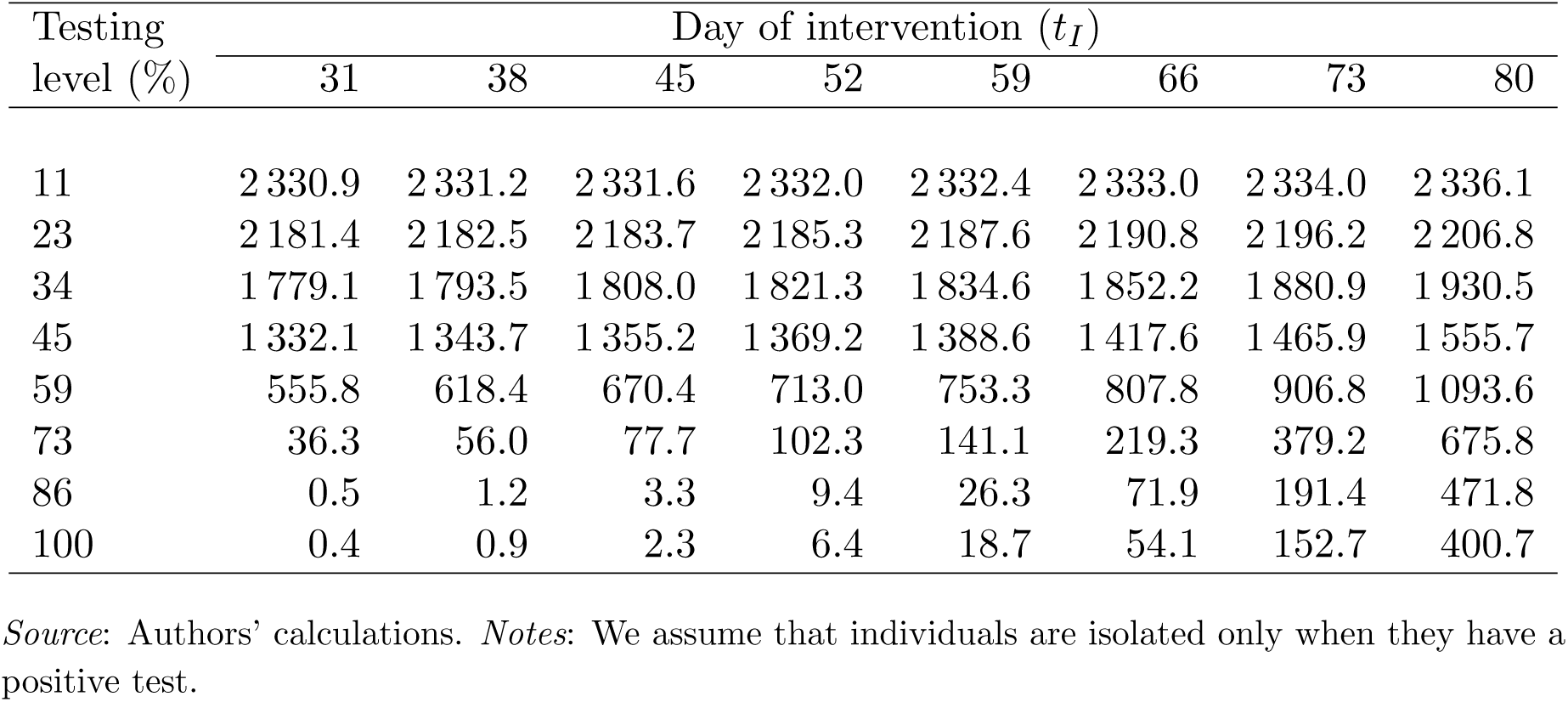
Average death toll by day of intervention and testing level in US after 365 days (in 1 000s)

| Testing<br>level (%) | Day of intervention ($t_I$) | | | | | | | |
| --- | --- | --- | --- | --- | --- | --- | --- | --- |
|  | 31 | 38 | 45 | 52 | 59 | 66 | 73 | 80 |
| 11 | 2 330.9 | 2 331.2 | 2 331.6 | 2 332.0 | 2 332.4 | 2 333.0 | 2 334.0 | 2 336.1 |
| 23 | 2 181.4 | 2 182.5 | 2 183.7 | 2 185.3 | 2 187.6 | 2 190.8 | 2 196.2 | 2 206.8 |
| 34 | 1 779.1 | 1 793.5 | 1 808.0 | 1 821.3 | 1 834.6 | 1 852.2 | 1 880.9 | 1 930.5 |
| 45 | 1 332.1 | 1 343.7 | 1 355.2 | 1 369.2 | 1 388.6 | 1 417.6 | 1 465.9 | 1 555.7 |
| 59 | 555.8 | 618.4 | 670.4 | 713.0 | 753.3 | 807.8 | 906.8 | 1 093.6 |
| 73 | 36.3 | 56.0 | 77.7 | 102.3 | 141.1 | 219.3 | 379.2 | 675.8 |
| 86 | 0.5 | 1.2 | 3.3 | 9.4 | 26.3 | 71.9 | 191.4 | 471.8 |
| 100 | 0.4 | 0.9 | 2.3 | 6.4 | 18.7 | 54.1 | 152.7 | 400.7 |

*Source:* Authors' calculations. *Notes:* We assume that individuals are isolated only when they have a positive test.

### 3.3 The role of herd immunity

In this subsection we study the impact of herd immunity on reducing the future death toll. We define herd immunity as the resistance to the spread of the disease due to the existence of recovered individuals who are immune to the virus. Thus, herd immunity will prevent the spread of the virus by reducing the infection rate *β_t_*. To study the effect of herd immunity we run the model, for all the countries contained in Tab. S2, setting the intervention day (*t_I_*) at time 0 and assuming three alternative scenarios in which the total proportion of recovered people is 25%, 50%, and 75%. We set *t_I_* at 0 because we assume that countries who have attained a specific level of herd immunity will continue testing individuals.

Panel A in Figure S10 shows the relationship between the average COVID-19 fatality rate and the mean age of the population for five different levels of testing and three possible degrees of herd immunity. It is not surprising that the fatality rate does not change much across different levels of herd immunity, since both the number of deaths and the number of infected individuals decrease. As a consequence, the fatality rate becomes rather insensitive to the herd immunity level. The herd immunity level, however, changes the percentage of lives saved relative to the population size. Panel B in Figure S10 shows that, for a given level of testing, the higher is the herd immunity level in a population, the greater is the percentage of lives saved (relative to the initial susceptible population). Moreover, this effect is more intense in large populations and the greater is the level of testing.

**Figure S10:**
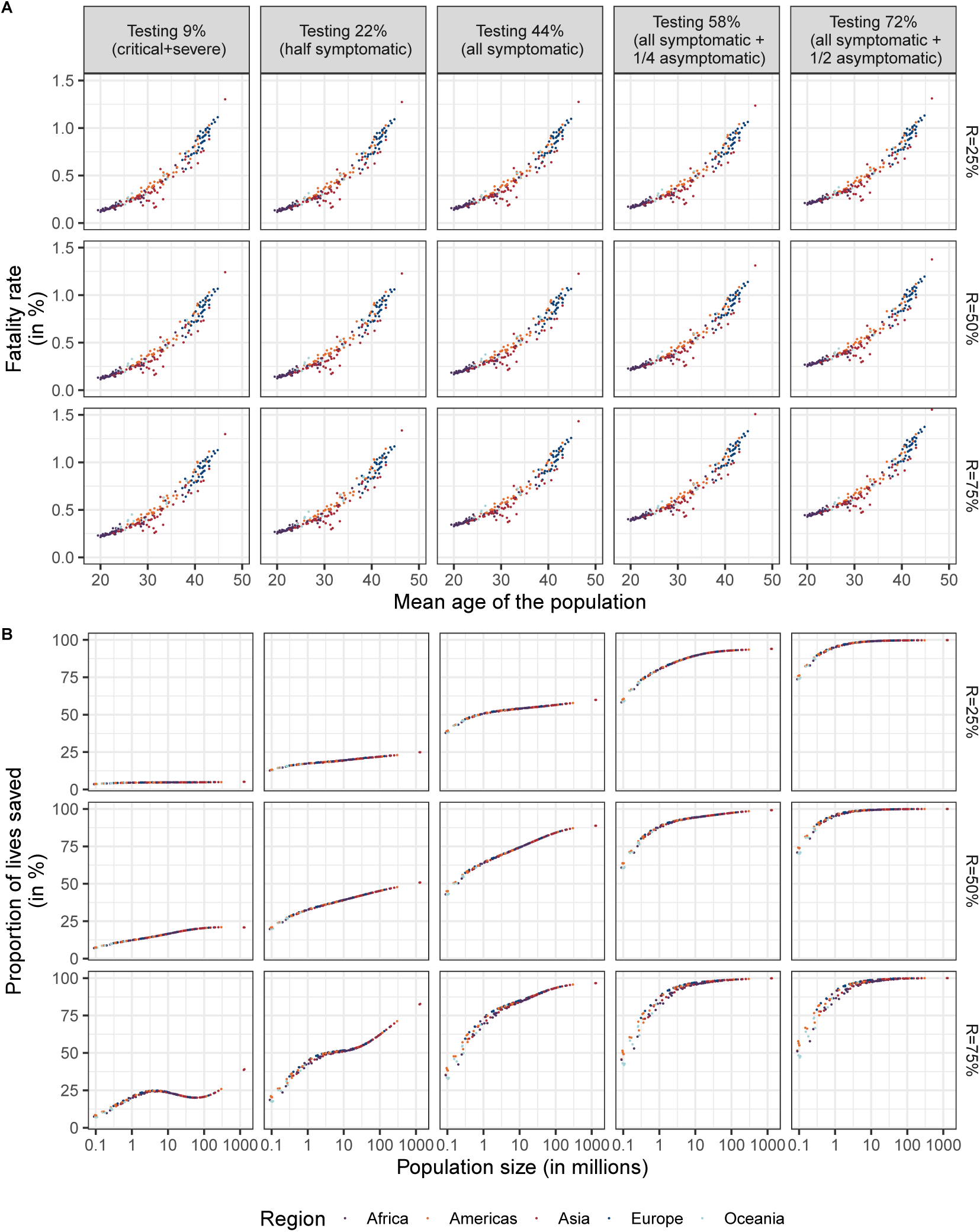
Fatality rate by mean age of the population (**A**) and proportion of lives saved by population size (**B**) at different levels of testing and herd immunity level after one year (day=365). *Source*: UN Population data and authors’ calculations.

## 4 Total infected individuals and the case fatality rates

In this section we investigate how to infer the number of infected people in each country using the model. To do so, we use the case fatality rate, which differs from the (true) fatality rate used in previous sections. We define the case fatality rate (CFR) as the ratio between the total number of deaths and the total number of infected individuals that are detected. Note that information about the total number of deaths and the total number of infected and detected individuals are provided in country reports. In our case we derive the CFR dividing the (true) fatality rate by the fraction of infected individuals who are tested.

**Figure S11:**
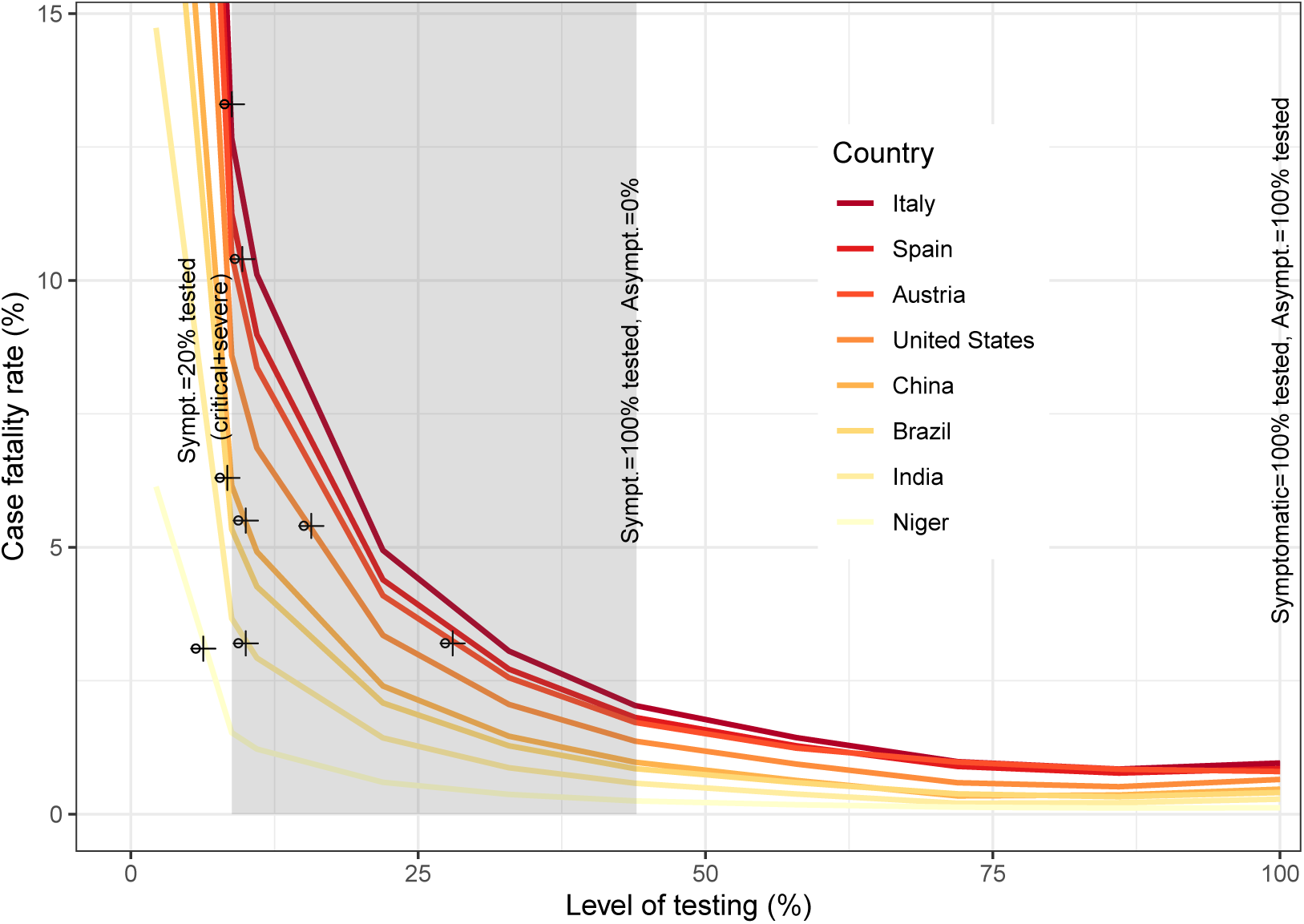
Average case fatality rate (CFR) by level of testing according to symptomatic and asymptomatic individuals after 365 days. *Source*: Authors’ calculations.

Figure S11 shows the CFR that would be reached for a given level of testing, that is maintained after one year since the onset of the epidemic outbreak, in eight selected countries (Italy, Spain, Austria, USA, China, Brazil, India, and Niger). By using the CFR in day 365, the CFR value reported is the maximum that a country will reach under a specific level of testing.^12^ Equivalently, each country curve, depicted in Fig. S11, also shows the maximum level of testing for a given CFR value. The gray area represents the level of testing for the symptomatic individuals. For each country curve, the CFR values at the left-hand side of the gray area correspond to testing infected individuals who only develop critical and severe symptoms, or 20% of the symptomatic individuals. The CFR values at the right-hand side of the gray area correspond to testing 100% of the symptomatic individuals. While the true fatality rates and the CFRs only coincide when the level of testing is one hundred percent, which corresponds to the far right point in the diagram.

The CFR values until April 21st, 2020 were 13.3% (=24114 deaths/181 228 cases) in Italy, 10.4%(=20 852/200 210) in Spain, 3.2% (=470/14 783) in Austria, 5.4% (=42 539/787 752) in US, 5.5% (=4 636/83 849) in China, 6.3% (=2575/40 581) in Brazil, 3.2% (=590/18 600) in India, and 3.1% (=20/655) in Niger. Thus, according to Fig. S11 the CFR values in China, India, Italy, and Spain suggest that these countries are mainly testing individuals who develop critical and severe symptoms (see the crossed circles over each country curve). Hence, the total number of infected people is at least ten times higher than those being reported in China, India, Italy, and Spain. The CFR values in Brazil and Niger suggest that less than 9% of the total infected individuals are tested. Thereby, the total number of infected people is more than ten times higher than those being reported in these two countries. In the US, the CFR value suggests that close to 15% of the infected individuals are tested (see the crossed circle over the orange curve) and therefore it is likely that total number of infected people is more than six times higher than those being detected. In Austria, which is a country that started testing at high proportions, the CFR value suggests that around 25% of the infected individuals are tested (see the crossed circle in third curve starting from the top). As a result, the total number of infected individuals is at least more than four times the total infected individuals tested, which coincides with the lower bound estimated by (5) for Austria.

**Figure S12:**
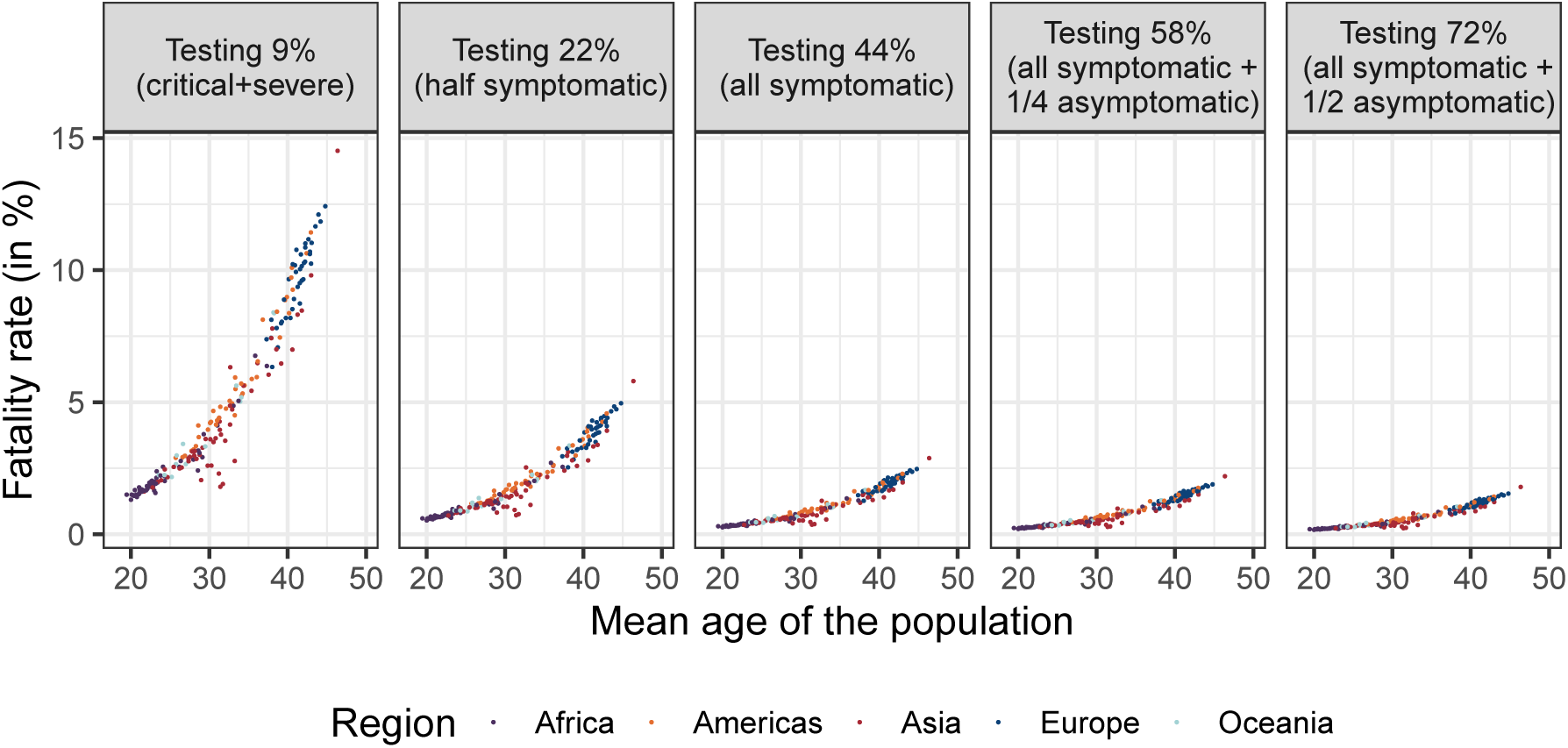
Average relationship between the COVID-19 case fatality rate and the mean age of the population by level of testing (day=365). *Source*: UN Population data and authors’ calculations.

We extend the analysis to the whole world by showing in Figure S12 and Table S7 the average CFR across countries by mean age of the population and testing level. When only testing at individuals who develop severe or critical symptoms, Fig. S12 shows that the CFRs will be above 10% in countries with a mean age of the population above 42 years. In contrast, when only testing those individuals with severe and critical symptoms, the CFRs will be close to 2% in countries with a mean age of the population below 25 years.^13^ Once that the level of testing reaches all the symptomatic individuals (i.e. 44% of all infected), we observe values for the CFR lower than 2.5% in all countries. Moreover, Fig. S12 shows that the CFRs can be halved with respect to testing all the symptomatic individuals by testing at least 72% of the total infected population.

In sum, despite the uncertainty associated to the COVID-19 outbreak, we have found an important feature that may help us to shed light on the number of people infected. This is because the day of intervention, the population size, and the herd immunity affect on the total number of people infected, as shown in Table S4 and Figs. S9 and S10. However, conditional on being infected the probability of dying from the epidemic (or fatality rate) just depends on age. Thus, given the demographic characteristic of each population —e.g. the mean age of the population— we can infer through the case fatality rate the total number of people infected in each country.

## 5 The impact of COVID-19 in the World

We complement the information provided in previous sections with four additional tables. Table S5 shows the death toll of COVID-19 for each region in the world by degree of testing under fully susceptible populations. In Table S6 we report the distribution of the fatality rate of COVID-19 obtained through the Bayesian Melding method for 200 countries. Table S7 reports the average case fatality rates by level of testing after one year in the 200 countries plotted in Fig. S12. Table S8 shows the average death toll from COVID-19 for 200 countries by level of testing in fully susceptible populations after 365 days.

**Table S5:** Death toll of COVID-19 for each region in the world by degree of testing in fully susceptible populations after 365 days: Day of intervention=45.

| Testing options | World | Region |  |  |  |  |
| --- | --- | --- | --- | --- | --- | --- |
|  |  | Africa | Americas | Asia | Europe | Oceania |
| Testing 0% |  |  |  |  |  |  |
| Deaths (in 1000s) | 33 350.5 | 2 380.7 | 5 460.0 | 18 900.2 | 6 364.9 | 244.8 |
| Avg. fatality rate (in %) | 0.378 | 0.172 | 0.440 | 0.315 | 0.719 | 0.319 |
| Testing 9% |  |  |  |  |  |  |
| Deaths (in 1000s) | 32 678.6 | 2 332.8 | 5 350.7 | 18 516.9 | 6 238.2 | 240.0 |
| Avg. fatality rate (in %) | 0.358 | 0.163 | 0.421 | 0.295 | 0.679 | 0.308 |
| Testing 22% |  |  |  |  |  |  |
| Deaths (in 1000s) | 30 402.6 | 2171.9 | 4 980.4 | 17 214.3 | 5 811.9 | 224.1 |
| Avg. fatality rate (in %) | 0.318 | 0.144 | 0.383 | 0.256 | 0.596 | 0.287 |
| Testing 44% |  |  |  |  |  |  |
| Deaths (in 1000s) | 19 047.0 | 1 385.0 | 3 137.2 | 10 663.2 | 3 714.7 | 146.8 |
| Avg. fatality rate (in %) | 0.227 | 0.102 | 0.295 | 0.172 | 0.407 | 0.235 |
| Testing 58% |  |  |  |  |  |  |
| Deaths (in 1000s) | 9 546.3 | 755.2 | 1 638.0 | 5 025.6 | 2 041.8 | 85.6 |
| Avg. fatality rate (in %) | 0.219 | 0.099 | 0.289 | 0.164 | 0.391 | 0.232 |
| Testing 72% |  |  |  |  |  |  |
| Deaths (in 1000s) | 1 487.2 | 153.9 | 275.3 | 596.3 | 436.1 | 25.6 |
| Avg. fatality rate (in %) | 0.222 | 0.101 | 0.293 | 0.159 | 0.405 | 0.239 |

**Table S6:**
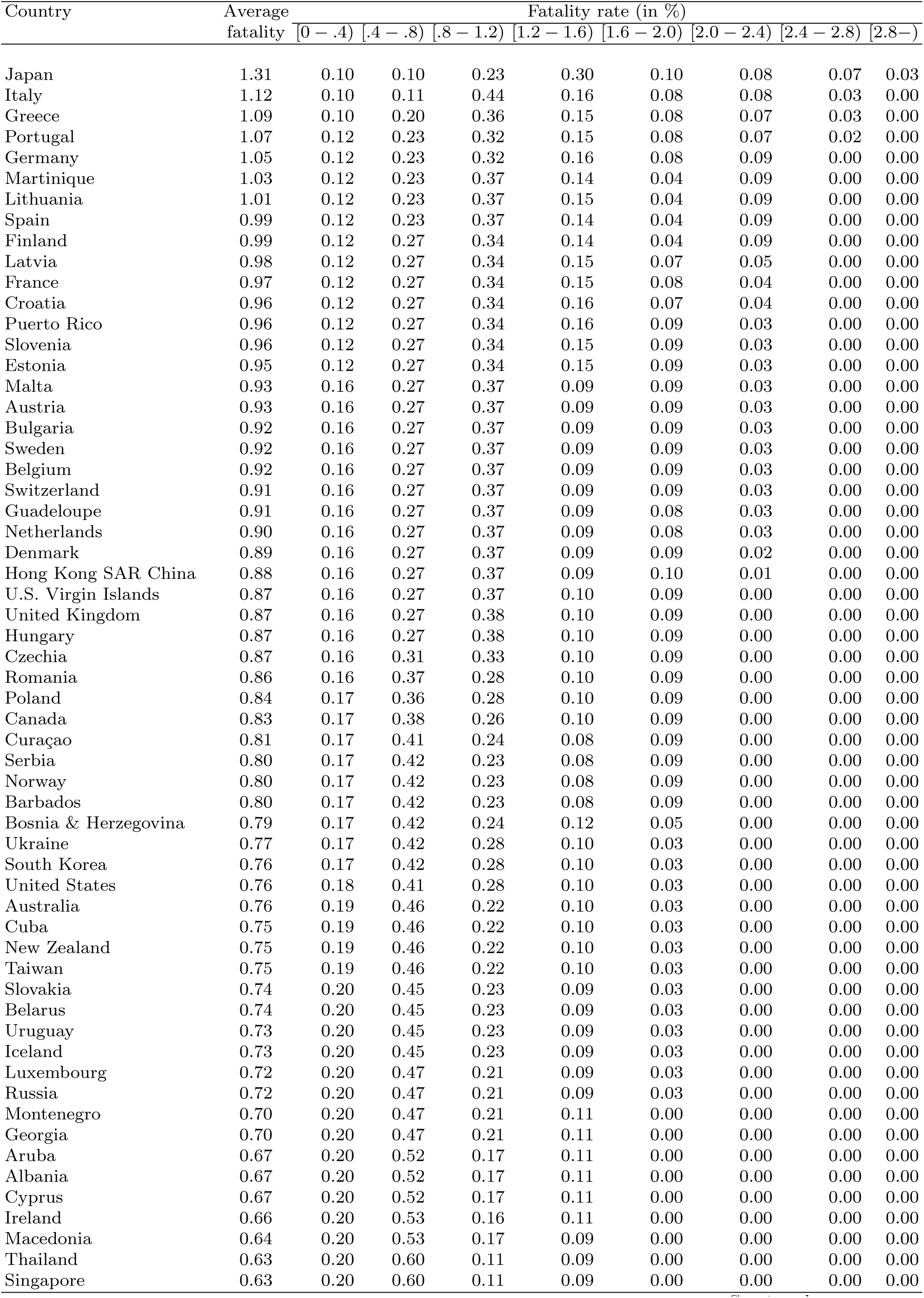
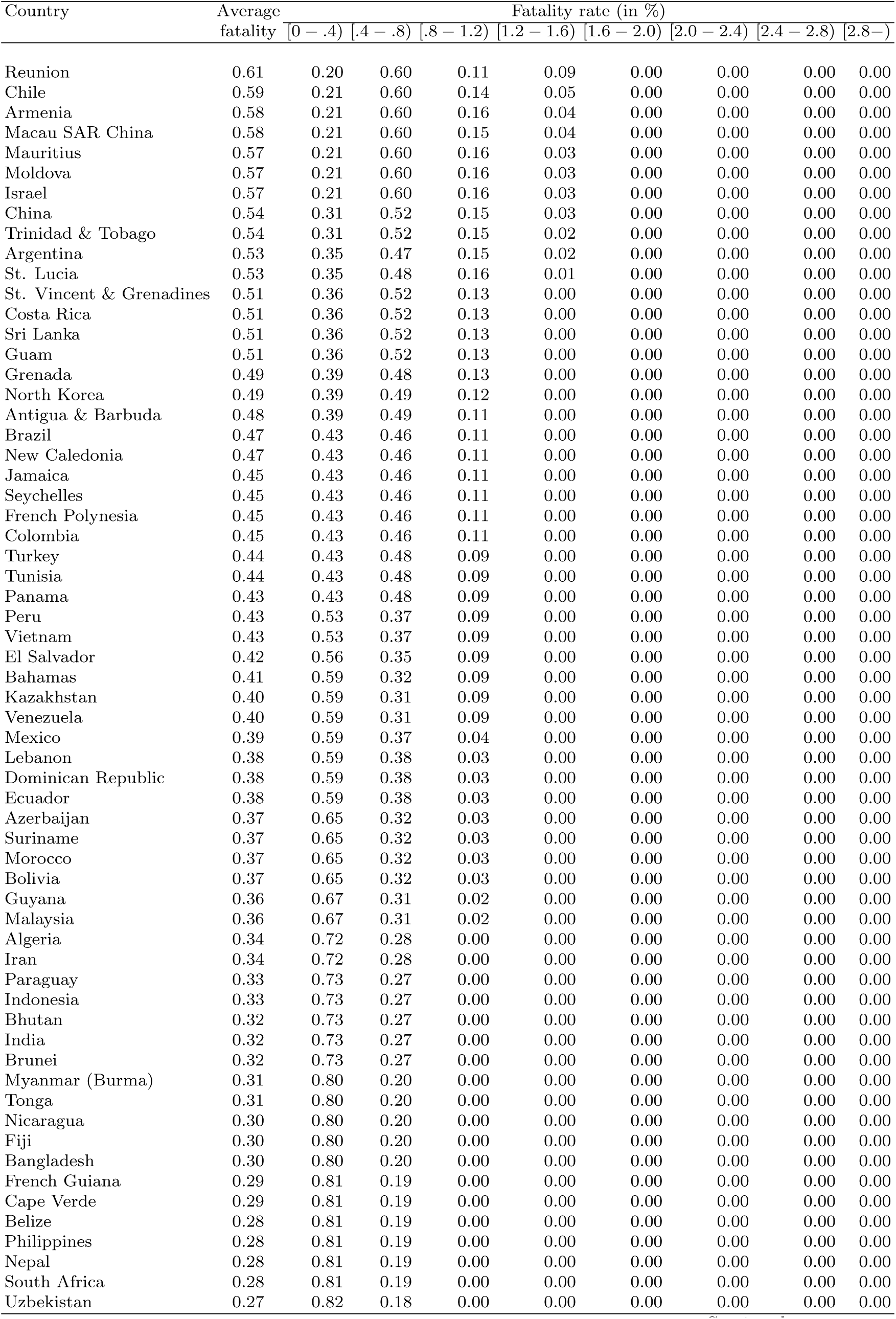
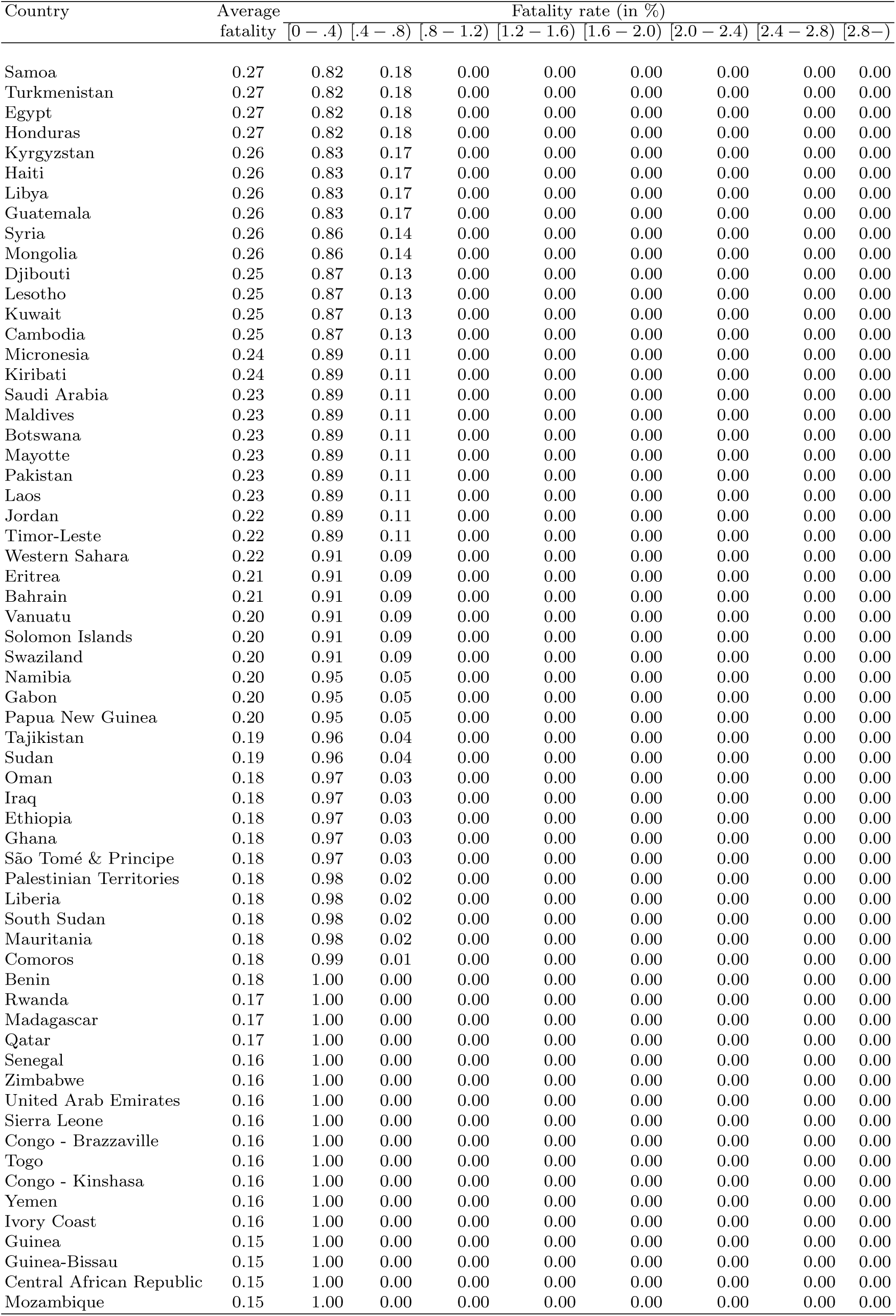
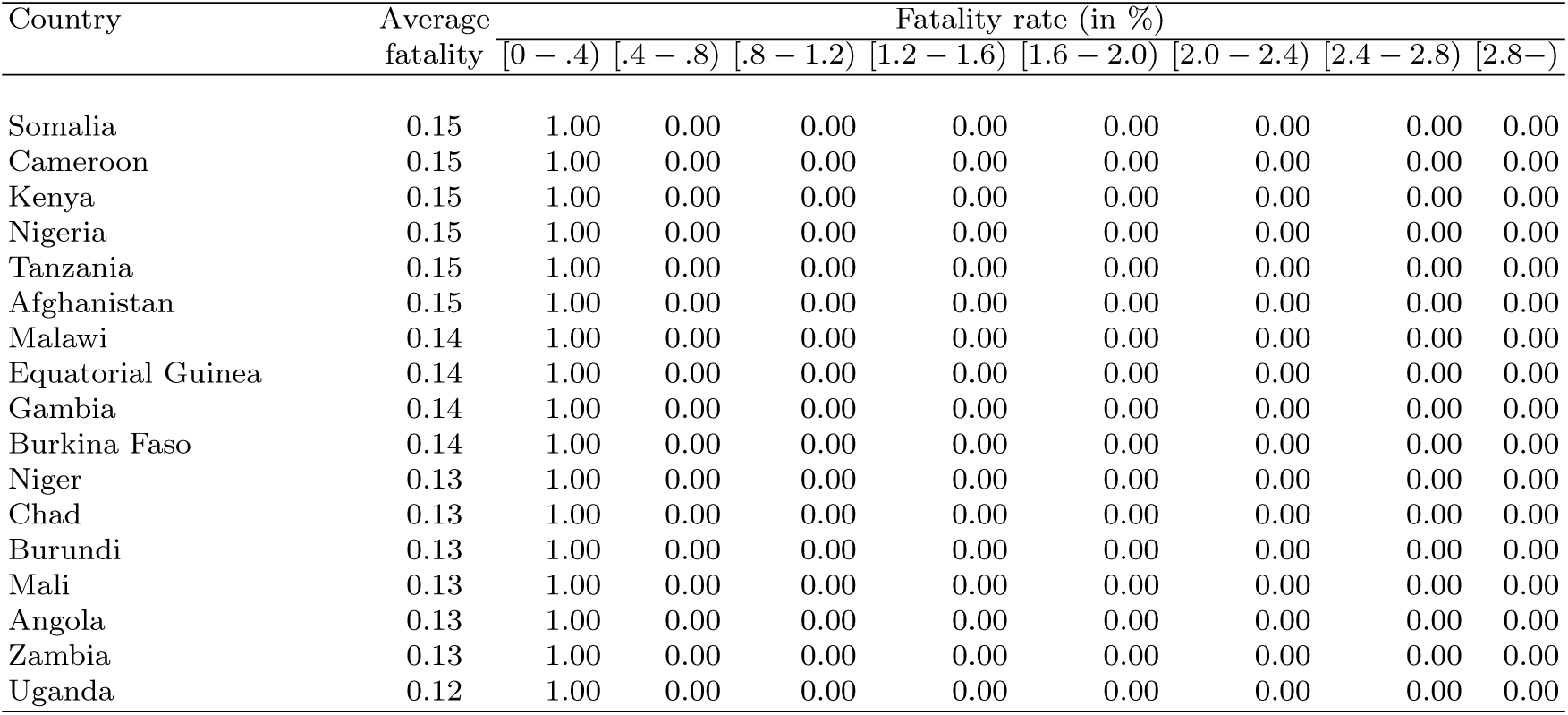
Distribution of the fatality rate of COVID-19 after 365 days in fully susceptible populations. Countries ordered from high to low fatality rate level.

| Country | Average fatality | Fatality rate (in %) |  |  |  |  |  |  |  |
| --- | --- | --- | --- | --- | --- | --- | --- | --- | --- |
|  |  | [0 – .4) | [.4 – .8) | [.8 – 1.2) | [1.2 – 1.6) | [1.6 – 2.0) | [2.0 – 2.4) | [2.4 – 2.8) | [2.8 – ) |
| Japan | 1.31 | 0.10 | 0.10 | 0.23 | 0.30 | 0.10 | 0.08 | 0.07 | 0.03 |
| Italy | 1.12 | 0.10 | 0.11 | 0.44 | 0.16 | 0.08 | 0.08 | 0.03 | 0.00 |
| Greece | 1.09 | 0.10 | 0.20 | 0.36 | 0.15 | 0.08 | 0.07 | 0.03 | 0.00 |
| Portugal | 1.07 | 0.12 | 0.23 | 0.32 | 0.15 | 0.08 | 0.07 | 0.02 | 0.00 |
| Germany | 1.05 | 0.12 | 0.23 | 0.32 | 0.16 | 0.08 | 0.09 | 0.00 | 0.00 |
| Martinique | 1.03 | 0.12 | 0.23 | 0.37 | 0.14 | 0.04 | 0.09 | 0.00 | 0.00 |
| Lithuania | 1.01 | 0.12 | 0.23 | 0.37 | 0.15 | 0.04 | 0.09 | 0.00 | 0.00 |
| Spain | 0.99 | 0.12 | 0.23 | 0.37 | 0.14 | 0.04 | 0.09 | 0.00 | 0.00 |
| Finland | 0.99 | 0.12 | 0.27 | 0.34 | 0.14 | 0.04 | 0.09 | 0.00 | 0.00 |
| Latvia | 0.98 | 0.12 | 0.27 | 0.34 | 0.15 | 0.07 | 0.05 | 0.00 | 0.00 |
| France | 0.97 | 0.12 | 0.27 | 0.34 | 0.15 | 0.08 | 0.04 | 0.00 | 0.00 |
| Croatia | 0.96 | 0.12 | 0.27 | 0.34 | 0.16 | 0.07 | 0.04 | 0.00 | 0.00 |
| Puerto Rico | 0.96 | 0.12 | 0.27 | 0.34 | 0.16 | 0.09 | 0.03 | 0.00 | 0.00 |
| Slovenia | 0.96 | 0.12 | 0.27 | 0.34 | 0.15 | 0.09 | 0.03 | 0.00 | 0.00 |
| Estonia | 0.95 | 0.12 | 0.27 | 0.34 | 0.15 | 0.09 | 0.03 | 0.00 | 0.00 |
| Malta | 0.93 | 0.16 | 0.27 | 0.37 | 0.09 | 0.09 | 0.03 | 0.00 | 0.00 |
| Austria | 0.93 | 0.16 | 0.27 | 0.37 | 0.09 | 0.09 | 0.03 | 0.00 | 0.00 |
| Bulgaria | 0.92 | 0.16 | 0.27 | 0.37 | 0.09 | 0.09 | 0.03 | 0.00 | 0.00 |
| Sweden | 0.92 | 0.16 | 0.27 | 0.37 | 0.09 | 0.09 | 0.03 | 0.00 | 0.00 |
| Belgium | 0.92 | 0.16 | 0.27 | 0.37 | 0.09 | 0.09 | 0.03 | 0.00 | 0.00 |
| Switzerland | 0.91 | 0.16 | 0.27 | 0.37 | 0.09 | 0.09 | 0.03 | 0.00 | 0.00 |
| Guadeloupe | 0.91 | 0.16 | 0.27 | 0.37 | 0.09 | 0.08 | 0.03 | 0.00 | 0.00 |
| Netherlands | 0.90 | 0.16 | 0.27 | 0.37 | 0.09 | 0.08 | 0.03 | 0.00 | 0.00 |
| Denmark | 0.89 | 0.16 | 0.27 | 0.37 | 0.09 | 0.09 | 0.02 | 0.00 | 0.00 |
| Hong Kong SAR China | 0.88 | 0.16 | 0.27 | 0.37 | 0.09 | 0.10 | 0.01 | 0.00 | 0.00 |
| U.S. Virgin Islands | 0.87 | 0.16 | 0.27 | 0.37 | 0.10 | 0.09 | 0.00 | 0.00 | 0.00 |
| United Kingdom | 0.87 | 0.16 | 0.27 | 0.38 | 0.10 | 0.09 | 0.00 | 0.00 | 0.00 |
| Hungary | 0.87 | 0.16 | 0.27 | 0.38 | 0.10 | 0.09 | 0.00 | 0.00 | 0.00 |
| Czechia | 0.87 | 0.16 | 0.31 | 0.33 | 0.10 | 0.09 | 0.00 | 0.00 | 0.00 |
| Romania | 0.86 | 0.16 | 0.37 | 0.28 | 0.10 | 0.09 | 0.00 | 0.00 | 0.00 |
| Poland | 0.84 | 0.17 | 0.36 | 0.28 | 0.10 | 0.09 | 0.00 | 0.00 | 0.00 |
| Canada | 0.83 | 0.17 | 0.38 | 0.26 | 0.10 | 0.09 | 0.00 | 0.00 | 0.00 |
| Curaçao | 0.81 | 0.17 | 0.41 | 0.24 | 0.08 | 0.09 | 0.00 | 0.00 | 0.00 |
| Serbia | 0.80 | 0.17 | 0.42 | 0.23 | 0.08 | 0.09 | 0.00 | 0.00 | 0.00 |
| Norway | 0.80 | 0.17 | 0.42 | 0.23 | 0.08 | 0.09 | 0.00 | 0.00 | 0.00 |
| Barbados | 0.80 | 0.17 | 0.42 | 0.23 | 0.08 | 0.09 | 0.00 | 0.00 | 0.00 |
| Bosnia & Herzegovina | 0.79 | 0.17 | 0.42 | 0.24 | 0.12 | 0.05 | 0.00 | 0.00 | 0.00 |
| Ukraine | 0.77 | 0.17 | 0.42 | 0.28 | 0.10 | 0.03 | 0.00 | 0.00 | 0.00 |
| South Korea | 0.76 | 0.17 | 0.42 | 0.28 | 0.10 | 0.03 | 0.00 | 0.00 | 0.00 |
| United States | 0.76 | 0.18 | 0.41 | 0.28 | 0.10 | 0.03 | 0.00 | 0.00 | 0.00 |
| Australia | 0.76 | 0.19 | 0.46 | 0.22 | 0.10 | 0.03 | 0.00 | 0.00 | 0.00 |
| Cuba | 0.75 | 0.19 | 0.46 | 0.22 | 0.10 | 0.03 | 0.00 | 0.00 | 0.00 |
| New Zealand | 0.75 | 0.19 | 0.46 | 0.22 | 0.10 | 0.03 | 0.00 | 0.00 | 0.00 |
| Taiwan | 0.75 | 0.19 | 0.46 | 0.22 | 0.10 | 0.03 | 0.00 | 0.00 | 0.00 |
| Slovakia | 0.74 | 0.20 | 0.45 | 0.23 | 0.09 | 0.03 | 0.00 | 0.00 | 0.00 |
| Belarus | 0.74 | 0.20 | 0.45 | 0.23 | 0.09 | 0.03 | 0.00 | 0.00 | 0.00 |
| Uruguay | 0.73 | 0.20 | 0.45 | 0.23 | 0.09 | 0.03 | 0.00 | 0.00 | 0.00 |
| Iceland | 0.73 | 0.20 | 0.45 | 0.23 | 0.09 | 0.03 | 0.00 | 0.00 | 0.00 |
| Luxembourg | 0.72 | 0.20 | 0.47 | 0.21 | 0.09 | 0.03 | 0.00 | 0.00 | 0.00 |
| Russia | 0.72 | 0.20 | 0.47 | 0.21 | 0.09 | 0.03 | 0.00 | 0.00 | 0.00 |
| Montenegro | 0.70 | 0.20 | 0.47 | 0.21 | 0.11 | 0.00 | 0.00 | 0.00 | 0.00 |
| Georgia | 0.70 | 0.20 | 0.47 | 0.21 | 0.11 | 0.00 | 0.00 | 0.00 | 0.00 |
| Aruba | 0.67 | 0.20 | 0.52 | 0.17 | 0.11 | 0.00 | 0.00 | 0.00 | 0.00 |
| Albania | 0.67 | 0.20 | 0.52 | 0.17 | 0.11 | 0.00 | 0.00 | 0.00 | 0.00 |
| Cyprus | 0.67 | 0.20 | 0.52 | 0.17 | 0.11 | 0.00 | 0.00 | 0.00 | 0.00 |
| Ireland | 0.66 | 0.20 | 0.53 | 0.16 | 0.11 | 0.00 | 0.00 | 0.00 | 0.00 |
| Macedonia | 0.64 | 0.20 | 0.53 | 0.17 | 0.09 | 0.00 | 0.00 | 0.00 | 0.00 |
| Thailand | 0.63 | 0.20 | 0.60 | 0.11 | 0.09 | 0.00 | 0.00 | 0.00 | 0.00 |
| Singapore | 0.63 | 0.20 | 0.60 | 0.11 | 0.09 | 0.00 | 0.00 | 0.00 | 0.00 |
|  |  | [0 – .4) | [.4 – .8) | [.8 – 1.2) | [1.2 – 1.6) | [1.6 – 2.0) | [2.0 – 2.4) | [2.4 – 2.8) | [2.8 – ) |
| Reunion | 0.61 | 0.20 | 0.60 | 0.11 | 0.09 | 0.00 | 0.00 | 0.00 | 0.00 |
| Chile | 0.59 | 0.21 | 0.60 | 0.14 | 0.05 | 0.00 | 0.00 | 0.00 | 0.00 |
| Armenia | 0.58 | 0.21 | 0.60 | 0.16 | 0.04 | 0.00 | 0.00 | 0.00 | 0.00 |
| Macau SAR China | 0.58 | 0.21 | 0.60 | 0.15 | 0.04 | 0.00 | 0.00 | 0.00 | 0.00 |
| Mauritius | 0.57 | 0.21 | 0.60 | 0.16 | 0.03 | 0.00 | 0.00 | 0.00 | 0.00 |
| Moldova | 0.57 | 0.21 | 0.60 | 0.16 | 0.03 | 0.00 | 0.00 | 0.00 | 0.00 |
| Israel | 0.57 | 0.21 | 0.60 | 0.16 | 0.03 | 0.00 | 0.00 | 0.00 | 0.00 |
| China | 0.54 | 0.31 | 0.52 | 0.15 | 0.03 | 0.00 | 0.00 | 0.00 | 0.00 |
| Trinidad & Tobago | 0.54 | 0.31 | 0.52 | 0.15 | 0.02 | 0.00 | 0.00 | 0.00 | 0.00 |
| Argentina | 0.53 | 0.35 | 0.47 | 0.15 | 0.02 | 0.00 | 0.00 | 0.00 | 0.00 |
| St. Lucia | 0.53 | 0.35 | 0.48 | 0.16 | 0.01 | 0.00 | 0.00 | 0.00 | 0.00 |
| St. Vincent & Grenadines | 0.51 | 0.36 | 0.52 | 0.13 | 0.00 | 0.00 | 0.00 | 0.00 | 0.00 |
| Costa Rica | 0.51 | 0.36 | 0.52 | 0.13 | 0.00 | 0.00 | 0.00 | 0.00 | 0.00 |
| Sri Lanka | 0.51 | 0.36 | 0.52 | 0.13 | 0.00 | 0.00 | 0.00 | 0.00 | 0.00 |
| Guam | 0.51 | 0.36 | 0.52 | 0.13 | 0.00 | 0.00 | 0.00 | 0.00 | 0.00 |
| Grenada | 0.49 | 0.39 | 0.48 | 0.13 | 0.00 | 0.00 | 0.00 | 0.00 | 0.00 |
| North Korea | 0.49 | 0.39 | 0.49 | 0.12 | 0.00 | 0.00 | 0.00 | 0.00 | 0.00 |
| Antigua & Barbuda | 0.48 | 0.39 | 0.49 | 0.11 | 0.00 | 0.00 | 0.00 | 0.00 | 0.00 |
| Brazil | 0.47 | 0.43 | 0.46 | 0.11 | 0.00 | 0.00 | 0.00 | 0.00 | 0.00 |
| New Caledonia | 0.47 | 0.43 | 0.46 | 0.11 | 0.00 | 0.00 | 0.00 | 0.00 | 0.00 |
| Jamaica | 0.45 | 0.43 | 0.46 | 0.11 | 0.00 | 0.00 | 0.00 | 0.00 | 0.00 |
| Seychelles | 0.45 | 0.43 | 0.46 | 0.11 | 0.00 | 0.00 | 0.00 | 0.00 | 0.00 |
| French Polynesia | 0.45 | 0.43 | 0.46 | 0.11 | 0.00 | 0.00 | 0.00 | 0.00 | 0.00 |
| Colombia | 0.45 | 0.43 | 0.46 | 0.11 | 0.00 | 0.00 | 0.00 | 0.00 | 0.00 |
| Turkey | 0.44 | 0.43 | 0.48 | 0.09 | 0.00 | 0.00 | 0.00 | 0.00 | 0.00 |
| Tunisia | 0.44 | 0.43 | 0.48 | 0.09 | 0.00 | 0.00 | 0.00 | 0.00 | 0.00 |
| Panama | 0.43 | 0.43 | 0.48 | 0.09 | 0.00 | 0.00 | 0.00 | 0.00 | 0.00 |
| Peru | 0.43 | 0.53 | 0.37 | 0.09 | 0.00 | 0.00 | 0.00 | 0.00 | 0.00 |
| Vietnam | 0.43 | 0.53 | 0.37 | 0.09 | 0.00 | 0.00 | 0.00 | 0.00 | 0.00 |
| El Salvador | 0.42 | 0.56 | 0.35 | 0.09 | 0.00 | 0.00 | 0.00 | 0.00 | 0.00 |
| Bahamas | 0.41 | 0.59 | 0.32 | 0.09 | 0.00 | 0.00 | 0.00 | 0.00 | 0.00 |
| Kazakhstan | 0.40 | 0.59 | 0.31 | 0.09 | 0.00 | 0.00 | 0.00 | 0.00 | 0.00 |
| Venezuela | 0.40 | 0.59 | 0.31 | 0.09 | 0.00 | 0.00 | 0.00 | 0.00 | 0.00 |
| Mexico | 0.39 | 0.59 | 0.37 | 0.04 | 0.00 | 0.00 | 0.00 | 0.00 | 0.00 |
| Lebanon | 0.38 | 0.59 | 0.38 | 0.03 | 0.00 | 0.00 | 0.00 | 0.00 | 0.00 |
| Dominican Republic | 0.38 | 0.59 | 0.38 | 0.03 | 0.00 | 0.00 | 0.00 | 0.00 | 0.00 |
| Ecuador | 0.38 | 0.59 | 0.38 | 0.03 | 0.00 | 0.00 | 0.00 | 0.00 | 0.00 |
| Azerbaijan | 0.37 | 0.65 | 0.32 | 0.03 | 0.00 | 0.00 | 0.00 | 0.00 | 0.00 |
| Suriname | 0.37 | 0.65 | 0.32 | 0.03 | 0.00 | 0.00 | 0.00 | 0.00 | 0.00 |
| Morocco | 0.37 | 0.65 | 0.32 | 0.03 | 0.00 | 0.00 | 0.00 | 0.00 | 0.00 |
| Bolivia | 0.37 | 0.65 | 0.32 | 0.03 | 0.00 | 0.00 | 0.00 | 0.00 | 0.00 |
| Guyana | 0.36 | 0.67 | 0.31 | 0.02 | 0.00 | 0.00 | 0.00 | 0.00 | 0.00 |
| Malaysia | 0.36 | 0.67 | 0.31 | 0.02 | 0.00 | 0.00 | 0.00 | 0.00 | 0.00 |
| Algeria | 0.34 | 0.72 | 0.28 | 0.00 | 0.00 | 0.00 | 0.00 | 0.00 | 0.00 |
| Iran | 0.34 | 0.72 | 0.28 | 0.00 | 0.00 | 0.00 | 0.00 | 0.00 | 0.00 |
| Paraguay | 0.33 | 0.73 | 0.27 | 0.00 | 0.00 | 0.00 | 0.00 | 0.00 | 0.00 |
| Indonesia | 0.33 | 0.73 | 0.27 | 0.00 | 0.00 | 0.00 | 0.00 | 0.00 | 0.00 |
| Bhutan | 0.32 | 0.73 | 0.27 | 0.00 | 0.00 | 0.00 | 0.00 | 0.00 | 0.00 |
| India | 0.32 | 0.73 | 0.27 | 0.00 | 0.00 | 0.00 | 0.00 | 0.00 | 0.00 |
| Brunei | 0.32 | 0.73 | 0.27 | 0.00 | 0.00 | 0.00 | 0.00 | 0.00 | 0.00 |
| Myanmar (Burma) | 0.31 | 0.80 | 0.20 | 0.00 | 0.00 | 0.00 | 0.00 | 0.00 | 0.00 |
| Tonga | 0.31 | 0.80 | 0.20 | 0.00 | 0.00 | 0.00 | 0.00 | 0.00 | 0.00 |
| Nicaragua | 0.30 | 0.80 | 0.20 | 0.00 | 0.00 | 0.00 | 0.00 | 0.00 | 0.00 |
| Fiji | 0.30 | 0.80 | 0.20 | 0.00 | 0.00 | 0.00 | 0.00 | 0.00 | 0.00 |
| Bangladesh | 0.30 | 0.80 | 0.20 | 0.00 | 0.00 | 0.00 | 0.00 | 0.00 | 0.00 |
| French Guiana | 0.29 | 0.81 | 0.19 | 0.00 | 0.00 | 0.00 | 0.00 | 0.00 | 0.00 |
| Cape Verde | 0.29 | 0.81 | 0.19 | 0.00 | 0.00 | 0.00 | 0.00 | 0.00 | 0.00 |
| Belize | 0.28 | 0.81 | 0.19 | 0.00 | 0.00 | 0.00 | 0.00 | 0.00 | 0.00 |
| Philippines | 0.28 | 0.81 | 0.19 | 0.00 | 0.00 | 0.00 | 0.00 | 0.00 | 0.00 |
| Nepal | 0.28 | 0.81 | 0.19 | 0.00 | 0.00 | 0.00 | 0.00 | 0.00 | 0.00 |
| South Africa | 0.28 | 0.81 | 0.19 | 0.00 | 0.00 | 0.00 | 0.00 | 0.00 | 0.00 |
| Uzbekistan | 0.27 | 0.82 | 0.18 | 0.00 | 0.00 | 0.00 | 0.00 | 0.00 | 0.00 |
|  |  | [0 – .4) | [.4 – .8) | [.8 – 1.2) | [1.2 – 1.6) | [1.6 – 2.0) | [2.0 – 2.4) | [2.4 – 2.8) | [2.8 – ) |
| Samoa | 0.27 | 0.82 | 0.18 | 0.00 | 0.00 | 0.00 | 0.00 | 0.00 | 0.00 |
| Turkmenistan | 0.27 | 0.82 | 0.18 | 0.00 | 0.00 | 0.00 | 0.00 | 0.00 | 0.00 |
| Egypt | 0.27 | 0.82 | 0.18 | 0.00 | 0.00 | 0.00 | 0.00 | 0.00 | 0.00 |
| Honduras | 0.27 | 0.82 | 0.18 | 0.00 | 0.00 | 0.00 | 0.00 | 0.00 | 0.00 |
| Kyrgyzstan | 0.26 | 0.83 | 0.17 | 0.00 | 0.00 | 0.00 | 0.00 | 0.00 | 0.00 |
| Haiti | 0.26 | 0.83 | 0.17 | 0.00 | 0.00 | 0.00 | 0.00 | 0.00 | 0.00 |
| Libya | 0.26 | 0.83 | 0.17 | 0.00 | 0.00 | 0.00 | 0.00 | 0.00 | 0.00 |
| Guatemala | 0.26 | 0.83 | 0.17 | 0.00 | 0.00 | 0.00 | 0.00 | 0.00 | 0.00 |
| Syria | 0.26 | 0.86 | 0.14 | 0.00 | 0.00 | 0.00 | 0.00 | 0.00 | 0.00 |
| Mongolia | 0.26 | 0.86 | 0.14 | 0.00 | 0.00 | 0.00 | 0.00 | 0.00 | 0.00 |
| Djibouti | 0.25 | 0.87 | 0.13 | 0.00 | 0.00 | 0.00 | 0.00 | 0.00 | 0.00 |
| Lesotho | 0.25 | 0.87 | 0.13 | 0.00 | 0.00 | 0.00 | 0.00 | 0.00 | 0.00 |
| Kuwait | 0.25 | 0.87 | 0.13 | 0.00 | 0.00 | 0.00 | 0.00 | 0.00 | 0.00 |
| Cambodia | 0.25 | 0.87 | 0.13 | 0.00 | 0.00 | 0.00 | 0.00 | 0.00 | 0.00 |
| Micronesia | 0.24 | 0.89 | 0.11 | 0.00 | 0.00 | 0.00 | 0.00 | 0.00 | 0.00 |
| Kiribati | 0.24 | 0.89 | 0.11 | 0.00 | 0.00 | 0.00 | 0.00 | 0.00 | 0.00 |
| Saudi Arabia | 0.23 | 0.89 | 0.11 | 0.00 | 0.00 | 0.00 | 0.00 | 0.00 | 0.00 |
| Maldives | 0.23 | 0.89 | 0.11 | 0.00 | 0.00 | 0.00 | 0.00 | 0.00 | 0.00 |
| Botswana | 0.23 | 0.89 | 0.11 | 0.00 | 0.00 | 0.00 | 0.00 | 0.00 | 0.00 |
| Mayotte | 0.23 | 0.89 | 0.11 | 0.00 | 0.00 | 0.00 | 0.00 | 0.00 | 0.00 |
| Pakistan | 0.23 | 0.89 | 0.11 | 0.00 | 0.00 | 0.00 | 0.00 | 0.00 | 0.00 |
| Laos | 0.23 | 0.89 | 0.11 | 0.00 | 0.00 | 0.00 | 0.00 | 0.00 | 0.00 |
| Jordan | 0.22 | 0.89 | 0.11 | 0.00 | 0.00 | 0.00 | 0.00 | 0.00 | 0.00 |
| Timor-Leste | 0.22 | 0.89 | 0.11 | 0.00 | 0.00 | 0.00 | 0.00 | 0.00 | 0.00 |
| Western Sahara | 0.22 | 0.91 | 0.09 | 0.00 | 0.00 | 0.00 | 0.00 | 0.00 | 0.00 |
| Eritrea | 0.21 | 0.91 | 0.09 | 0.00 | 0.00 | 0.00 | 0.00 | 0.00 | 0.00 |
| Bahrain | 0.21 | 0.91 | 0.09 | 0.00 | 0.00 | 0.00 | 0.00 | 0.00 | 0.00 |
| Vanuatu | 0.20 | 0.91 | 0.09 | 0.00 | 0.00 | 0.00 | 0.00 | 0.00 | 0.00 |
| Solomon Islands | 0.20 | 0.91 | 0.09 | 0.00 | 0.00 | 0.00 | 0.00 | 0.00 | 0.00 |
| Swaziland | 0.20 | 0.91 | 0.09 | 0.00 | 0.00 | 0.00 | 0.00 | 0.00 | 0.00 |
| Namibia | 0.20 | 0.95 | 0.05 | 0.00 | 0.00 | 0.00 | 0.00 | 0.00 | 0.00 |
| Gabon | 0.20 | 0.95 | 0.05 | 0.00 | 0.00 | 0.00 | 0.00 | 0.00 | 0.00 |
| Papua New Guinea | 0.20 | 0.95 | 0.05 | 0.00 | 0.00 | 0.00 | 0.00 | 0.00 | 0.00 |
| Tajikistan | 0.19 | 0.96 | 0.04 | 0.00 | 0.00 | 0.00 | 0.00 | 0.00 | 0.00 |
| Sudan | 0.19 | 0.96 | 0.04 | 0.00 | 0.00 | 0.00 | 0.00 | 0.00 | 0.00 |
| Oman | 0.18 | 0.97 | 0.03 | 0.00 | 0.00 | 0.00 | 0.00 | 0.00 | 0.00 |
| Iraq | 0.18 | 0.97 | 0.03 | 0.00 | 0.00 | 0.00 | 0.00 | 0.00 | 0.00 |
| Ethiopia | 0.18 | 0.97 | 0.03 | 0.00 | 0.00 | 0.00 | 0.00 | 0.00 | 0.00 |
| Ghana | 0.18 | 0.97 | 0.03 | 0.00 | 0.00 | 0.00 | 0.00 | 0.00 | 0.00 |
| São Tomé & Príncipe | 0.18 | 0.97 | 0.03 | 0.00 | 0.00 | 0.00 | 0.00 | 0.00 | 0.00 |
| Palestinian Territories | 0.18 | 0.98 | 0.02 | 0.00 | 0.00 | 0.00 | 0.00 | 0.00 | 0.00 |
| Liberia | 0.18 | 0.98 | 0.02 | 0.00 | 0.00 | 0.00 | 0.00 | 0.00 | 0.00 |
| South Sudan | 0.18 | 0.98 | 0.02 | 0.00 | 0.00 | 0.00 | 0.00 | 0.00 | 0.00 |
| Mauritania | 0.18 | 0.98 | 0.02 | 0.00 | 0.00 | 0.00 | 0.00 | 0.00 | 0.00 |
| Comoros | 0.18 | 0.99 | 0.01 | 0.00 | 0.00 | 0.00 | 0.00 | 0.00 | 0.00 |
| Benin | 0.18 | 1.00 | 0.00 | 0.00 | 0.00 | 0.00 | 0.00 | 0.00 | 0.00 |
| Rwanda | 0.17 | 1.00 | 0.00 | 0.00 | 0.00 | 0.00 | 0.00 | 0.00 | 0.00 |
| Madagascar | 0.17 | 1.00 | 0.00 | 0.00 | 0.00 | 0.00 | 0.00 | 0.00 | 0.00 |
| Qatar | 0.17 | 1.00 | 0.00 | 0.00 | 0.00 | 0.00 | 0.00 | 0.00 | 0.00 |
| Senegal | 0.16 | 1.00 | 0.00 | 0.00 | 0.00 | 0.00 | 0.00 | 0.00 | 0.00 |
| Zimbabwe | 0.16 | 1.00 | 0.00 | 0.00 | 0.00 | 0.00 | 0.00 | 0.00 | 0.00 |
| United Arab Emirates | 0.16 | 1.00 | 0.00 | 0.00 | 0.00 | 0.00 | 0.00 | 0.00 | 0.00 |
| Sierra Leone | 0.16 | 1.00 | 0.00 | 0.00 | 0.00 | 0.00 | 0.00 | 0.00 | 0.00 |
| Congo - Brazzaville | 0.16 | 1.00 | 0.00 | 0.00 | 0.00 | 0.00 | 0.00 | 0.00 | 0.00 |
| Togo | 0.16 | 1.00 | 0.00 | 0.00 | 0.00 | 0.00 | 0.00 | 0.00 | 0.00 |
| Congo - Kinshasa | 0.16 | 1.00 | 0.00 | 0.00 | 0.00 | 0.00 | 0.00 | 0.00 | 0.00 |
| Yemen | 0.16 | 1.00 | 0.00 | 0.00 | 0.00 | 0.00 | 0.00 | 0.00 | 0.00 |
| Ivory Coast | 0.16 | 1.00 | 0.00 | 0.00 | 0.00 | 0.00 | 0.00 | 0.00 | 0.00 |
| Guinea | 0.15 | 1.00 | 0.00 | 0.00 | 0.00 | 0.00 | 0.00 | 0.00 | 0.00 |
| Guinea-Bissau | 0.15 | 1.00 | 0.00 | 0.00 | 0.00 | 0.00 | 0.00 | 0.00 | 0.00 |
| Central African Republic | 0.15 | 1.00 | 0.00 | 0.00 | 0.00 | 0.00 | 0.00 | 0.00 | 0.00 |
| Mozambique | 0.15 | 1.00 | 0.00 | 0.00 | 0.00 | 0.00 | 0.00 | 0.00 | 0.00 |
|  |  | [0 – .4) | [.4 – .8) | [.8 – 1.2) | [1.2 – 1.6) | [1.6 – 2.0) | [2.0 – 2.4) | [2.4 – 2.8) | [2.8 – ) |
| Somalia | 0.15 | 1.00 | 0.00 | 0.00 | 0.00 | 0.00 | 0.00 | 0.00 | 0.00 |
| Cameroon | 0.15 | 1.00 | 0.00 | 0.00 | 0.00 | 0.00 | 0.00 | 0.00 | 0.00 |
| Kenya | 0.15 | 1.00 | 0.00 | 0.00 | 0.00 | 0.00 | 0.00 | 0.00 | 0.00 |
| Nigeria | 0.15 | 1.00 | 0.00 | 0.00 | 0.00 | 0.00 | 0.00 | 0.00 | 0.00 |
| Tanzania | 0.15 | 1.00 | 0.00 | 0.00 | 0.00 | 0.00 | 0.00 | 0.00 | 0.00 |
| Afghanistan | 0.15 | 1.00 | 0.00 | 0.00 | 0.00 | 0.00 | 0.00 | 0.00 | 0.00 |
| Malawi | 0.14 | 1.00 | 0.00 | 0.00 | 0.00 | 0.00 | 0.00 | 0.00 | 0.00 |
| Equatorial Guinea | 0.14 | 1.00 | 0.00 | 0.00 | 0.00 | 0.00 | 0.00 | 0.00 | 0.00 |
| Gambia | 0.14 | 1.00 | 0.00 | 0.00 | 0.00 | 0.00 | 0.00 | 0.00 | 0.00 |
| Burkina Faso | 0.14 | 1.00 | 0.00 | 0.00 | 0.00 | 0.00 | 0.00 | 0.00 | 0.00 |
| Niger | 0.13 | 1.00 | 0.00 | 0.00 | 0.00 | 0.00 | 0.00 | 0.00 | 0.00 |
| Chad | 0.13 | 1.00 | 0.00 | 0.00 | 0.00 | 0.00 | 0.00 | 0.00 | 0.00 |
| Burundi | 0.13 | 1.00 | 0.00 | 0.00 | 0.00 | 0.00 | 0.00 | 0.00 | 0.00 |
| Mali | 0.13 | 1.00 | 0.00 | 0.00 | 0.00 | 0.00 | 0.00 | 0.00 | 0.00 |
| Angola | 0.13 | 1.00 | 0.00 | 0.00 | 0.00 | 0.00 | 0.00 | 0.00 | 0.00 |
| Zambia | 0.13 | 1.00 | 0.00 | 0.00 | 0.00 | 0.00 | 0.00 | 0.00 | 0.00 |
| Uganda | 0.12 | 1.00 | 0.00 | 0.00 | 0.00 | 0.00 | 0.00 | 0.00 | 0.00 |

**Table S7:**
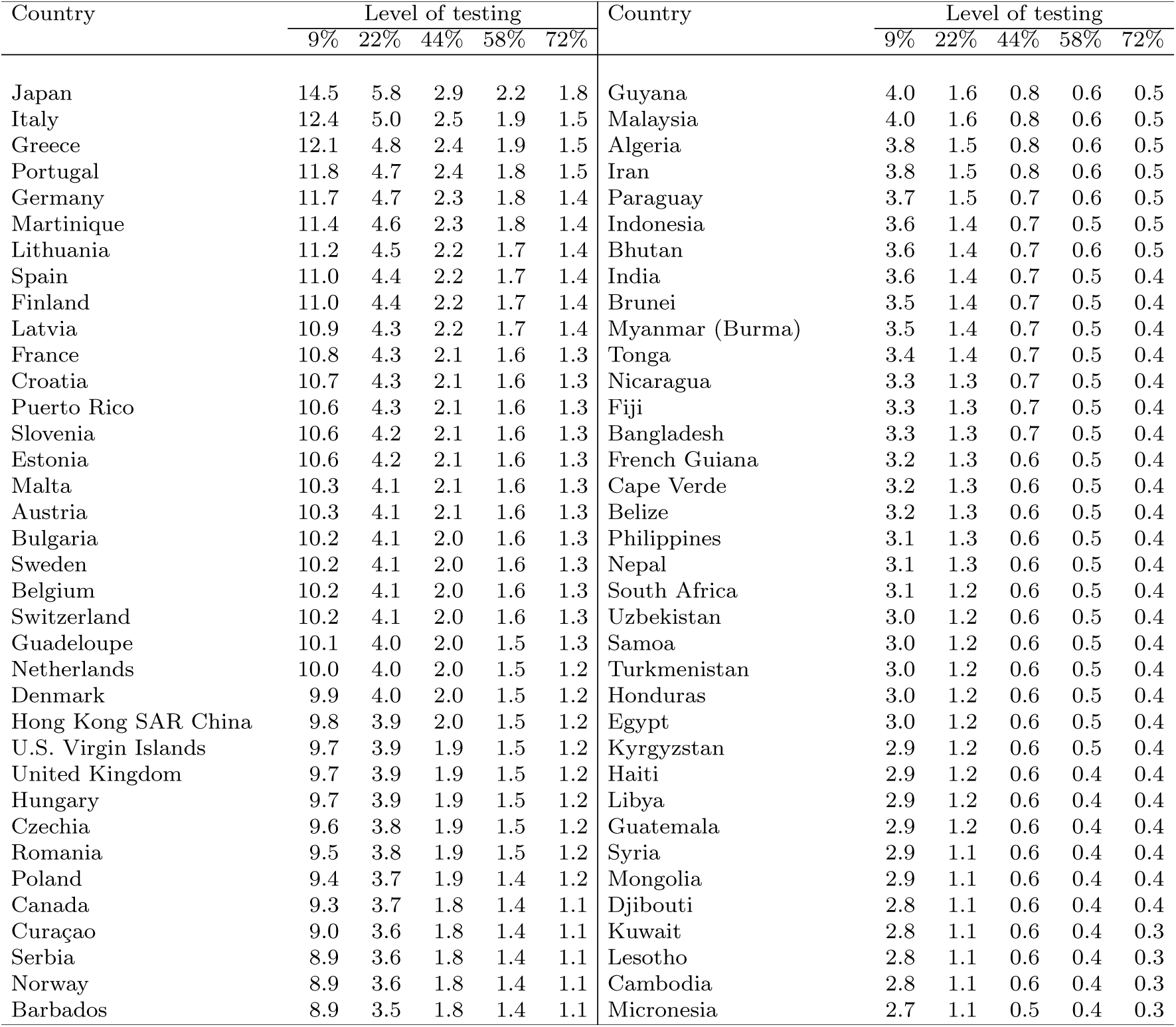
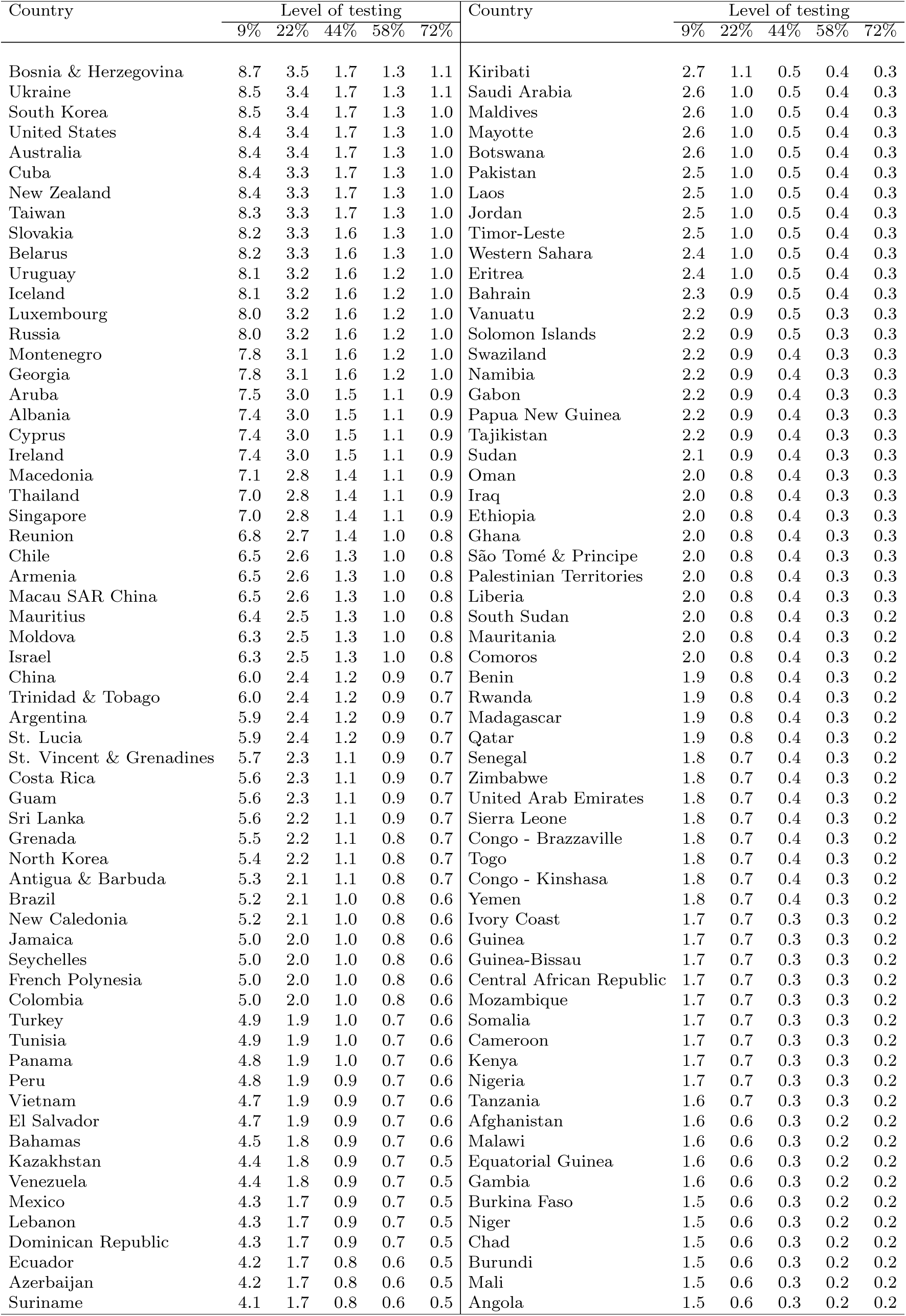
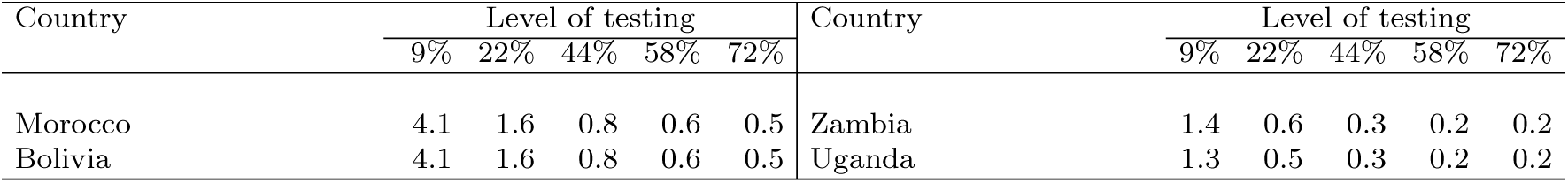
Average case fatality rate from COVID-19 after 365 days in fully susceptible populations by level of testing (in %): Day of intervention 45. Countries ordered from high to low case fatality rate level.

| Country | Level of testing |  |  |  |  | Country | Level of testing |  |  |  |  |
| --- | --- | --- | --- | --- | --- | --- | --- | --- | --- | --- | --- |
|  | 9% | 22% | 44% | 58% | 72% |  | 9% | 22% | 44% | 58% | 72% |
| Japan | 14.5 | 5.8 | 2.9 | 2.2 | 1.8 | Guyana | 4.0 | 1.6 | 0.8 | 0.6 | 0.5 |
| Italy | 12.4 | 5.0 | 2.5 | 1.9 | 1.5 | Malaysia | 4.0 | 1.6 | 0.8 | 0.6 | 0.5 |
| Greece | 12.1 | 4.8 | 2.4 | 1.9 | 1.5 | Algeria | 3.8 | 1.5 | 0.8 | 0.6 | 0.5 |
| Portugal | 11.8 | 4.7 | 2.4 | 1.8 | 1.5 | Iran | 3.8 | 1.5 | 0.8 | 0.6 | 0.5 |
| Germany | 11.7 | 4.7 | 2.3 | 1.8 | 1.4 | Paraguay | 3.7 | 1.5 | 0.7 | 0.6 | 0.5 |
| Martinique | 11.4 | 4.6 | 2.3 | 1.8 | 1.4 | Indonesia | 3.6 | 1.4 | 0.7 | 0.5 | 0.5 |
| Lithuania | 11.2 | 4.5 | 2.2 | 1.7 | 1.4 | Bhutan | 3.6 | 1.4 | 0.7 | 0.6 | 0.5 |
| Spain | 11.0 | 4.4 | 2.2 | 1.7 | 1.4 | India | 3.6 | 1.4 | 0.7 | 0.5 | 0.4 |
| Finland | 11.0 | 4.4 | 2.2 | 1.7 | 1.4 | Brunei | 3.5 | 1.4 | 0.7 | 0.5 | 0.4 |
| Latvia | 10.9 | 4.3 | 2.2 | 1.7 | 1.4 | Myanmar (Burma) | 3.5 | 1.4 | 0.7 | 0.5 | 0.4 |
| France | 10.8 | 4.3 | 2.1 | 1.6 | 1.3 | Tonga | 3.4 | 1.4 | 0.7 | 0.5 | 0.4 |
| Croatia | 10.7 | 4.3 | 2.1 | 1.6 | 1.3 | Nicaragua | 3.3 | 1.3 | 0.7 | 0.5 | 0.4 |
| Puerto Rico | 10.6 | 4.3 | 2.1 | 1.6 | 1.3 | Fiji | 3.3 | 1.3 | 0.7 | 0.5 | 0.4 |
| Slovenia | 10.6 | 4.2 | 2.1 | 1.6 | 1.3 | Bangladesh | 3.3 | 1.3 | 0.7 | 0.5 | 0.4 |
| Estonia | 10.6 | 4.2 | 2.1 | 1.6 | 1.3 | French Guiana | 3.2 | 1.3 | 0.6 | 0.5 | 0.4 |
| Malta | 10.3 | 4.1 | 2.1 | 1.6 | 1.3 | Cape Verde | 3.2 | 1.3 | 0.6 | 0.5 | 0.4 |
| Austria | 10.3 | 4.1 | 2.1 | 1.6 | 1.3 | Belize | 3.2 | 1.3 | 0.6 | 0.5 | 0.4 |
| Bulgaria | 10.2 | 4.1 | 2.0 | 1.6 | 1.3 | Philippines | 3.1 | 1.3 | 0.6 | 0.5 | 0.4 |
| Sweden | 10.2 | 4.1 | 2.0 | 1.6 | 1.3 | Nepal | 3.1 | 1.3 | 0.6 | 0.5 | 0.4 |
| Belgium | 10.2 | 4.1 | 2.0 | 1.6 | 1.3 | South Africa | 3.1 | 1.2 | 0.6 | 0.5 | 0.4 |
| Switzerland | 10.2 | 4.1 | 2.0 | 1.6 | 1.3 | Uzbekistan | 3.0 | 1.2 | 0.6 | 0.5 | 0.4 |
| Guadeloupe | 10.1 | 4.0 | 2.0 | 1.5 | 1.3 | Samoa | 3.0 | 1.2 | 0.6 | 0.5 | 0.4 |
| Netherlands | 10.0 | 4.0 | 2.0 | 1.5 | 1.2 | Turkmenistan | 3.0 | 1.2 | 0.6 | 0.5 | 0.4 |
| Denmark | 9.9 | 4.0 | 2.0 | 1.5 | 1.2 | Honduras | 3.0 | 1.2 | 0.6 | 0.5 | 0.4 |
| Hong Kong SAR China | 9.8 | 3.9 | 2.0 | 1.5 | 1.2 | Egypt | 3.0 | 1.2 | 0.6 | 0.5 | 0.4 |
| U.S. Virgin Islands | 9.7 | 3.9 | 1.9 | 1.5 | 1.2 | Kyrgyzstan | 2.9 | 1.2 | 0.6 | 0.5 | 0.4 |
| United Kingdom | 9.7 | 3.9 | 1.9 | 1.5 | 1.2 | Haiti | 2.9 | 1.2 | 0.6 | 0.4 | 0.4 |
| Hungary | 9.7 | 3.9 | 1.9 | 1.5 | 1.2 | Libya | 2.9 | 1.2 | 0.6 | 0.4 | 0.4 |
| Czechia | 9.6 | 3.8 | 1.9 | 1.5 | 1.2 | Guatemala | 2.9 | 1.2 | 0.6 | 0.4 | 0.4 |
| Romania | 9.5 | 3.8 | 1.9 | 1.5 | 1.2 | Syria | 2.9 | 1.1 | 0.6 | 0.4 | 0.4 |
| Poland | 9.4 | 3.7 | 1.9 | 1.4 | 1.2 | Mongolia | 2.9 | 1.1 | 0.6 | 0.4 | 0.4 |
| Canada | 9.3 | 3.7 | 1.8 | 1.4 | 1.1 | Djibouti | 2.8 | 1.1 | 0.6 | 0.4 | 0.4 |
| Curaçao | 9.0 | 3.6 | 1.8 | 1.4 | 1.1 | Kuwait | 2.8 | 1.1 | 0.6 | 0.4 | 0.3 |
| Serbia | 8.9 | 3.6 | 1.8 | 1.4 | 1.1 | Lesotho | 2.8 | 1.1 | 0.6 | 0.4 | 0.3 |
| Norway | 8.9 | 3.6 | 1.8 | 1.4 | 1.1 | Cambodia | 2.8 | 1.1 | 0.6 | 0.4 | 0.3 |
| Barbados | 8.9 | 3.5 | 1.8 | 1.4 | 1.1 | Micronesia | 2.7 | 1.1 | 0.5 | 0.4 | 0.3 |
|  | 9% | 22% | 44% | 58% | 72% |  | 9% | 22% | 44% | 58% | 72% |
| Bosnia & Herzegovina | 8.7 | 3.5 | 1.7 | 1.3 | 1.1 | Kiribati | 2.7 | 1.1 | 0.5 | 0.4 | 0.3 |
| Ukraine | 8.5 | 3.4 | 1.7 | 1.3 | 1.1 | Saudi Arabia | 2.6 | 1.0 | 0.5 | 0.4 | 0.3 |
| South Korea | 8.5 | 3.4 | 1.7 | 1.3 | 1.0 | Maldives | 2.6 | 1.0 | 0.5 | 0.4 | 0.3 |
| United States | 8.4 | 3.4 | 1.7 | 1.3 | 1.0 | Mayotte | 2.6 | 1.0 | 0.5 | 0.4 | 0.3 |
| Australia | 8.4 | 3.4 | 1.7 | 1.3 | 1.0 | Botswana | 2.6 | 1.0 | 0.5 | 0.4 | 0.3 |
| Cuba | 8.4 | 3.3 | 1.7 | 1.3 | 1.0 | Pakistan | 2.5 | 1.0 | 0.5 | 0.4 | 0.3 |
| New Zealand | 8.4 | 3.3 | 1.7 | 1.3 | 1.0 | Laos | 2.5 | 1.0 | 0.5 | 0.4 | 0.3 |
| Taiwan | 8.3 | 3.3 | 1.7 | 1.3 | 1.0 | Jordan | 2.5 | 1.0 | 0.5 | 0.4 | 0.3 |
| Slovakia | 8.2 | 3.3 | 1.6 | 1.3 | 1.0 | Timor-Leste | 2.5 | 1.0 | 0.5 | 0.4 | 0.3 |
| Belarus | 8.2 | 3.3 | 1.6 | 1.3 | 1.0 | Western Sahara | 2.4 | 1.0 | 0.5 | 0.4 | 0.3 |
| Uruguay | 8.1 | 3.2 | 1.6 | 1.2 | 1.0 | Eritrea | 2.4 | 1.0 | 0.5 | 0.4 | 0.3 |
| Iceland | 8.1 | 3.2 | 1.6 | 1.2 | 1.0 | Bahrain | 2.3 | 0.9 | 0.5 | 0.4 | 0.3 |
| Luxembourg | 8.0 | 3.2 | 1.6 | 1.2 | 1.0 | Vanuatu | 2.2 | 0.9 | 0.5 | 0.3 | 0.3 |
| Russia | 8.0 | 3.2 | 1.6 | 1.2 | 1.0 | Solomon Islands | 2.2 | 0.9 | 0.5 | 0.3 | 0.3 |
| Montenegro | 7.8 | 3.1 | 1.6 | 1.2 | 1.0 | Swaziland | 2.2 | 0.9 | 0.4 | 0.3 | 0.3 |
| Georgia | 7.8 | 3.1 | 1.6 | 1.2 | 1.0 | Namibia | 2.2 | 0.9 | 0.4 | 0.3 | 0.3 |
| Aruba | 7.5 | 3.0 | 1.5 | 1.1 | 0.9 | Gabon | 2.2 | 0.9 | 0.4 | 0.3 | 0.3 |
| Albania | 7.4 | 3.0 | 1.5 | 1.1 | 0.9 | Papua New Guinea | 2.2 | 0.9 | 0.4 | 0.3 | 0.3 |
| Cyprus | 7.4 | 3.0 | 1.5 | 1.1 | 0.9 | Tajikistan | 2.2 | 0.9 | 0.4 | 0.3 | 0.3 |
| Ireland | 7.4 | 3.0 | 1.5 | 1.1 | 0.9 | Sudan | 2.1 | 0.9 | 0.4 | 0.3 | 0.3 |
| Macedonia | 7.1 | 2.8 | 1.4 | 1.1 | 0.9 | Oman | 2.0 | 0.8 | 0.4 | 0.3 | 0.3 |
| Thailand | 7.0 | 2.8 | 1.4 | 1.1 | 0.9 | Iraq | 2.0 | 0.8 | 0.4 | 0.3 | 0.3 |
| Singapore | 7.0 | 2.8 | 1.4 | 1.1 | 0.9 | Ethiopia | 2.0 | 0.8 | 0.4 | 0.3 | 0.3 |
| Reunion | 6.8 | 2.7 | 1.4 | 1.0 | 0.8 | Ghana | 2.0 | 0.8 | 0.4 | 0.3 | 0.3 |
| Chile | 6.5 | 2.6 | 1.3 | 1.0 | 0.8 | São Tomé & Príncipe | 2.0 | 0.8 | 0.4 | 0.3 | 0.3 |
| Armenia | 6.5 | 2.6 | 1.3 | 1.0 | 0.8 | Palestinian Territories | 2.0 | 0.8 | 0.4 | 0.3 | 0.3 |
| Macau SAR China | 6.5 | 2.6 | 1.3 | 1.0 | 0.8 | Liberia | 2.0 | 0.8 | 0.4 | 0.3 | 0.3 |
| Mauritius | 6.4 | 2.5 | 1.3 | 1.0 | 0.8 | South Sudan | 2.0 | 0.8 | 0.4 | 0.3 | 0.2 |
| Moldova | 6.3 | 2.5 | 1.3 | 1.0 | 0.8 | Mauritania | 2.0 | 0.8 | 0.4 | 0.3 | 0.2 |
| Israel | 6.3 | 2.5 | 1.3 | 1.0 | 0.8 | Comoros | 2.0 | 0.8 | 0.4 | 0.3 | 0.2 |
| China | 6.0 | 2.4 | 1.2 | 0.9 | 0.7 | Benin | 1.9 | 0.8 | 0.4 | 0.3 | 0.2 |
| Trinidad & Tobago | 6.0 | 2.4 | 1.2 | 0.9 | 0.7 | Rwanda | 1.9 | 0.8 | 0.4 | 0.3 | 0.2 |
| Argentina | 5.9 | 2.4 | 1.2 | 0.9 | 0.7 | Madagascar | 1.9 | 0.8 | 0.4 | 0.3 | 0.2 |
| St. Lucia | 5.9 | 2.4 | 1.2 | 0.9 | 0.7 | Qatar | 1.9 | 0.8 | 0.4 | 0.3 | 0.2 |
| St. Vincent & Grenadines | 5.7 | 2.3 | 1.1 | 0.9 | 0.7 | Senegal | 1.8 | 0.7 | 0.4 | 0.3 | 0.2 |
| Costa Rica | 5.6 | 2.3 | 1.1 | 0.9 | 0.7 | Zimbabwe | 1.8 | 0.7 | 0.4 | 0.3 | 0.2 |
| Guam | 5.6 | 2.3 | 1.1 | 0.9 | 0.7 | United Arab Emirates | 1.8 | 0.7 | 0.4 | 0.3 | 0.2 |
| Sri Lanka | 5.6 | 2.2 | 1.1 | 0.9 | 0.7 | Sierra Leone | 1.8 | 0.7 | 0.4 | 0.3 | 0.2 |
| Grenada | 5.5 | 2.2 | 1.1 | 0.8 | 0.7 | Congo - Brazzaville | 1.8 | 0.7 | 0.4 | 0.3 | 0.2 |
| North Korea | 5.4 | 2.2 | 1.1 | 0.8 | 0.7 | Togo | 1.8 | 0.7 | 0.4 | 0.3 | 0.2 |
| Antigua & Barbuda | 5.3 | 2.1 | 1.1 | 0.8 | 0.7 | Congo - Kinshasa | 1.8 | 0.7 | 0.4 | 0.3 | 0.2 |
| Brazil | 5.2 | 2.1 | 1.0 | 0.8 | 0.6 | Yemen | 1.8 | 0.7 | 0.4 | 0.3 | 0.2 |
| New Caledonia | 5.2 | 2.1 | 1.0 | 0.8 | 0.6 | Ivory Coast | 1.7 | 0.7 | 0.3 | 0.3 | 0.2 |
| Jamaica | 5.0 | 2.0 | 1.0 | 0.8 | 0.6 | Guinea | 1.7 | 0.7 | 0.3 | 0.3 | 0.2 |
| Seychelles | 5.0 | 2.0 | 1.0 | 0.8 | 0.6 | Guinea-Bissau | 1.7 | 0.7 | 0.3 | 0.3 | 0.2 |
| French Polynesia | 5.0 | 2.0 | 1.0 | 0.8 | 0.6 | Central African Republic | 1.7 | 0.7 | 0.3 | 0.3 | 0.2 |
| Colombia | 5.0 | 2.0 | 1.0 | 0.8 | 0.6 | Mozambique | 1.7 | 0.7 | 0.3 | 0.3 | 0.2 |
| Turkey | 4.9 | 1.9 | 1.0 | 0.7 | 0.6 | Somalia | 1.7 | 0.7 | 0.3 | 0.3 | 0.2 |
| Tunisia | 4.9 | 1.9 | 1.0 | 0.7 | 0.6 | Cameroon | 1.7 | 0.7 | 0.3 | 0.3 | 0.2 |
| Panama | 4.8 | 1.9 | 1.0 | 0.7 | 0.6 | Kenya | 1.7 | 0.7 | 0.3 | 0.3 | 0.2 |
| Peru | 4.8 | 1.9 | 0.9 | 0.7 | 0.6 | Nigeria | 1.7 | 0.7 | 0.3 | 0.3 | 0.2 |
| Vietnam | 4.7 | 1.9 | 0.9 | 0.7 | 0.6 | Tanzania | 1.6 | 0.7 | 0.3 | 0.3 | 0.2 |
| El Salvador | 4.7 | 1.9 | 0.9 | 0.7 | 0.6 | Afghanistan | 1.6 | 0.6 | 0.3 | 0.2 | 0.2 |
| Bahamas | 4.5 | 1.8 | 0.9 | 0.7 | 0.6 | Malawi | 1.6 | 0.6 | 0.3 | 0.2 | 0.2 |
| Kazakhstan | 4.4 | 1.8 | 0.9 | 0.7 | 0.5 | Equatorial Guinea | 1.6 | 0.6 | 0.3 | 0.2 | 0.2 |
| Venezuela | 4.4 | 1.8 | 0.9 | 0.7 | 0.5 | Gambia | 1.6 | 0.6 | 0.3 | 0.2 | 0.2 |
| Mexico | 4.3 | 1.7 | 0.9 | 0.7 | 0.5 | Burkina Faso | 1.5 | 0.6 | 0.3 | 0.2 | 0.2 |
| Lebanon | 4.3 | 1.7 | 0.9 | 0.7 | 0.5 | Niger | 1.5 | 0.6 | 0.3 | 0.2 | 0.2 |
| Dominican Republic | 4.3 | 1.7 | 0.9 | 0.7 | 0.5 | Chad | 1.5 | 0.6 | 0.3 | 0.2 | 0.2 |
| Ecuador | 4.2 | 1.7 | 0.8 | 0.6 | 0.5 | Burundi | 1.5 | 0.6 | 0.3 | 0.2 | 0.2 |
| Azerbaijan | 4.2 | 1.7 | 0.8 | 0.6 | 0.5 | Mali | 1.5 | 0.6 | 0.3 | 0.2 | 0.2 |
| Suriname | 4.1 | 1.7 | 0.8 | 0.6 | 0.5 | Angola | 1.5 | 0.6 | 0.3 | 0.2 | 0.2 |
|  | 9% | 22% | 44% | 58% | 72% |  | 9% | 22% | 44% | 58% | 72% |
| Morocco | 4.1 | 1.6 | 0.8 | 0.6 | 0.5 | Zambia | 1.4 | 0.6 | 0.3 | 0.2 | 0.2 |
| Bolivia | 4.1 | 1.6 | 0.8 | 0.6 | 0.5 | Uganda | 1.3 | 0.5 | 0.3 | 0.2 | 0.2 |

**Table S8:**
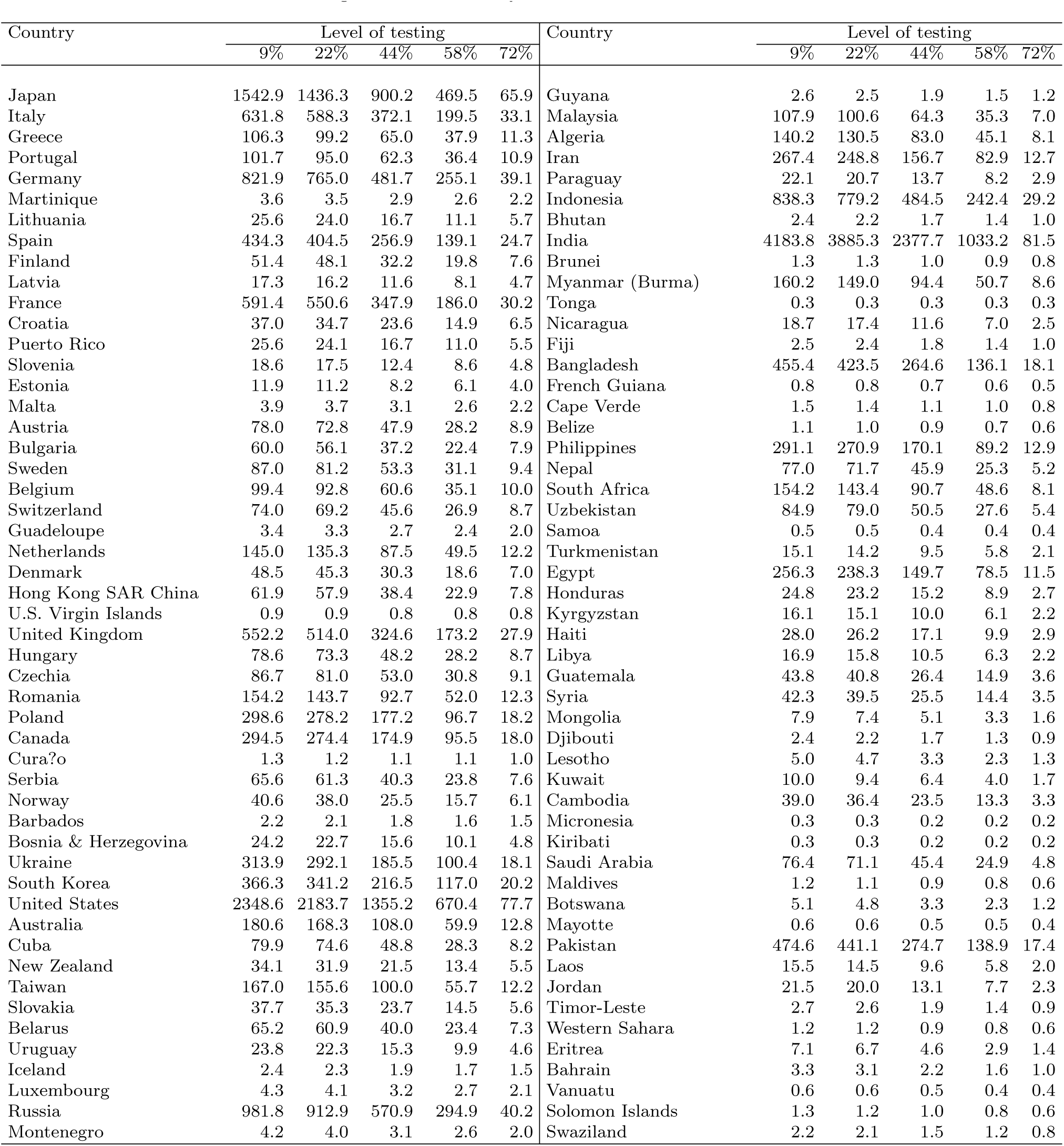
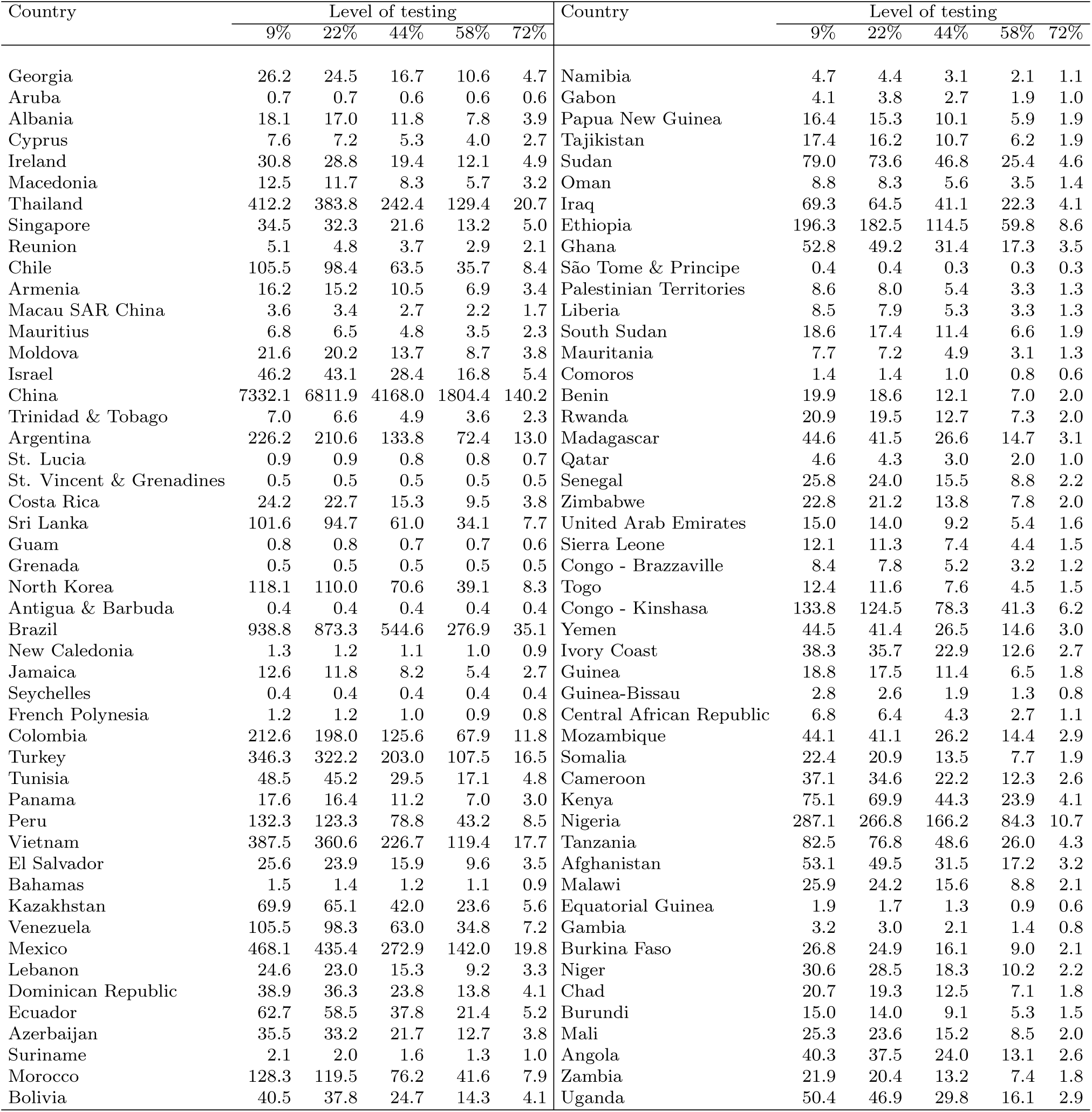
Average death toll (in 1000s) from COVID-19 by level of testing in fully susceptible populations after 365 days: Day of intervention 45. Countries ordered from high to low case fatality rate level.

**Figure S13:**
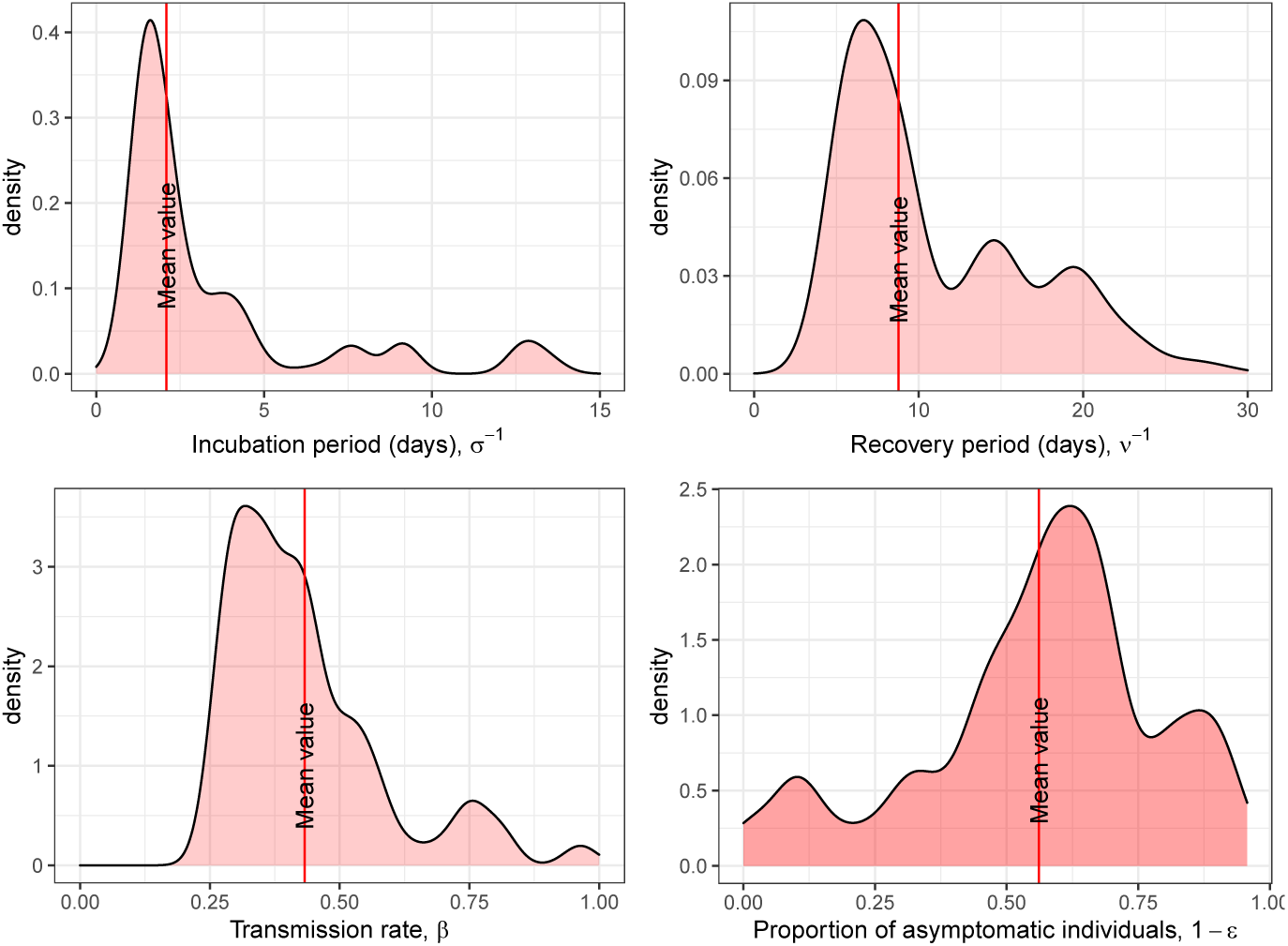
Marginal posterior distribution of the parameters of the SEIR mode (Θ).

## Footnotes

1 South Korea, Taiwan, Singapore, Germany, Iceland and Vò employed aggressive testing; the first three Asian countries had health protocols based on previous epidemic experience that enabled them to act quickly, while Germany had extensive testing availability and both Iceland and Vò are small, the latter a 3,000 inhabitants Italian town.

2 The proportion of lives saved is defined as the relative difference between the total deaths without testing and the total deaths for a specific level of testing (See Tables S2 and S8).

3 Our estimated CFR values represent the figures that would have been observed, for a given level of testing, after one whole year since the onset of the epidemic outbreak. When a country maintains a specific level of testing over time, the CFR increases monotonically over time until reaching a maximum. Thus, by using the CFR at the end of the pandemic we avoid the discrepancies that may result from changing testing policies, such as those happening because of test shortage.

1 Weassumethatthelikelihoodthatasusceptiblemeetsaninfectiousindividualisreducedbythenumber of exposed and infectious individuals that are detected and isolated 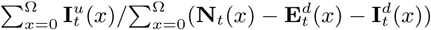 similar to(4).

2 The graphs with the smoothed mortality profiles are available for all countries upon request.

3 Since no isolation measures were implemented during the period modeled, we set *π_E_* and *π*_I_ to zero for the calibration.

4 The final support for the parameters {*σ*, *ν*, *β*, *ε* } were chosen after a first run of the Bayesian Melding with a sample of more than one million simulations.

5 Our model *M*(Θ) might not be invertible because the parameters *β* and *σ* are highlycorrelated.

6 The error distribution of *e_s_* is the result of assuming that the difference between the model's deaths and the observed deaths at anytime *t* is i.i.d. according to a 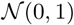.

7 See Figure S13 for a graphical representation of the marginal posterior distributions of the model parameters.

8 Recall that we have ignored states **E***^d^* and **I**^d^ in our calibration, since isolation measures were not implemented during the period analyzed. This strategy reduces the potential bias that results from estimating these parameters in countries with unknown imported cases and/or in countries with undocumented cases that have implemented testing and isolation measures.

9 The case fatality rate calculated as the ratio between the total number of deaths and the total number of infected and detected cases.

10 Table S2 and Figure S8 only reports average fatality rate values. The distribution of fatality rates for each country is reported in Table S6.

11 Given the existence of tests shortages, it is also true that countries with smaller populations may have an advantage over countries with large populations because they do not need to buy as many tests.

12 When a country maintains a specific level of testing, the CFR increases monotonically over time until reaching a maximum. This is because death might occur days after the individual is infected and detected. Thus, by using the CFR at the end of the pandemic we avoid the discrepancies that may result from changing testing policies, such as those happening because of the shortage of tests.

13 Nonetheless, these numbers should be taken with caution since they are also a function of the quality of the health care system.

